# Polygenic predisposition to venous thromboembolism is associated with increased COVID-19 positive testing rates

**DOI:** 10.1101/2022.01.29.22270094

**Authors:** Jessica Minnier, Jennifer E Huffman, Lina Gao, Jacob Joseph, Emily S Wan, Wen-Chih Wu, Ayako Suzuki, Gita A Pathak, Renato Polimanti, Mehrdad Arjomandi, Kyong-Mi Chang, Helene Garcon, Anurag Verma, Yuk-Lam Ho, James B Meigs, Kelly Cho, Robert A Bonomo, Bryan R Gorman, Saiju Pyarajan, Elise Gatsby, Nallakkandi Rajeevan, Kristine E Lynch, Julie A Lynch, Seyedeh Maryam Zekavat, Pradeep Natarajan, Cecelia J Madison, Jin J Zhou, Darshana N Jhala, Curtis J Donskey, John E McGeary, Peter D Reaven, Yan V Sun, Mat Freiberg, Joel Gelernter, Jeffrey M Petersen, Adriana Hung, Rose DL Huang, Ravi K Madduri, Sharvari Dalal, Quinn S Wells, Katherine P Liao, Peter W.F. Wilson, Philip S Tsao, Christopher J O’Donnell, John M Gaziano, VA Million Veteran Program, Richard L Hauger, Sudha K. Iyengar, Shiuh-Wen Luoh

**Affiliations:** OHSU-PSU School of Public Health, Oregon Health & Science University, Portland, OR, 97239, USA; Knight Cancer Institute, Biostatistics Shared Resource, Oregon Health & Science University, Portland, OR, 97239, USA; VA Portland Health Care System, Portland, OR, 97239, USA; MAVERIC, VA Boston Healthcare System, 150 S Huntington Ave, Boston, MA, 02130, USA; Department of Medicine, VA Boston Healthcare System, Boston, MA, 02132, USA; Brigham & Women’s Hospital, Boston, MA, 02115, USA; Department of Medicine, Pulmonary, Critical Care, Sleep, and Allergy Section, VA Boston Healthcare System, 150 S Huntington Ave, Boston, MA, 02130, USA; Channing Division of Network Medicine, Brigham & Women’s Hospital, Boston, MA, 02115, USA; Department of Medicine, Cardiology, Providence VA Healthcare System, Providence, RI, 02908, USA; Alpert Medical School & School of Public Health, Brown University, Providence, RI, 02903, USA; Department of Medicine, Gastroenterology, Durham VA Medical Center, 508 Fulton St, Durham, NC, 27705, USA; Department of Medicine, Gastroenterology, Duke University, Durham, NC, 27710, USA; Department of Psychiatry, Division of Human Genetics, Yale School of Medicine, New Haven, CT, 06511, USA; VA Connecticut Healthcare System, West Haven, CT, 06516, USA; Medicine, Pulmonary and Critical Care, San Francisco VA Healthcare System; University of California San Francisco, San Francisco, CA, 94121, USA; Corporal Michael J Crescenz VA Medical Center, Philadelphia, PA, 19104, USA; Department of Medicine, Perelman School of Medicine, University of Pennsylvania, Philadelphia, PA, 19104, USA; Medicine, General Internal Medicine, Massachusetts General Hospital, 55 Fruit St, Boston, MA, 02114, USA; Medicine, Aging, Brigham and Women’s Hospital, Harvard Medical School, Boston, MA, 02115, USA; Louis Stokes Cleveland VA, Cleveland, OH, 44106, USA; VA Boston Healthcare System, 150 S Huntington Ave, Boston, MA, 02130, USA; Department of Medicine, Harvard Medical School, Boston, MA, 02115, USA; VA Informatics and Computing Infrastructure (VINCI), VA Salt Lake City Healthcare System, Salt Lake City, UT, USA; Internal Medicine, Epidemiology, University of Utah School of Medicine, Salt Lake City, UT, USA; Yale Center for Medical Informatics, Yale School of Medicine, New Haven, CT, 06511, USA; Clinical Epidemiology Research Center (CERC), VA Connecticut Healthcare System, West Haven, CT, 06516, USA; VA Informatics and Computing Infrastructure (VINCI), VA Salt Lake City Healthcare System, Salt Lake City, UT, 84148, USA; VA Informatics & Computing Infrastructure, VA Salt Lake City Health Care System, Salt Lake City, UT, 84148, USA; Computational Biology & Bioinformatics, Yale School of Medicine, 333 Cedar St, New Haven, CT, 06510, USA; Program in Medical and Population Genetics, Cardiovascular Disease Initiative, Broad Institute of Harvard and MIT, Cambridge, MA, 02142, USA; Cardiovascular Research Center, Massachusetts General Hospital, 55 Fruit St, Boston, MA, 02114, USA; Clinical Data Science Research Group, ORD, Portland VA Medical Center, Portland, OR, 97239, USA; Medicine, University of California, Los Angeles, Los Angeles, CA, 90024, USA; Epidemiology and Biostatistics, University of Arizona, AZ, 85724, USA; Pathology and Laboratory Medicine, Corporal Michael J Crescenz VA Medical Center, Philadelphia, PA, 19104, USA; Perelman School of Medicine, University of Pennsylvania, 19104, USA; Infectious Disease Section, Louis Stokes Cleveland VA and Case Western Reserve University, Cleveland, OH, 44106, USA; Department of Psychiatry and Human Behavior, Providence VA Medical Center, Providence, RI, 02908, USA; Brown University Medical School, USA; Deptartment of Medicine, Phoenix VA Healthcare System, Phoenix, AZ, 85012, USA; Univ. of AZ, USA; Atlanta VA Health Care System, 1670 Clairmont Road, Decatur, GA, 30033, USA; Epidemiology, Emory University School of Public Health, 1518 Clifton Rd. NE, Atlanta, GA, 30322, USA; Vanderbilt University Medical Center, Nashville, TN, 37232, USA; Psychiatry, Human Genetics, Yale School of Medicine, 950 Campbell Avenue, West Haven, CT, 06516, USA; VA CT Healthcare Center, USA; Perelman School of Medicine, University of Pennsylvania, Philadelphia, PA, 19104, USA; Tennesse Valley Healthcare System, Nashville, TN, 37212, USA; Data Science and Learning, Argonne National Laboratory, 9700 S Cass Ave, Lemont, IL, 60439, USA; Departments of Medicine, Biomedical Informatics, and Pharmacology, Vanderbilt University Medical Center, Nashville, TN, 37232, USA; Medicine, Rheumatology, VA Boston Healthcare System, 150 S Huntington Ave, Boston, MA, 02130, USA; Emory University School of Medicine, Atlanta, GA, 30322, USA; Precision Medicine, VA Palo Alto Health Care System, 3801 Miranda Avenue, Palo Alto, CA, 94304, USA; Medicine, Cardiology, VA Boston Healthcare System, 1400 VFW Parkway, Boston, MA, 02132, USA; VA Boston Healthcare System, 1400 VFW Parkway, Boston, MA, 02132, USA; Center of Excellence for Stress & Mental Health, VA San Diego Healthcare System, San Diego, CA, 92161, USA; Center for Behavioral Genetics of Aging, University of California San Diego, La Jolla, CA, 92093, USA; Case Western Reserve University, Cleveland, OH, 44106, USA; Louis Stokes Cleveland VA Medical Center, Cleveland, OH, 44106, USA; Knight Cancer Institute, Oregon Health & Science University, Portland, OR, 97239, USA

## Abstract

Genetic predisposition to venous thrombosis may impact COVID-19 infection and its sequelae. Participants in the ongoing prospective cohort study, Million Veteran Program (MVP), who were tested for COVID-19, with European ancestry, were evaluated for associations with polygenic venous thromboembolic risk, Factor V Leiden mutation (FVL) (rs6025) and prothrombin gene 3’ -UTR mutation (F2 G20210A)(rs1799963), and their interactions. Logistic regression models assessed genetic associations with VTE diagnosis, COVID-19 (positive) testing rates and outcome severity (modified WHO criteria), and post-test conditions, adjusting for outpatient anticoagulation medication usage, age, sex, and genetic principal components. 108,437 out of 464,961 European American MVP participants were tested for COVID-19 with 9786 (9%) positive. PRS(VTE), FVL, *F2* G20210A were not significantly associated with the propensity of being tested for COVID-19. PRS(VTE) was significantly associated with a positive COVID-19 test in *F5* wild type (WT) individuals (OR 1.05; 95% CI [1.02-1.07]), but not in FVL carriers (0.97, [0.91-1.94]). There was no association with severe outcome for FVL, *F2* G20210A or PRS(VTE). Outpatient anticoagulation usage in the two years prior to testing was associated with worse clinical outcomes. PRS(VTE) was associated with prevalent VTE diagnosis among both FVL carriers or *F5* wild type individuals as well as incident VTE in the two years prior to testing. Increased genetic propensity for VTE in the MVP was associated with increased COVID-19 positive testing rates, suggesting a role of coagulation in the initial steps of COVID-19 infection.

**Key Points:**

- Increased genetic predisposition to venous thrombosis is associated with increased COVID-19 positive testing rates.
- PRS for VTE further risk stratifies factor V Leiden carriers regarding their VTE risk.

## Introduction

Venous and arterial micro- and macro-thrombosis are common manifestations of severe COVID-19 infection^1^. Venous thromboembolism (VTE) is the most commonly reported thrombotic complication, with higher incidence rates among critically ill patients. One review estimated that up to 28% of critically ill patients with COVID-19 had VTE^2^. Increased levels of fibrinogen, fibrin, D-dimers and fibrin degradation products were seen in COVID-19 infection^3^. Both in-situ immuno-thrombosis and “classical” pulmonary thromboembolism are recognized to be associated with the development of pulmonary vascular occlusion^4–7^. Mechanistically, both endothelial injury and a prothrombotic milieu may contribute to the COVID-19 associated thrombosis^4,8,9^.

Factor V Leiden mutation (FVL) results from a point mutation (rs6025) in the *F5* gene, which encodes the factor V protein in the coagulation cascade^10^. FVL, which is inherited in an autosomal dominant fashion, renders factor V protein insensitive to the cleavage and inactivation by the activated protein C (aPC), a natural anticoagulant. As a result, individuals who carry one or two copies of the FVL variant are at elevated risk for venous thromboembolism (VTE)^10^. Prothrombin G20210A is the second most common inherited thrombophilia after factor V Leiden. The G20210A point mutation (rs17999863) results from a substitution of adenine (A) for guanine (G) at position 20210 in a non-coding region of the *F2* gene^11^.Corresponding to the terminal nucleotide at the 3’ untranslated region of the gene, prothrombin G20210A is a gain of function mutation with an approximately 30% higher prothrombin protein level in the blood^12^. Although both FVL and prothrombin G20210A are very common among populations of European descent, many carriers will never have a VTE. Additional genetic and/or environmental insults are likely required to trigger VTE events among these carriers^13^.

In addition to other rarer monogenic mutations that are associated with inherited hypercoagulable states such as protein C, Protein S and antithrombin deficiency^14^, there are a number of other genetic variants associated with an increased risk for VTEs which have been discovered by both candidate as well as genome wide association studies (GWAS). Though individually each of these variants was associated with a small risk of VTE, together they conferred increased VTE risk comparable to that of FVL. One such polygenic risk score (PRS) for VTE has been previously established and validated by MVP; individuals that were FVL carriers and had the top 5% PRS(VTE) values had the highest risk for VTE^15^.

In this study we investigated whether genetic predisposition to VTEs, either due to one of the most common monogenic mutations such as FVL or Prothrombin G20210A or higher polygenic risk scores (PRS) for VTE will impact on the COVID-19 testing positive and disease severity. We performed a study in the Million Veteran Program (MVP)^16^ funded by Department of the Veterans Affairs (VA) which includes a comprehensive electronic health record system (EHR) with documented data for pre-COVID conditions, as well as post-COVID acute events and genotyping status for *F2, F5* and PRS in a cohort of over 650,000 veterans. This large database thus offers an opportunity to study the effect of genetic predisposition to PRS(VTE) on COVID-19 infection and its severity in a greater detail and a prospective manner.

## Materials and methods

### Study design and participants

This study included 464,961 participants from the Million Veteran Program who had European ancestry with available genetic data (release 4), 108,437 of which were tested for COVID-19. The MVP cohort has been described elsewhere^16,17^, with details in supplementary material. MVP participants who were tested for COVID-19 within the VA healthcare system between March 1, 2020 and June 2, 2021 were included, with follow-up data studied until September 2, 2021.

COVID positive testing rate and prevalent VTE were studied in the cohort of participants tested for COVID-19. COVID severity was studied in participants who tested positive within the VA healthcare system by June 2, 2021 and who had severity measurements present at the September 22, 2021 follow up date. To evaluate VTE within the pre-index period up to 2 years prior to testing, the cohort included all COVID-tested participants with the exclusion of patients who had taken outpatient anticoagulation medication during the year before pre-index period (2 to 3 years prior to testing) as well as participants who had a history of VTE prior to the pre-index period.

All analyses were conducted using release 4 of the MVP genetic data; sensitivity analyses were conducted excluding participants from the genetic release v2.1 used for the training of the PRS(VTE)^15^ with results included in supplementary material. Summaries of analytical sample sizes are shown in Figure 1 (v4 - v2.1 subcohort in Supplemental Figure 1).

**Figure 1.**
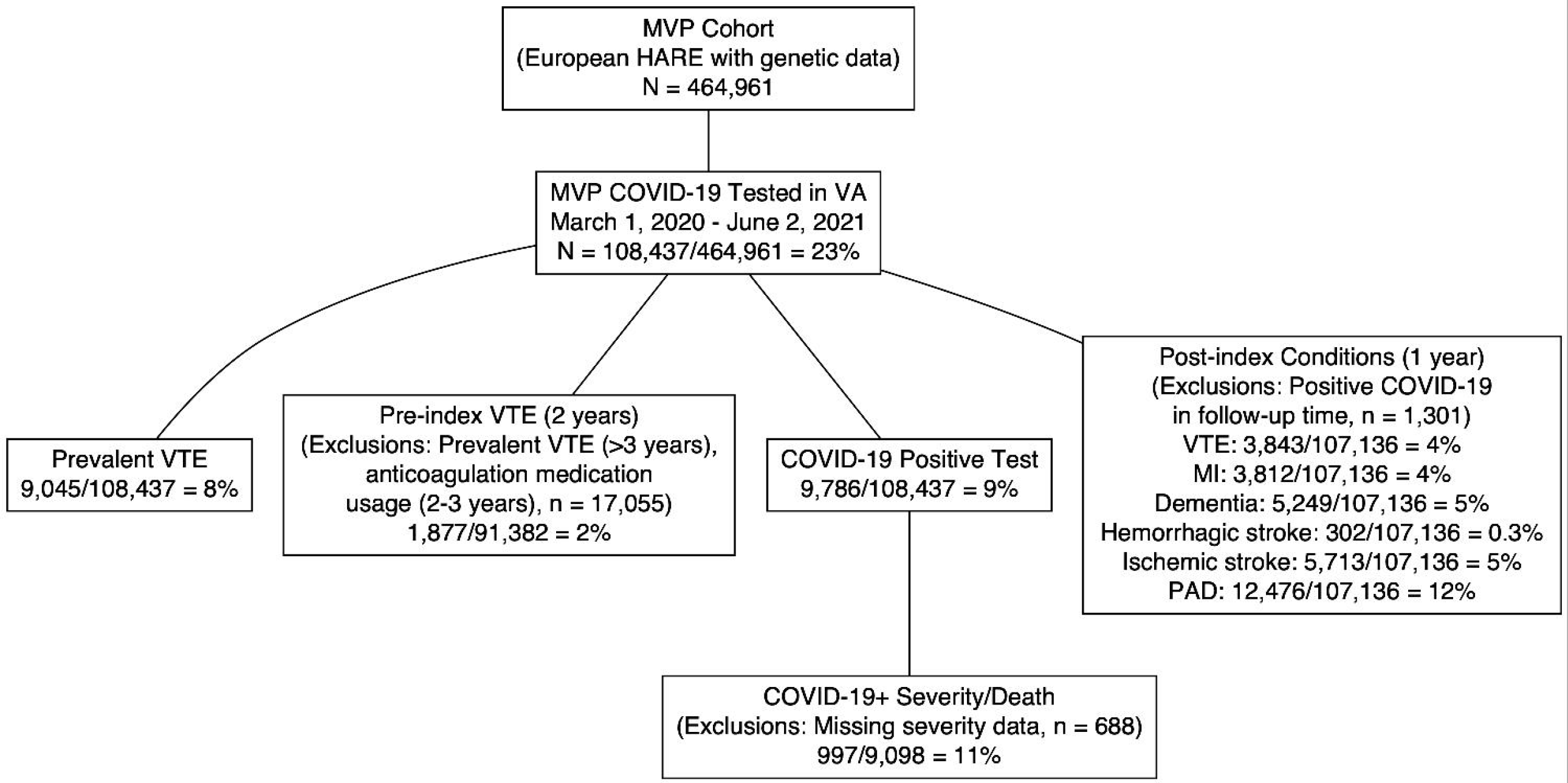
Study cohort flowchart of analytical sample sizes for each outcome.

### Definitions

The *index date* was defined as COVID-19 diagnosis date, i.e., specimen date; and for a hospitalized patient, the admission date up to 15 days prior to the COVID-19 diagnosis date. COVID-19 severity scale was derived from the WHO COVID-19 Disease Progression Scale^18^ as mild, moderate (hospitalization), severe (Intensive Care Unit-level care), or death within 30 days of index dates. Pre-index conditions within 2 years prior to the index dates were derived as described in Supplementary Methods and Supplemental Table 1. Post-index conditions were studied with up to 1 year follow-up after index date.VTE was defined in four periods: (1) prevalent: any time prior to the index date, (2) pre-index: VTE within the two years prior to index date, (3) post-index 60 days: within 60 days post index date, (4) post-index one year: within one year post-index date or date of last follow up if less than one year.

**Table 1.** Cohort demographics of COVID-19 tested European MVP participants, overall and stratified by COVID-19 test status as well as by prevalent VTE.

| Clinical and Demographic Variables | Overall | COVID-19 negative | COVID-19 positive | No prevalent VTE | Prevalent VTE |
| --- | --- | --- | --- | --- | --- |
| n | 108437 | 98651 | 9786 | 99392 | 9045 |
| Genetic data version v2.1 | 54850 (50.6) | 50032 (50.7) | 4818 (49.2) | 49870 (50.2) | 4980 ( 55.1) |
| **Demographics** |  |  |  |  |  |
| Prevalent VTE (%) | 9045 ( 8.3) | 8191 ( 8.3) | 854 ( 8.7) |  |  |
| BMI (kg/mi<sup>2</sup>) at the index date (mean (SD)) | 30.18 (6.43) | 30.10 (6.42) | 31.02 (6.52) | 30.11 (6.39) | 30.95 (6.86) |
| Age at the index date (mean (SD)) | 66.88 (12.74) | 67.00 (12.65) | 65.71 (13.58) | 66.59 (12.89) | 70.05 (10.47) |
| Sex = male (%) | 98397 (90.7) | 89459 (90.7) | 8938 (91.3) | 89946 (90.5) | 8451 ( 93.4) |
| **Genetics** |  |  |  |  |  |
| Polygenic Risk Score(VTE) (scaled with mean 0, SD=1 in overall population; mean(SD)) | 0.00 (1.00) | 0.00 (1.00) | 0.04 (0.99) | -0.03 (0.99) | 0.36 (1.09) |
| PRS(VTE) Quintiles (%) |  |  |  |  |  |
| ___q1 | 21688 (20.0) | 19871 (20.1) | 1817 (18.6) | 20558 (20.7) | 1130 ( 12.5) |
| ___q2 | 21689 (20.0) | 19772 (20.0) | 1917 (19.6) | 20290 (20.4) | 1399 ( 15.5) |
| ___q3 | 21687 (20.0) | 19734 (20.0) | 1953 (20.0) | 20050 (20.2) | 1637 ( 18.1) |
| ___q4 | 21685 (20.0) | 19604 (19.9) | 2081 (21.3) | 19663 (19.8) | 2022 ( 22.4) |
| ___q5 | 21688 (20.0) | 19670 (19.9) | 2018 (20.6) | 18831 (18.9) | 2857 ( 31.6) |
| PRS(VTE) highest 5th percentile (%) | 5423 ( 5.0) | 4918 ( 5.0) | 505 ( 5.2) | 4484 ( 4.5) | 939 ( 10.4) |
| F5 Leiden mutation carrier (%) | 5871 ( 5.4) | 5315 ( 5.4) | 556 ( 5.7) | 4834 ( 4.9) | 1037 ( 11.5) |
| F2 G20210A<br>mutation carrier (%) | 2968 ( 2.7) | 2701 ( 2.7) | 267 ( 2.7) | 2553 ( 2.6) | 415 ( 4.6) |
| **Pre-index conditions and medications in the 2 years prior to the index date** |  |  |  |  |  |
| Anticoagulation<br>medication usage<br>(%) | 19129<br>(17.6) | 17374<br>(17.6) | 1755 (17.9) | 13627 (13.7) | 5502 ( 60.8) |
| Venous<br>thromboembolism<br>(%) | 3786 ( 3.5) | 3427 ( 3.5) | 359 ( 3.7) | 0 ( 0.0) | 3786 ( 41.9) |
| Charlson<br>comorbidity index<br>(median [IQR]) | 2.00 [1.00,<br>4.00] | 2.00 [1.00,<br>4.00] | 2.00 [0.00,<br>4.00] | 2.00 [0.00,<br>4.00] | 3.00 [1.00, 5.00] |
| Hemorrhagic stroke<br>(%) | 299 ( 0.3) | 269 ( 0.3) | 30 ( 0.3) | 249 ( 0.3) | 50 ( 0.6) |
| Ischemic stroke (%) | 6549 ( 6.0) | 5958 ( 6.0) | 591 ( 6.0) | 5693 ( 5.7) | 856 ( 9.5) |
| Acute Myocardial<br>Infarction (%) | 3372 ( 3.1) | 3003 ( 3.0) | 369 ( 3.8) | 2853 ( 2.9) | 519 ( 5.7) |
| Peripheral artery<br>disease (%) | 19069<br>(17.6) | 17368<br>(17.6) | 1701 (17.4) | 16141 (16.2) | 2928 ( 32.4) |
| **Post-index conditions within 60 days after index date** |  |  |  |  |  |
| Hemorrhagic stroke<br>(%) | 1316 ( 1.2) | 1239 ( 1.3) | 77 ( 0.8) | 1183 ( 1.2) | 133 ( 1.5) |
| Ischemic stroke (%) | 1314 ( 1.3) | 1239 ( 1.4) | 75 ( 0.8) | 1180 ( 1.3) | 134 ( 1.7) |
| Acute Myocardial<br>Infarction (%) | 1769 ( 1.7) | 1565 ( 1.7) | 204 ( 2.2) | 1584 ( 1.7) | 185 ( 2.2) |
| Peripheral artery<br>disease (%) | 2553 ( 2.9) | 2392 ( 3.0) | 161 ( 2.0) | 2266 ( 2.7) | 287 ( 4.7) |
| Venous<br>thromboembolism<br>(%) | 1218 ( 1.2) | 1002 ( 1.1) | 216 ( 2.3) | 1005 ( 1.0) | 213 ( 4.1) |
| **Post-index conditions within 1 year after index date** |  |  |  |  |  |
| Venous<br>thromboembolism<br>(%) | 3886 ( 3.6) | 3429 ( 3.5) | 457 ( 4.7) | 1882 ( 1.9) | 2004 ( 22.2) |
| Acute Myocardial<br>Infarction (%) | 3838 ( 3.5) | 3454 ( 3.5) | 384 ( 3.9) | 3374 ( 3.4) | 464 ( 5.1) |
| Dementia (%) | 5293 ( 4.9) | 4660 ( 4.7) | 633 ( 6.5) | 4699 ( 4.7) | 594 ( 6.6) |
| Hemorrhagic stroke (%) | 302 ( 0.3) | 270 ( 0.3) | 32 ( 0.3) | 260 ( 0.3) | 42 ( 0.5) |
| Ischemic stroke (%) | 5738 ( 5.3) | 5299 ( 5.4) | 439 ( 4.5) | 5014 ( 5.0) | 724 ( 8.0) |
| Peripheral artery disease (%) | 12535 (11.6) | 11521 (11.7) | 1014 (10.4) | 10634 (10.7) | 1901 ( 21.0) |

Genotype data and polygenic risk score details are provided in Supplementary Methods. The VTE polygenic risk score (PRS) of 297 variants (Supplemental Table 3)^15^ was constructed to not include linkage disequilibrium-based regions containing the *F5* p.R506Q and *F2* G20210A variants.

### Statistical Methods

Demographic and clinical characteristics within each analytical cohort were summarized with means and standard deviations (SD) or medians and interquartile ranges (IQR) where appropriate.

The proportion of MVP participants tested for COVID-19 with 95% confidence intervals were estimated in the full MVP cohort as well as within subgroups defined by PRS(VTE) quintiles, FVL mutation status, and *F2* mutation status. Logistic regression models were fit to test the association of continuous PRS(VTE) with odds of testing, adjusting for age, age^2^, sex, and the first 5 ethnicity-specific principal components (PCs).

Firth logistic regression^19,20^ as implemented in the R package “brglm2” (version 0.7.1)^21^ was used to examine the association of prevalent and pre-index VTE, COVID-19 outcomes, and post-index conditions with PRS297, FVL carrier status, and *F2* mutation carrier status in COVID-19 tested participants. Firth logistic regression was used to reduce possible small sample bias in the logistic regression maximum likelihood coefficient estimates. Adjustment variables included age, age^2^, sex, anticoagulation medication usage (in all outcomes except VTE), COVID-19 positivity (in post-index condition analyses) and the first 5 ancestry PCs. The multiplicative interaction effects between PRS(VTE) and anticoagulation medication usage, PRS(VTE) and FVL carrier, PRS(VTE) and COVID-19 positivity (for post-index conditions) and FVL carrier and anticoagulation medication usage were also tested. When any of these interaction effects were significant at p<0.1, stratified models were fit within sub-cohorts defined by FVL carrier status, medication usage, or COVID-19 positivity.

The PRS(VTE) was studied as a centered (mean=0) continuous variable scaled to have standard deviation 1, so that the OR associated with PRS(VTE) effect was in terms of 1 SD increase in PRS(VTE). The PRS(VTE) was also studied as a categorical quintile variable, and a dichotomized variable comparing the individuals with the highest 5% of PRS(VTE) scores compared to the remaining cohort.

To evaluate possible non-linear associations of PRS(VTE) with outcomes, restricted cubic spline regression models for PRS(VTE) were fit for each outcome with the “rms” R package (version 5.1-4)^22^. Spline models were also fit separately by FVL status and anticoagulation medication status when interaction effects were suspected. Predicted relative odds ratio curves from spline models were fit with reference at scaled PRS(VTE) value of 0 representing the mean PRS(VTE) value of the cohort, with all other covariates set to the reference category or median value where appropriate.

All analyses were performed with R version 3.6.1^23^. Statistical significance for hypothesis testing was defined by a two-sided p-value of <0.05.

### Data Sharing Statement

Full GWAS summary statistics from published MVP investigations are available in dbGaP (https://www.ncbi.nlm.nih.gov/gap) under the MVP accession (phs001672). Individual-level genotype or clinical data cannot be shared outside of MVP.

## Results

### COVID-19 infection and outcome severity

Rates of COVID-19 testing (23.3%) did not differ significantly across PRS(VTE) quintiles, *F5* mutation status, and prothrombin mutation status, with less than a half a percentage point difference from the average testing rate between subgroups (23.2%-23.5%; Supplemental Table 4). PRS(VTE) was not significantly associated with COVID-19 testing in a logistic regression model adjusting for age, age-squared, sex, and PCs (OR=1.00, 95% CI [0.99, 1.01] for 1SD of PRS).

However, among participants tested for COVID-19, PRS(VTE) was significantly associated with positive COVID-19 test results. Of 108,487 participants tested for COVID-19, 9786 (9%) tested positive. The interaction of PRS(VTE) x FVL had p-value 0.052 (p = 0.029 in genetic release set v4-v2.1), and hence positive test models were stratified by FVL carrier status. In *F5* wild-type individuals, 8.8% (9225/10485) tested positive for COVID-19. PRS was associated with increased testing positive in *F5* WT with an adjusted OR of 1.05 (95%CI 1.02-1.07, p<0.001) for every 1-SD increase in PRS(VTE). Quintiles of PRS(VTE) were also significantly associated with COVID-19 status, with increased ORs compared to the lowest quintile (OR 1.06, 1.09, 1.16, 1.11 for q2-5 respectively, with p-values 0.10, 0.016, <0.001, 0.002) in WT individuals, after adjustment. In FVL individuals, 9.5% (556/5871) tested positive for COVID-19. Within these *F5* mutation carriers, PRS(VTE) was not associated with COVID-19 testing positive (OR 1SD 0.97, [0.91-1.94]). Restricted cubic spline analysis of PRS(VTE) within *F5* strata showed a protective effect of low polygenic risk score (scaled PRS(VTE) < 0) within wild type individuals and a more constant effect of higher PRS(VTE) (Figure 3A; Supplemental Figure 2A).

Neither FVL nor *F2 G20210A* was associated with the odds of testing positive for COVID-19 in univariate models or adjusted models. There was no interaction with the usage of outpatient anticoagulants and PRS(VTE).

Outpatient anticoagulation medication was associated with positive COVID-19 test within *F5* WT individuals, with a 7% increase in odds of positive COVID-19 test with medication usage within 2 years of testing (OR 1.07, [1.01-1.13], adjusting for PRS(VTE) continuous).

To examine COVID-19 outcome severity, 9098 COVID positive participants were included in the analysis, where 2656 (29.2%) were hospitalized, 997 (11%) were documented with a severe outcome or death. Outpatient anticoagulant medication usage within 2 years of positive test was associated with COVID-19 severity, with a univariate odds ratio of 1.75 [1.50-2.04], and adjusted odds ratio of 1.16 [0.99-1.36]. Genetic factors such as PRS(VTE), FVL, and F2 mutation were not associated with COVID-19 severity.

### Prevalent VTE

Of the 108,437 participants tested for COVID-19, 9045 (8.3%) had a prior diagnosis of VTE. These were considered prevalent VTE cases at index date. Outpatient anticoagulation medication usage (within 2 years prior to index date) was more common in prevalent VTE cases (60.8% vs 13.7% in participants without VTE), and VTE cases were more likely to be carriers of FVL (11.5% vs 4.9% in non-VTE participants) or *F2 G20210A* (4.6% vs 2.6%). The multiplicative interaction effect between PRS(VTE) (continuous) and FVL (FVL vs *F5* WT) was marginally significant (p=0.053) but not significant in the smaller cohort excluding v2.1 participants (p=0.5). Since one of our main interests was determining whether PRS(VTE) conferred additional VTE risk in participants with FVL, we also stratified the models by FVL status (Figure 2). PRS(VTE) was highly significantly associated with an increased risk of VTE after adjustment for basic confounders age, quadratic age, sex, and PCs. In *F5* WT individuals, PRS had an adjusted OR of 1.40 (95% CI [1.37-1.44]) for every 1SD increase, validating results from Klarin et al in genetic release v4, and 1.35 [1.31-1.40] in the validation set removing the training set v2.1. In FVL carriers, the OR for 1SD increase in PRS(VTE) was 1.33 (95% CI [1.37-1.44]) and 1.32 [1.22-1.43] in v4 and v4 - v2.1, respectively. An increased risk was also seen with increasing quintiles of PRS(VTE) (Figure 2) and in participants within the highest 5th percentile of PRS(VTE) (OR 1.74 [1.50-2.02] in v4-v2.1 in *F5* WT and 1.68 [1.36-2.07] in v4-v2.1 in FVL carriers; Supplemental Table 5). FVL was significantly associated with VTE diagnosis, with OR 1.84 (95% CI [1.66-2.04] in adjusted PRS(VTE)-FVL interaction model, at scaled PRS=0, Figure 2) and *F2 G20210A* had an adjusted OR of 1.75 [1.57-1.95] (adjusted for PRS(VTE) continuous, age, age-squared, sex, pc1-5).

**Figure 2.**
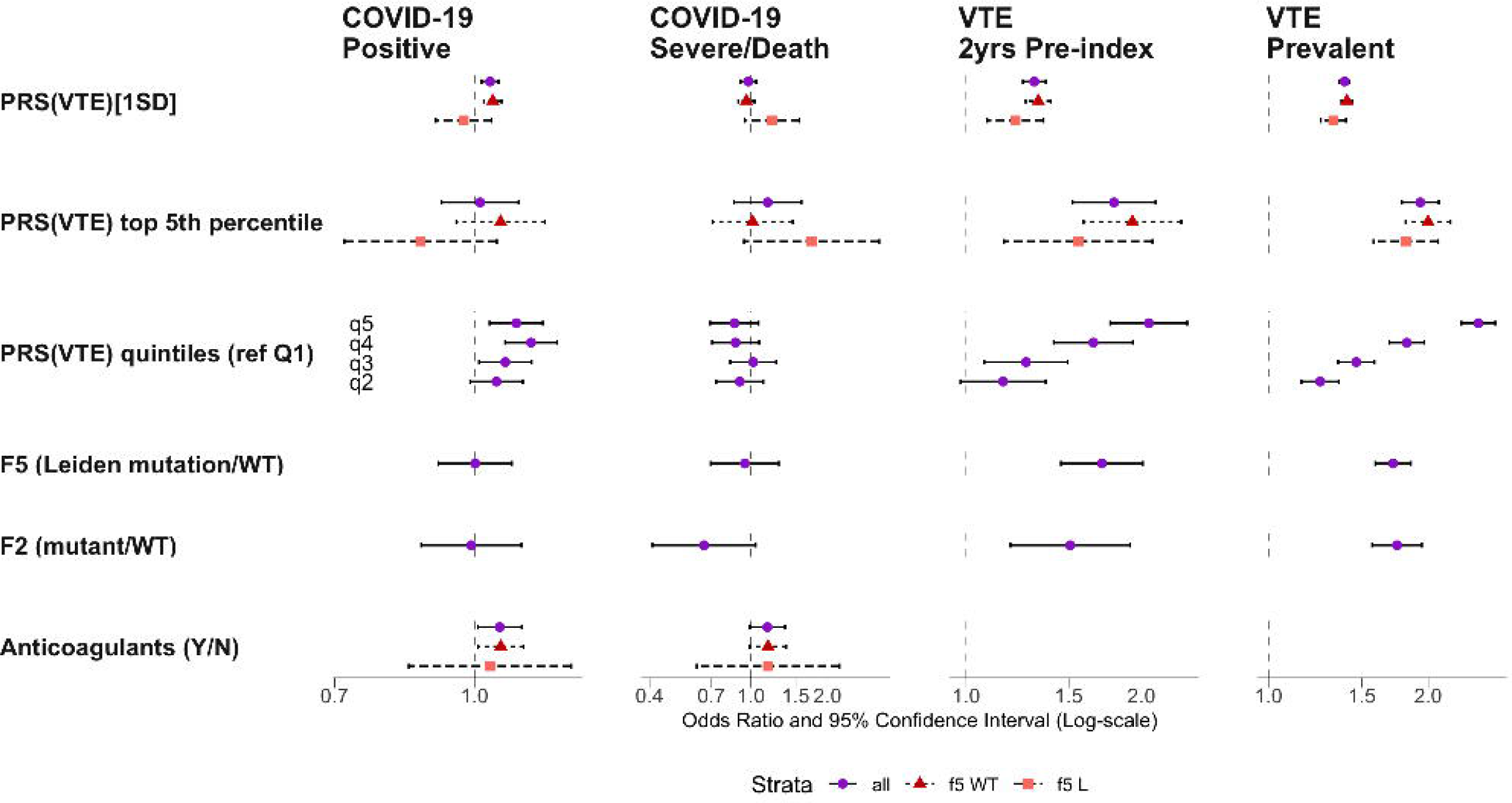
Forest plot of odds ratios and 95% confidence intervals. Odds ratios (ORs) shown for PRS(VTE) continuous (OR with respect to 1SD increase), PRS(VTE) quintiles, PRS(VTE) top 5% compared to bottom 95%, FVL carriers, F2 G20210A carriers, and outpatient anticoagulation medication usage (anticoagulants) for study outcomes: COVID-19 positive test result, COVID-19 severity (severe or death), pre-index history of VTE (within 2 years of index date), prevalent VTE. Strata denotes stratified models within FVL carriers versus F5 WT individuals, and all participants.

### Incident VTE in the pre-index period

In tested participants, 14,074 (13.0%) had a recent history of anticoagulation medication usage between 2- and 3-years pre-index date. These participants were removed from the analysis of pre-index VTE (0 to 2 years prior to index) along with prevalent VTE cases prior to 2 years pre-index date in order to study the impact of FVL and PRS(VTE) on the incidence of VTE in the two-year pre-index period (Figure 1). This resulted in a cohort of 91,382 tested participants with 1877 cases (2.0%) of VTE in the 2-year pre-index period. Pre-index VTE cases were very likely to take anticoagulation medication in that period (74.1% vs 5.1% in non-cases), and were also more likely to be carriers of FVL (10.6% vs 4.8% in non-VTE pre-index participants) or *F2 G20210A* (3.9% vs 2.6%). The rate of COVID-19 positive testing was similar between groups (9.7% in VTE cases vs 8.9% in non-cases). Results were similar to prevalent VTE analyses. In *F5* wild type individuals, PRS(VTE) had an adjusted OR of pre-index VTE of 1.33 (95% CI [1.27-1.40]) for every 1SD increase, and 1.31 [1.22-1.41] in the validation set removing the training set v2.1. In FVL carriers, the PRS(VTE) odds ratio for 1SD increase was 1.21 [1.09-1.36] and 1.244 [1.06-1.44] in v4 and v4-v2.1, respectively. Similar patterns were seen for quintiles and top 5th percentile (Figure 2, Supplementary Table 5). The results in v4-v2.1 validated the performance of this PRS(VTE) with regards to VTE development in EUR ancestry and exhibited additional contribution of PRS(VTE) in FVL carriers (Supplementary Table 5). FVL was significantly associated with VTE diagnosis, with OR 1.91 (95% CI [1.55-2.37] in adjusted PRS(VTE)-FVL interaction model, at scaled PRS=0, Figure 2) and *F2 G20210A* had an adjusted OR of 1.51 [1.19-1.91] (adjusted for PRS continuous, age, age-squared, sex, pc1-5).

To examine possible non-linear association of PRS(VTE) with prevalent and pre-index VTE, restricted cubic spline analysis of PRS(VTE) within *F5* strata was performed. These models showed a strong linear association of increasing PRS(VTE) with prevalent VTE within *F5* WT participants. In FVL carriers, a slight plateau effect is seen where the slope of log-odds ratio comparing high PRS(VTE) to PRS=0 does not increase as steadily in FVL carriers as seen in *F5* WT individuals (Figure 3B; Supplemental Figure 2B). Pre-index VTE associations are similar to prevalent VTE associations in *F5* WT individuals.However, within FVL carriers, increased association of pre-index VTE was significant only in higher PRS(VTE) scores greater than approximately PRS(VTE)=2 compared to PRS(VTE)=0 (Figure 3C; Supplemental Figure 2C).

**Figure 3.**
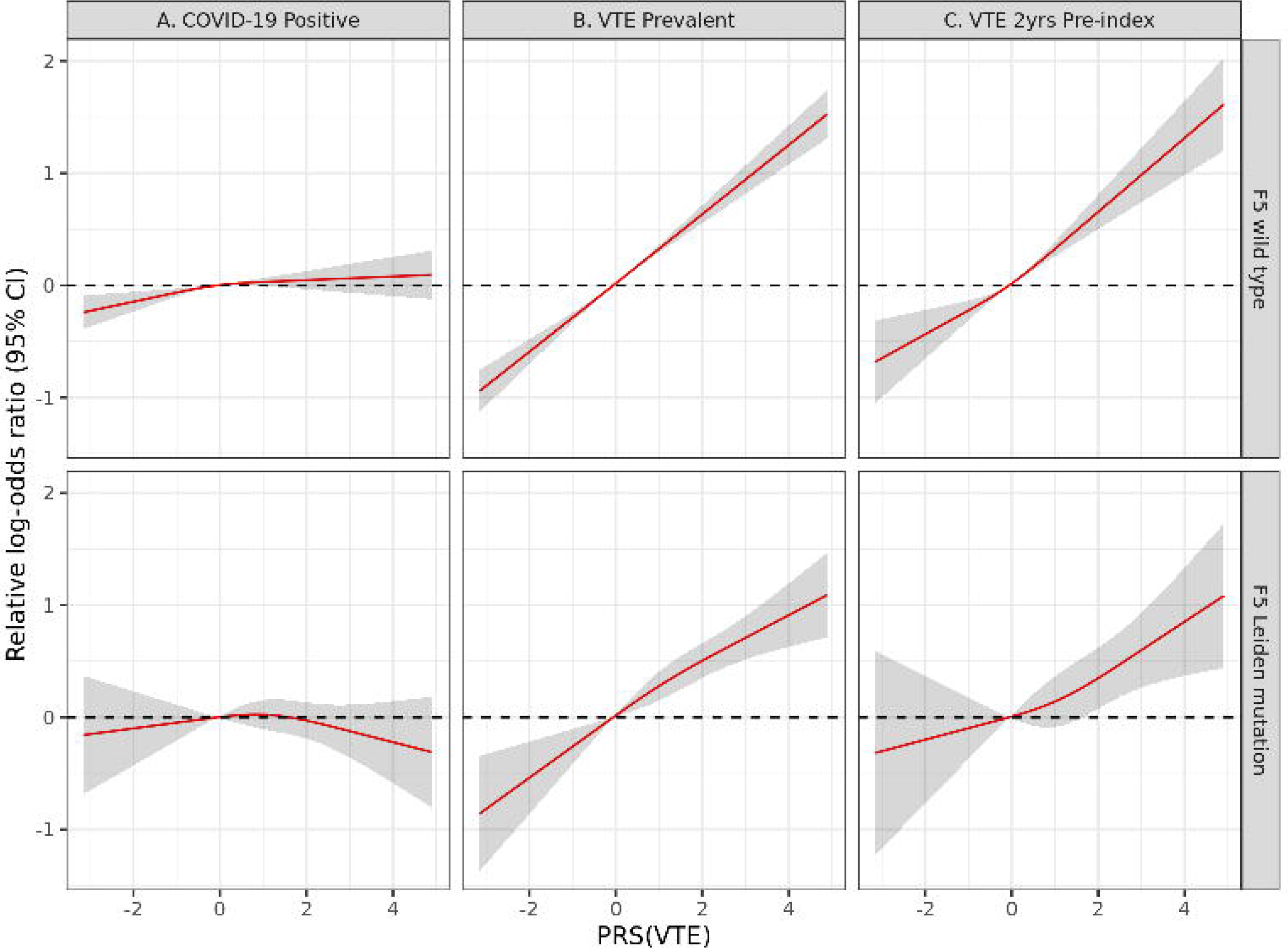
Predicted relative log-odds ratio curves from spline regression analyses. Log-odds of COVID-19 positive test (A) and prevalent VTE diagnosis (B), and pre-index incident VTE diagnosis within two years prior to testing (C). ORs for PRS(VTE) are compared to scaled PRS(VTE) value of 0 (mean PRS), from cubic spline models stratified by FVL carrier status. ORs evaluated at median or reference group of other model covariates. Shaded area represents the 95% confidence interval.

### Post-COVID-19 conditions

We examined the development of incident diagnoses of peripheral artery disease (PAD), VTE, acute myocardial infarction (AMI), Ischemic stroke, hemorrhagic stroke, or dementia up to 1 year following the post-index date. In the 107,136 eligible, tested individuals, 12476 (11.6%) were diagnosed with post-index PAD, 3843 (3.6%) with VTE, 3812 (3.6%) with AMI, 5713 (5.3%) with ischemic stroke, 302(0.3%) with hemorrhagic stroke, and 5249 (4.9%) with dementia.

FVL carrier status was associated with decreased risk of hemorrhagic stroke (univariate OR 0.50, [0.25-1.00]; adjusted OR 0.47 [0.24-0.94]), and an increased risk of PAD (univariate OR 1.14 [1.06-1.24], adjusted OR 1.08 [1.00-1.18]) in the whole cohort after adjusting for COVID-19 status. PRS(VTE) was also significantly associated with an increased risk of PAD, with an adjusted OR of 1.02 [1.00-1.04].

COVID-19 positivity was associated with increased risk of acute MI (adjusted OR 1.16 [1.04-1.29]), VTE (OR 1.37 [1.24-1.52]), and dementia (OR 1.49 [1.36-1.62]). However, testing positive for COVID-19 was associated with a decreased risk of ischemic stroke (0.85, [0.77-0.94]), and PAD (0.90, [0.84-0.96]).

There was suggestive evidence of effect modification by COVID-19 status with respect to the PRS association of post-index PAD, with a PRS-COVID+ interaction p-value of 0.035 and an OR in a stratified model with OR of 1.09 [1.02-1.17] in COVID-19 positive participants, and 1.01 [0.99-1.03] in COVID-19 negative patients. The PRS-COVID-19+ interaction effect was significant for post-index VTE as well, with p-value 0.006. Stratified by COVID-19 positivity, the COVID-19 negative participants had a higher risk of post-index VTE, with OR 1.31 [1.25-1.38] in COVID- and OR 1.18 [1.03-1.35] in COVID-19+ individuals.

Anticoagulation medication usage 2 years prior to index date was associated with increased risk of all post-index conditions examined in both univariate and adjusted analyses.

## Discussion

We found that increased polygenic propensity for VTE is associated with increased COVID-19 testing positive rates, suggesting a role for coagulation in the initial phase of COVID-19 infection. There was an interaction between FVL and PRS(VTE) as PRS(VTE) did not confer risk among FVL carriers but only in *F5* WT individuals. Neither FVL nor *F2* G20210A was associated with being tested or having a positive testing result. Though PRS(VTE) and FVL both contributed to increased risk of VTE, the PRS(VTE) association with testing positive was not observed in FVL individuals, suggesting that unique process(es) represented by PRS(VTE) but not F5 were involved. Alternatively, factor V Leiden may be epistatic over PRS(VTE). The parallel between increased risks of VTE and COVID-19 testing positive conferred by PRS, modified by the *F5* status, supported involvement of polygenic risk towards VTE in the COVID-19 infection.

The significance of the study is it provides biological insights into the earliest phases of COVID-19 infection. Further molecular characterization of the interaction with clotting may yield new therapeutic and/or prognostic options.

In critically ill patients, therapeutic dose anticoagulation with heparin did not improve clinical outcomes and was associated with an excess risk of major bleeding events when compared with routine prophylactic heparin^24^. Trials in moderately ill patients with COVID-19 have reached mixed results but therapeutic dose anticoagulation was associated with excess bleeding^25–28^. The HEP-COVID trial showed that therapeutic-dose LMWH reduced major thromboembolism and death compared with institutional standard heparin thromboprophylaxis among inpatients with COVID-19 with very elevated D-dimer levels. The treatment effect was not seen in ICU patients^29^. The lack of uniform impact of the intensity of the anti-coagulation on the COVID-19 outcome is consistent with our observations that there was no association of the outcome severity with PRS(VTE), FVL or *F2* G20210A. Whether low dose anticoagulation for example would have any role in the prevention and/or post-exposure prophylaxis for COVID-19 infection for patients with high PRS(VTE) remains to be evaluated.

Though prior anticoagulant usage was associated with an increased risk of being tested positive. Whether prior anticoagulant treatment was associated with getting tested for COVID-19 was not known, making interpretation of these results on testing positive more difficult.

Prior outpatient anticoagulant usage was associated with worse clinical outcomes and the development of more post-index clinical events. This could be related to associated medical conditions that necessitated the usage of oral anticoagulants.

The impact of these monogenic or polygenic thrombotic risks on the development of specific clinical events after COVID-19 infection was explored in this study. FVL was associated with fewer incidents of hemorrhagic stroke in all the tested individuals as has been previously reported^30^. Both FVL and PRS(VTE) were associated with an increased incident diagnosis of PAD, replicating an earlier finding of a role of FVL in PAD in the MVP^31^. The genetic interaction between PRS(VTE) and COVID-19 positivity with regards to PAD and VTE in the prospective follow up post-index merits further study.Additionally, our results suggested clinical attention be paid to PAD among COVID-19 positive patients with FVL or high PRS(VTE). The association of COVID-19 positive test with an increased risk of dementia in the MVP is consistent with an earlier report^32^. Caution is needed in the interpretation of the decreased incidence of hemorrhagic stroke or PAD in COVID-19 positive individuals and these results are considered hypothesis generating.

Additionally, PRS(VTE) can further risk stratify patient populations regarding their VTE risk among FVL carriers among prevalent or pre-index patient populations. PRS(VTE) may be clinically useful for us to distinguish FVL carriers that are on the higher versus lower end of the risk spectrum for VTE events. Individuals in the top quintile of PRS(VTE) had a twofold increase in risk compared to individuals in the lowest quintile (OR 1.99 [1.68-2.36]). Considering 6% of the EUR populations are FVL carriers this is clinically significant and merits further investigation.

Within FVL carriers, increased association of pre-index VTE was significant only in higher PRS(VTE) scores greater than approximately PRS(VTE)=2 compared to PRS(VTE)=0 in restricted spline models (Figure 3C). This may be due to smaller sample sizes underrepresenting low PRS(VTE) scores in FVL carriers, or may support the use of quintiles or top 5th percentile to better model the effect of a non-linear PRS(VTE) association in FVL participants.

Study limitations are that only European ancestry participants were studied as PRS(VTE) was developed within cohorts of European ancestry. Only patients that were COVID-19 tested were included in this study. The last patients that were tested in the study cohort were on June 1 2021 so vaccination status might play a role in the study. The serotype of viruses involved was not known. Treatment modalities had evolved during the study period. Diagnosis of clinical events in the immediate post index period might be affected by potential underutilization of imaging modalities required to make certain diagnoses. The latter was expected to dissipate however after a recommended short period of quarantine and with longer follow up. An important consideration in the issue of testing positive rates was exposure. It is unlikely that PRS(VTE) was associated with differential environmental exposure however.

Strengths include a large sample size and geographically diverse population evaluated, the prospective nature of this study to examine testing (positive) rates and clinical sequelae of COVID-19 infection, and the use of genetic stratification to investigate post COVID-19 events.

In summary, PRS(VTE) was associated with COVID-19 positive testing rates for EUR in this large prospective cohort from the MVP. The linkage of COVID-19 infection with thromboembolism processes is novel and may reveal mechanisms of early steps of viral infection and/or treatment and prevention strategies.

## Acknowledgements

S.-W. L was an ASH fellow scholar.

This research is based on data from the Million Veteran Program, Office of Research and Development, Veterans Health Administration, and was supported by award # MVP035. This publication does not represent the views of the Department of Veteran Affairs or the United States Government.

## Authorship

Contribution: Conception and Design, J.M., J.E.H., L.G., J.J., E.S.W., W.W., A.S., G.A.P., R.P., M.A., K.C., H.G., A.V., Y.H., J.B.M., K.C., R.A.B., B.R.G., S.P., E.G., N.R., K.E.L., J.A.L., S.M.Z., P.N., C.J.M., J.J.Z., D.N.J., C.J.D., J.E.M., P.D.R., Y.V.S., M.F., J.G., J.M.P., A.H., R.D.H., R.K.M., S.D., Q.S.W., K.P.L., P.W.W., P.S.T., C.J.O., J.M.G., R.L.H., S.K.I., S.-W.L.; Development of Methodology, J.M., S.K.I., S.-W.L; Acquisition of Data, J.M., L.G., S.K.I., S.-W.L; Analysis and Interpretation of Data, J.M., J.E.H., L.G., S.K.I., S.-W.L; Writing of Manuscript, J.M., J.E.H., L.G., J.J., E.S.W., W.W., A.S., G.A.P., R.P., M.A., K.C., H.G., A.V., Y.H., J.B.M., K.C., R.A.B., B.R.G., S.P., E.G., N.R., K.E.L., J.A.L., S.M.Z., P.N., C.J.M., J.J.Z., D.N.J., C.J.D., J.E.M., P.D.R., Y.V.S., M.F., J.G., J.M.P., A.H., R.D.H., R.K.M., S.D., Q.S.W., K.P.L., P.W.W., P.S.T., C.J.O., J.M.G., R.L.H., S.K.I., S.-W.L.; Study Supervision, R.L.H., S.K.I., S.-W.L.

