## Supplemental Materials for "Polygenic predisposition to venous thromboembolism is associated with increased COVID-19 positive testing rates"

### Supplemental Methods

*Study Cohort*

The MVP cohort has been described elsewhere[1]. Briefly, United States veterans were recruited from 63 participating Department of Veterans Affairs (VA) medical facilities starting in 2011. Participants provided consent to access their electronic health records for research, provided blood samples for genotyping, and were given surveys to obtain demographic and lifestyle information. The MVP protocol was approved by the VA Central Institutional Review Board in accordance with principles outlined in the Declaration of Helsinki.

This study utilized the COVID-19 Shared Data Resource (SDR) which was created by the VA for COVID-19 studies in April-August 2020. The effort was led by the VA Informatics and Computing Infrastructure (VINCI). Metadata on documentations of conditions, laboratory measures, medications, and procedures pertaining to the COVID-19 pandemic and dissemination of information regarding SDR were provided by the VA Phenomics Library, Centralized Interactive Phenomics Resource (CIPHER), to users across the VA healthcare systems[2]. De-identification and security was implemented by the VA National Surveillance Team. Curation of data was conducted by an experienced VA research team in order to ensure consistent applications of dates across the EHR tables, as well as uniform definitions of events preceding and subsequent to COVID-19, in an effort to characterize the trajectory of COVID-19 and other diseases and conditions. VA-wide efforts describing these data are provided elsewhere. Structured data obtained via CPT and ICD9 or ICD10 codes, deposited in the VA Corporate Data Warehouse (CDW),was further enriched with rule-based unstructured events recorded in patient notes via natural language processing (NLP), as was done previously for other projects[3]. The purpose of NLP-boosting was to fill gaps in knowledge about the severity of the disease, as well as extract specific dates when procedures (e.g. intubation and extubation) were performed. These curated data were then provisioned in the Million Veteran Program (MVP) study mart for COVID-19 in VINCI ensuring data security behind the VA firewall.

European ancestry was determined from genotype data and self-identified race/ethnicity using the Harmonized Ancestry and Race/Ethnicity (HARE)[4] classification algorithm.

Participants who were tested for COVID-19 within the VA healthcare system between March 1, 2020 and June 2, 2021 were included (Patient table dated 06-15-2021), with follow-up data studied until September 2, 2021. Severity of COVID-19 was measured with a maximum follow up date of September 22, 2021. COVID-19 cases were identified by the VA COVID National Surveillance Tool (NST) algorithm [5]. They were accessed in the Patient table from the Shared Data Resource (SDR). All analyses were conducted using the release 4 of the genetic data from the MVP program; sensitivity analyses were conducted in the subset of participants who were not included in the genetic release v2.1 used for the training of the PRS(VTE)[6] with results included in supplemental material. Summaries of the analytical sample size are shown in Figure 1 (genetic v4, Supplemental Figure 1 for v4 - v2.1 subcohort).

*COVID-19 index date and severity definition*

For this work we only studied MVP participants who received their Polymerase chain reaction (PCR)-based COVID-19 testing within VA systems. MVP individuals that did not have clinical follow up information or were missing genotyping information were excluded.

COVID-19 severity scale was derived from the WHO COVID-19 Disease Progression Scale[7] as mild, moderate (hospitalization), severe (Intensive Care Unit-level care), or death within 30 days of index dates. Severe outcomes include use of any of the following: intubation, pressor, hemodialysis, non-invasive positive pressure ventilation or dialysis. All data and variables were assessed centrally by the MVP data core's Shared Data Repository (SDR).

*Pre and post-index conditions*

Pre-index conditions were derived using natural language processing (NLP)-boosted analysis of unstructured notes, ICD and Current Procedural Terminology (CPT) codes, and medications taken 2 years prior to the index dates. Post-index conditions were derived using ICD and CPT codes, and medications up to 365 days after the index date.

The list of ICD codes used to define/pull the pre- and post-index diagnosis of ischemic stroke, hemorrhagic stroke, acute myocardial infarction, venous-thromboembolism (VTE), peripheral arterial disease, and dementia are presented in Supplemental Table 1.

The study of post-index conditions was conducted in all COVID tested participants with the exclusion of participants who tested negative at the last testing date (June 2, 2021) but had a positive test by the last follow up date (September 2, 2021).

*Anticoagulation Medication*

The list of outpatient anticoagulants studied in this work is provided in Supplemental Table 2. Only outpatient usage was studied.

*Genotype data*

The single nucleotide polymorphism (SNP) data in the MVP cohort was generated using a custom Thermo Fisher Axiom genotyping platform called MVP 1.0. The quality control steps and genotyping imputation using 1000 Genomes cosmopolitan reference panel for the MVP cohort have been previously reported[8]. Harmonized ancestry, race and ethnicity (HARE) is a composite variable derived from a combination of self-reported survey information and genetically-derived ancestry [4] and was employed as a proxy for population stratification and global ancestry. For *F2* and *F5*, directly genotyped allele information (rs6025 and rs1799963, respectively) was used. For the calculation of the Polygenic Risk Score (PRS) discussed below, either directly genotyped or imputed allele information (with r2>0.6) was used. Altogether 287 (out of 297) SNPs were used (Supplemental Table 3).

*Polygenic Risk Score*

Genotype data and polygenic risk score details are provided in Supplemental Methods. The VTE polygenic risk score (PRS) of 297 variants (Supplemental Table 3) was originally constructed by Klarin et. al [6]. The PRS(VTE) was calculated as the sum of the number of risk alleles or allele dosage multiplied by the log-odds ratio estimated from the meta-analysis of European-descent Million Veteran Program (genetic release 2.1) and the European-descent UK Biobank summary statistics. The PRS(VTE) formula is

PRS297 = $\beta_1 x_1 + \beta_2 x_2 + \ldots + \beta_{297} x_{297}$

Where $x_j$ is the number of risk alleles (0, 1, 2) for SNP $k$, and $\beta_k$ are the estimated log-odds ratios from the study by Klarin and colleagues [6]. The PRS(VTE) was constructed to not include linkage disequilibrium-based regions containing the *F5* p.R506Q and *F2* G20210A variants. In this study, the PRS(VTE) was calculated for European MVP participants with release 4 genetic data available, using genotyped or imputed allele information from 287 SNPs (with $R^2$ in HARE EUR > 0.6). To avoid possible training set overfitting bias, analyses were additionally performed in the subset of the release 4 cohort excluding participants in release 2.1 studied in the MVP analysis [6] with results provided in supplemental materials.

### Supplemental Tables

**Supplemental Table 1. The ICD codes used for data pull of post-index diagnoses in this study.**

| Phenotype | phenotype description | vocabulary | code | Code description |
|---|---|---|---|---|
| ActMI | Acute Myocardial Infarction | ICD10 | I21.01 | ST elevation (STEMI) myocardial infarction involving left main coronary artery |
| ActMI | Acute Myocardial Infarction | ICD10 | I21.02 | ST elevation (STEMI) myocardial infarction involving left anterior descending coronary artery |
| ActMI | Acute Myocardial Infarction | ICD10 | I21.09 | ST elevation (STEMI) myocardial infarction involving other coronary artery of anterior wall |
| ActMI | Acute Myocardial Infarction | ICD10 | I21.11 | ST elevation (STEMI) myocardial infarction involving right coronary artery |
| ActMI | Acute Myocardial Infarction | ICD10 | I21.19 | ST elevation (STEMI) myocardial infarction involving other coronary artery of inferior wall |
| ActMI | Acute Myocardial Infarction | ICD10 | I21.21 | ST elevation (STEMI) myocardial infarction involving left circumflex coronary artery |
| ActMI | Acute Myocardial Infarction | ICD10 | I21.29 | ST elevation (STEMI) myocardial infarction involving other sites |
| ActMI | Acute Myocardial Infarction | ICD10 | I21.3 | ST elevation (STEMI) myocardial infarction of unspecified site |

| Phenotype | phenotype description | vocabulary | code | Code description |
|---|---|---|---|---|
| ActMI | Acute Myocardial Infarction | ICD10 | I21.4 | Non-ST elevation (NSTEMI) myocardial infarction |
| ActMI | Acute Myocardial Infarction | ICD10 | I21.9 | Acute myocardial infarction, unspecified |
| ActMI | Acute Myocardial Infarction | ICD10 | I21.A1 | Myocardial infarction type 2 |
| ActMI | Acute Myocardial Infarction | ICD10 | I21.A9 | Other myocardial infarction type |
| ActMI | Acute Myocardial Infarction | ICD9 | 410 | ACUTE MYOCARDIAL INFARCTION OF ANTEROLATERAL WALL |
| ActMI | Acute Myocardial Infarction | ICD9 | 410 | ACUTE MYOCARDIAL INFARCTION, OF ANTEROLATERAL WALL, EPISODE OF CARE, UNSPECIFIED |
| ActMI | Acute Myocardial Infarction | ICD9 | 410.01 | ACUTE MYOCARDIAL INFARCTION, OF ANTEROLATERAL WALL, INITIAL EPISODE OF CARE |
| ActMI | Acute Myocardial Infarction | ICD9 | 410.02 | ACUTE MYOCRADIAL INFARCTION, OF ANTEROLATERAL WALL, SUBSEQUENT EPISODE OF CARE |
| ActMI | Acute Myocardial Infarction | ICD9 | 410.1 | ACUTE MYOCARDIAL INFARCTION OF OTHER ANTERIOR WALL |
| ActMI | Acute Myocardial Infarction | ICD9 | 410.1 | ACUTE MYOCARDIAL INFARCTION, OF OTHER ANTERIOR WALL, SUBSEQUENT EPISODE OF CARE UNSPECIFIED |

| Phenotype | phenotype description | vocabulary | code | Code description |
|---|---|---|---|---|
| ActMI | Acute Myocardial Infarction | ICD9 | 410.11 | ACUTE MYOCARDIAL INFARCTION, OF OTHER ANTERIOR WALL, INITIAL EPISODE OF CARE |
| ActMI | Acute Myocardial Infarction | ICD9 | 410.12 | ACUTE MYOCARDIAL INFARCTION, OF OTHER ANTERIOR WALL, SUBSEQUENT EPISODE OF CARE |
| ActMI | Acute Myocardial Infarction | ICD9 | 410.2 | ACUTE MYOCARDIAL INFARCTION OF INFEROLATERAL WALL |
| ActMI | Acute Myocardial Infarction | ICD9 | 410.2 | ACUTE MYOCARDIAL INFARCTION, OF INFEROLATERAL WALL, EPISODE OF CARE UNSPECIFIED |
| ActMI | Acute Myocardial Infarction | ICD9 | 410.21 | ACUTE MYOCARDIAL INFARCTION, OF INFEROLATERAL WALL, INITIAL EPISODE OF CARE |
| ActMI | Acute Myocardial Infarction | ICD9 | 410.22 | ACUTE MYOCARDIAL INFARCTION, OF INFEROLATAERAL WALL, SUBSEQUENT EPISODE OF CARE |
| ActMI | Acute Myocardial Infarction | ICD9 | 410.3 | ACUTE MYOCARDIAL INFARCTION OF INFEROPOSTERIOR WALL |
| ActMI | Acute Myocardial Infarction | ICD9 | 410.3 | ACUTE MYOCARDIAL INFARCTION, OF INFEROPOSTERIOR WALL, EPISODE OF CARE UNSPECIFIED |
| ActMI | Acute Myocardial Infarction | ICD9 | 410.31 | ACUTE MYOCARDIAL INFARCTION, OF INFEROPOSTERIOR WALL, INITIAL EPISODE OF CARE |
| ActMI | Acute Myocardial Infarction | ICD9 | 410.32 | ACUTE MYOCARDIAL INFARCTION, OF INFEROPOSTERIOR WALL, SUBSEQUENT EPISODE OF CARE |

| Phenotype | phenotype description | vocabulary | code | Code description |
|---|---|---|---|---|
| ActMI | Acute Myocardial Infarction | ICD9 | 410.4 | ACUTE MYOCARDIAL INFARCTION OF OTHER INFERIOR WALL |
| ActMI | Acute Myocardial Infarction | ICD9 | 410.4 | ACUTE MYOCARDIAL INFARCTION, OF OTHER INFERIOR WALL, EPISODE OF CARE UNSPECIFIED |
| ActMI | Acute Myocardial Infarction | ICD9 | 410.41 | ACUTE MYOCARDIAL INFARCTION, OF OTHER INFERIOR WALL, INITIAL EPISODE OF CARE |
| ActMI | Acute Myocardial Infarction | ICD9 | 410.42 | ACUTE MYOCARDIAL INFARCTION, OF OTHER INFERIOR WALL, SUBSEQUENT EPISODE OF CARE |
| ActMI | Acute Myocardial Infarction | ICD9 | 410.5 | ACUTE MYOCARDIAL INFARCTION OF OTHER LATERAL WALL |
| ActMI | Acute Myocardial Infarction | ICD9 | 410.5 | ACUTE MYOCARDIAL INFARCTION, OF OTHER LATERAL WALL, EPISODE OF CARE UNSPECIFIED |
| ActMI | Acute Myocardial Infarction | ICD9 | 410.51 | ACUTE MYOCARDIAL INFARCTION, OF OTHER LATERAL WALL, INITIAL EPISODE OF CARE |
| ActMI | Acute Myocardial Infarction | ICD9 | 410.52 | ACUTE MYOCARDIAL INFARCTION, OF OTHER LATERAL WALL, SUBSEQUENT EPISODE OF CARE |
| ActMI | Acute Myocardial Infarction | ICD9 | 410.6 | TRUE POSTERIOR WALL INFARCTION |
| ActMI | Acute Myocardial Infarction | ICD9 | 410.6 | ACUTE MYOCARDIAL INFARCTION, TRUE POSTERIOR WALL INFARCTION, EPISODE OF CARE UNSPECIFIED |

| Phenotype | phenotype description | vocabulary | code | Code description |
|---|---|---|---|---|
| ActMI | Acute Myocardial Infarction | ICD9 | 410.61 | ACUTE MYOCARDIAL INFARCTION, TRUE POSTERIOR WALL INFARCTION, INITIAL EPISODE OF CARE |
| ActMI | Acute Myocardial Infarction | ICD9 | 410.62 | ACUTE MYOCARDIAL INFARCTION, TRUE POSTERIOR WALL INFARCTION, SUBSEQUENT EPISODE OF CARE |
| ActMI | Acute Myocardial Infarction | ICD9 | 410.7 | SUBENDOCARDIAL INFARCTION |
| ActMI | Acute Myocardial Infarction | ICD9 | 410.7 | ACUTE MYOCARDIAL INFARCTION, SUBENDOCARDIAL INFARCTION, EPISODE OF CARE UNSPECIFIED |
| ActMI | Acute Myocardial Infarction | ICD9 | 410.71 | ACUTE MYOCARDIAL INFARCTION, SUBENDOCARDIAL INFARCTION, INITIAL EPISODE OF CARE |
| ActMI | Acute Myocardial Infarction | ICD9 | 410.72 | ACUTE MYOCARDIAL INFARCTION, SUBENDOCARDIAL INFARCTION, SUBSEQUENT EPISODE OF CARE |
| ActMI | Acute Myocardial Infarction | ICD9 | 410.8 | ACUTE MYOCARDIAL INFARCTION OF OTHER SPECIFIED SITES |
| ActMI | Acute Myocardial Infarction | ICD9 | 410.8 | ACUTE MYOCARDIAL INFARCTION, OF OTHER SPECIFIED SITES, EPISODE OF CARE UNSPECIFIED |
| ActMI | Acute Myocardial Infarction | ICD9 | 410.81 | ACUTE MYOCARDIAL INFARCTION, OF OTHER SPECIFIED SITES, INITIAL EPISODE OF CARE |
| ActMI | Acute Myocardial Infarction | ICD9 | 410.82 | ACUTE MYOCARDIAL INFARCTION, OF OTHER SPECIFIED SITES, SUBSEQUENT EPISODE OF CARE |

| Phenotype | phenotype description | vocabulary | code | Code description |
|---|---|---|---|---|
| ActMI | Acute Myocardial Infarction | ICD9 | 410.9 | ACUTE MYOCARDIAL INFARCTION OF UNSPECIFIED SITE |
| ActMI | Acute Myocardial Infarction | ICD9 | 410.9 | ACUTE MYOCARDIAL INFARCTION, UNSPECIFIED SITE, EPISODE OF CARE UNSPECIFIED |
| ActMI | Acute Myocardial Infarction | ICD9 | 410.91 | ACUTE MYOCARDIAL INFARCTION, UNSPECIFIED SITE, INITIAL EPISODE OF CARE |
| ActMI | Acute Myocardial Infarction | ICD9 | 410.92 | ACUTE MYOCARDIAL INFARCTION, UNSPECIFIED SITE, SUBSEQUENT EPISODE OF CARE |
| Alz | Alzheimers disease | ICD10 | G30.0 | Alzheimer's disease with early onset |
| Alz | Alzheimers disease | ICD10 | G30.1 | Alzheimer's disease with late onset |
| Alz | Alzheimers disease | ICD10 | G30.8 | Other Alzheimer's disease |
| Alz | Alzheimers disease | ICD10 | G30.9 | Alzheimer's disease, unspecified |
| Alz | Alzheimers disease | ICD9 | 331 | ALZHEIMER'S DISEASE |
| CTE | chronic traumatic encephalopathy | ICD10 | F07.81 | Postconcussional syndrome |

| Phenotype | phenotype description | vocabulary | code | Code description |
|---|---|---|---|---|
| CTE | chronic traumatic encephalopathy | ICD9 | 310.2 | POSTCONCUSSION SYNDROME |
| FrontoDem | Frontotemporal dementia | ICD10 | G31.09 | Other frontotemporal dementia |
| FrontoDem | Frontotemporal dementia | ICD9 | 331.1 | PICK'S DISEASE |
| FrontoDem | Frontotemporal dementia | ICD9 | 331.11 | PICK'S DISEASE |
| FrontoDem | Frontotemporal dementia | ICD9 | 331.19 | OTHER FRONTOTEMPORAL DEMENTIA |
| HemStroke | hemorrhagic stroke | ICD10 | I60.00 | Nontraumatic subarachnoid hemorrhage from unspecified carotid siphon and bifurcation |
| HemStroke | hemorrhagic stroke | ICD10 | I60.01 | Nontraumatic subarachnoid hemorrhage from right carotid siphon and bifurcation |
| HemStroke | hemorrhagic stroke | ICD10 | I60.02 | Nontraumatic subarachnoid hemorrhage from left carotid siphon and bifurcation |
| HemStroke | hemorrhagic stroke | ICD10 | I60.10 | Nontraumatic subarachnoid hemorrhage from unspecified middle cerebral artery |
| HemStroke | hemorrhagic stroke | ICD10 | I60.11 | Nontraumatic subarachnoid hemorrhage from right middle cerebral artery |

| **Phenotype** | **phenotype description** | **vocabulary** | **code** | **Code description** |
|---|---|---|---|---|
| HemStroke | hemorrhagic stroke | ICD10 | I60.12 | Nontraumatic subarachnoid hemorrhage from left middle cerebral artery |
| HemStroke | hemorrhagic stroke | ICD10 | I60.2 | Nontraumatic subarachnoid hemorrhage from anterior communicating artery |
| HemStroke | hemorrhagic stroke | ICD10 | I60.20 | Nontraumatic Subarachnoid Hemorrhage from unspecified Anterior Communicating Artery |
| HemStroke | hemorrhagic stroke | ICD10 | I60.21 | Nontraumatic Subarachnoid Hemorrhage from right Anterior Communicating Artery |
| HemStroke | hemorrhagic stroke | ICD10 | I60.22 | Nontraumatic Subarachnoid Hemorrhage from left Anterior Communicating Artery |
| HemStroke | hemorrhagic stroke | ICD10 | I60.30 | Nontraumatic subarachnoid hemorrhage from unspecified posterior communicating artery |
| HemStroke | hemorrhagic stroke | ICD10 | I60.31 | Nontraumatic subarachnoid hemorrhage from right posterior communicating artery |
| HemStroke | hemorrhagic stroke | ICD10 | I60.32 | Nontraumatic subarachnoid hemorrhage from left posterior communicating artery |
| HemStroke | hemorrhagic stroke | ICD10 | I60.4 | Nontraumatic subarachnoid hemorrhage from basilar artery |
| HemStroke | hemorrhagic stroke | ICD10 | I60.50 | Nontraumatic subarachnoid hemorrhage from unspecified vertebral artery |

| Phenotype | phenotype description | vocabul ary | code | Code description |
|---|---|---|---|---|
| HemStroke | hemorrhagic stroke | ICD10 | I60.51 | Nontraumatic subarachnoid hemorrhage from right vertebral artery |
| HemStroke | hemorrhagic stroke | ICD10 | I60.52 | Nontraumatic subarachnoid hemorrhage from left vertebral artery |
| HemStroke | hemorrhagic stroke | ICD10 | I60.6 | Nontraumatic subarachnoid hemorrhage from other intracranial arteries |
| HemStroke | hemorrhagic stroke | ICD10 | I60.7 | Nontraumatic subarachnoid hemorrhage from unspecified intracranial artery |
| HemStroke | hemorrhagic stroke | ICD10 | I60.8 | Other nontraumatic subarachnoid hemorrhage |
| HemStroke | hemorrhagic stroke | ICD10 | I60.9 | Nontraumatic subarachnoid hemorrhage, unspecified |
| HemStroke | hemorrhagic stroke | ICD10 | I61.0 | Nontraumatic intracerebral hemorrhage in hemisphere, subcortical |
| HemStroke | hemorrhagic stroke | ICD10 | I61.1 | Nontraumatic intracerebral hemorrhage in hemisphere, cortical |
| HemStroke | hemorrhagic stroke | ICD10 | I61.2 | Nontraumatic intracerebral hemorrhage in hemisphere, unspecified |
| HemStroke | hemorrhagic stroke | ICD10 | I61.3 | Nontraumatic intracerebral hemorrhage in brain stem |

| Phenotype | phenotype description | vocabulary | code | Code description |
|---|---|---|---|---|
| HemStroke | hemorrhagic stroke | ICD10 | I61.4 | Nontraumatic intracerebral hemorrhage in cerebellum |
| HemStroke | hemorrhagic stroke | ICD10 | I61.5 | Nontraumatic intracerebral hemorrhage, intraventricular |
| HemStroke | hemorrhagic stroke | ICD10 | I61.6 | Nontraumatic intracerebral hemorrhage, multiple localized |
| HemStroke | hemorrhagic stroke | ICD10 | I61.8 | Other nontraumatic intracerebral hemorrhage |
| HemStroke | hemorrhagic stroke | ICD10 | I61.9 | Nontraumatic intracerebral hemorrhage, unspecified |
| HemStroke | hemorrhagic stroke | ICD9 | 430 | SUBARACHNOID HEMORRHAGE |
| HemStroke | hemorrhagic stroke | ICD9 | 431 | INTRACEREBRAL HEMORRHAGE |
| IscStroke | ischemic stroke | ICD10 | G45.9 | Transient cerebral ischemic attack, unspecified |
| IscStroke | ischemic stroke | ICD10 | I63.00 | Cerebral infarction due to thrombosis of unspecified precerebral artery |
| IscStroke | ischemic stroke | ICD10 | I63.011 | Cerebral infarction due to thrombosis of right vertebral artery |

| Phenotype | phenotype description | vocabul ary | code | Code description |
|---|---|---|---|---|
| IscStroke | ischemic stroke | ICD10 | I63.012 | Cerebral infarction due to thrombosis of left vertebral artery |
| IscStroke | ischemic stroke | ICD10 | I63.013 | Cerebral infarction due to thrombosis of bilateral vertebral arteries |
| IscStroke | ischemic stroke | ICD10 | I63.019 | Cerebral infarction due to thrombosis of unspecified vertebral artery |
| IscStroke | ischemic stroke | ICD10 | I63.02 | Cerebral infarction due to thrombosis of basilar artery |
| IscStroke | ischemic stroke | ICD10 | I63.031 | Cerebral infarction due to thrombosis of right carotid artery |
| IscStroke | ischemic stroke | ICD10 | I63.032 | Cerebral infarction due to thrombosis of left carotid artery |
| IscStroke | ischemic stroke | ICD10 | I63.033 | Cerebral infarction due to thrombosis of bilateral carotid arteries |
| IscStroke | ischemic stroke | ICD10 | I63.039 | Cerebral infarction due to thrombosis of unspecified carotid artery |
| IscStroke | ischemic stroke | ICD10 | I63.09 | Cerebral infarction due to thrombosis of other precerebral artery |
| IscStroke | ischemic stroke | ICD10 | I63.10 | Cerebral infarction due to embolism of unspecified precerebral artery |

| **Phenotype** | **phenotype description** | **vocabulary** | **code** | **Code description** |
|---|---|---|---|---|
| IscStroke | ischemic stroke | ICD10 | I63.111 | Cerebral infarction due to embolism of right vertebral artery |
| IscStroke | ischemic stroke | ICD10 | I63.112 | Cerebral infarction due to embolism of left vertebral artery |
| IscStroke | ischemic stroke | ICD10 | I63.113 | Cerebral infarction due to embolism of bilateral vertebral arteries |
| IscStroke | ischemic stroke | ICD10 | I63.119 | Cerebral infarction due to embolism of unspecified vertebral artery |
| IscStroke | ischemic stroke | ICD10 | I63.12 | Cerebral infarction due to embolism of basilar artery |
| IscStroke | ischemic stroke | ICD10 | I63.131 | Cerebral infarction due to embolism of right carotid artery |
| IscStroke | ischemic stroke | ICD10 | I63.132 | Cerebral infarction due to embolism of left carotid artery |
| IscStroke | ischemic stroke | ICD10 | I63.133 | Cerebral infarction due to embolism of bilateral carotid arteries |
| IscStroke | ischemic stroke | ICD10 | I63.139 | Cerebral infarction due to embolism of unspecified carotid artery |
| IscStroke | ischemic stroke | ICD10 | I63.19 | Cerebral infarction due to embolism of other precerebral artery |

| Phenotype | phenotype description | vocabulary | code | Code description |
|---|---|---|---|---|
| IscStroke | ischemic stroke | ICD10 | I63.20 | Cerebral infarction due to unspecified occlusion or stenosis of unspecified precerebral arteries |
| IscStroke | ischemic stroke | ICD10 | I63.211 | Cerebral infarction due to unspecified occlusion or stenosis of right vertebral artery |
| IscStroke | ischemic stroke | ICD10 | I63.212 | Cerebral infarction due to unspecified occlusion or stenosis of left vertebral artery |
| IscStroke | ischemic stroke | ICD10 | I63.213 | Cerebral infarction due to unspecified occlusion or stenosis of bilateral vertebral arteries |
| IscStroke | ischemic stroke | ICD10 | I63.219 | Cerebral infarction due to unspecified occlusion or stenosis of unspecified vertebral artery |
| IscStroke | ischemic stroke | ICD10 | I63.22 | Cerebral infarction due to unspecified occlusion or stenosis of basilar artery |
| IscStroke | ischemic stroke | ICD10 | I63.231 | Cerebral infarction due to unspecified occlusion or stenosis of right carotid arteries |
| IscStroke | ischemic stroke | ICD10 | I63.232 | Cerebral infarction due to unspecified occlusion or stenosis of left carotid arteries |
| IscStroke | ischemic stroke | ICD10 | I63.233 | Cerebral infarction due to unspecified occlusion or stenosis of bilateral carotid arteries |
| IscStroke | ischemic stroke | ICD10 | I63.239 | Cerebral infarction due to unspecified occlusion or stenosis of unspecified carotid artery |

| Phenotype | phenotype description | vocabulary | code | Code description |
|---|---|---|---|---|
| IscStroke | ischemic stroke | ICD10 | I63.29 | Cerebral infarction due to unspecified occlusion or stenosis of other precerebral arteries |
| IscStroke | ischemic stroke | ICD10 | I63.30 | Cerebral infarction due to thrombosis of unspecified cerebral artery |
| IscStroke | ischemic stroke | ICD10 | I63.311 | Cerebral infarction due to thrombosis of right middle cerebral artery |
| IscStroke | ischemic stroke | ICD10 | I63.312 | Cerebral infarction due to thrombosis of left middle cerebral artery |
| IscStroke | ischemic stroke | ICD10 | I63.313 | Cerebral infarction due to thrombosis of bilateral middle cerebral arteries |
| IscStroke | ischemic stroke | ICD10 | I63.319 | Cerebral infarction due to thrombosis of unspecified middle cerebral artery |
| IscStroke | ischemic stroke | ICD10 | I63.321 | Cerebral infarction due to thrombosis of right anterior cerebral artery |
| IscStroke | ischemic stroke | ICD10 | I63.322 | Cerebral infarction due to thrombosis of left anterior cerebral artery |
| IscStroke | ischemic stroke | ICD10 | I63.323 | Cerebral infarction due to thrombosis of bilateral anterior cerebral arteries |
| IscStroke | ischemic stroke | ICD10 | I63.329 | Cerebral infarction due to thrombosis of unspecified anterior cerebral artery |

| Phenotype | phenotype description | vocabulary | code | Code description |
|---|---|---|---|---|
| IscStroke | ischemic stroke | ICD10 | I63.331 | Cerebral infarction due to thrombosis of right posterior cerebral artery |
| IscStroke | ischemic stroke | ICD10 | I63.332 | Cerebral infarction due to thrombosis of left posterior cerebral artery |
| IscStroke | ischemic stroke | ICD10 | I63.333 | Cerebral infarction due to thrombosis of bilateral posterior cerebral arteries |
| IscStroke | ischemic stroke | ICD10 | I63.339 | Cerebral infarction due to thrombosis of unspecified posterior cerebral artery |
| IscStroke | ischemic stroke | ICD10 | I63.341 | Cerebral infarction due to thrombosis of right cerebellar artery |
| IscStroke | ischemic stroke | ICD10 | I63.342 | Cerebral infarction due to thrombosis of left cerebellar artery |
| IscStroke | ischemic stroke | ICD10 | I63.343 | Cerebral infarction due to thrombosis of bilateral cerebellar arteries |
| IscStroke | ischemic stroke | ICD10 | I63.349 | Cerebral infarction due to thrombosis of unspecified cerebellar artery |
| IscStroke | ischemic stroke | ICD10 | I63.39 | Cerebral infarction due to thrombosis of other cerebral artery |
| IscStroke | ischemic stroke | ICD10 | I63.40 | Cerebral infarction due to embolism of unspecified cerebral artery |

| Phenotype | phenotype description | vocabulary | code | Code description |
|---|---|---|---|---|
| IscStroke | ischemic stroke | ICD10 | I63.411 | Cerebral infarction due to embolism of right middle cerebral artery |
| IscStroke | ischemic stroke | ICD10 | I63.412 | Cerebral infarction due to embolism of left middle cerebral artery |
| IscStroke | ischemic stroke | ICD10 | I63.413 | Cerebral infarction due to embolism of bilateral middle cerebral arteries |
| IscStroke | ischemic stroke | ICD10 | I63.419 | Cerebral infarction due to embolism of unspecified middle cerebral artery |
| IscStroke | ischemic stroke | ICD10 | I63.421 | Cerebral infarction due to embolism of right anterior cerebral artery |
| IscStroke | ischemic stroke | ICD10 | I63.422 | Cerebral infarction due to embolism of left anterior cerebral artery |
| IscStroke | ischemic stroke | ICD10 | I63.423 | Cerebral infarction due to embolism of bilateral anterior cerebral arteries |
| IscStroke | ischemic stroke | ICD10 | I63.429 | Cerebral infarction due to embolism of unspecified anterior cerebral artery |
| IscStroke | ischemic stroke | ICD10 | I63.431 | Cerebral infarction due to embolism of right posterior cerebral artery |
| IscStroke | ischemic stroke | ICD10 | I63.432 | Cerebral infarction due to embolism of left posterior cerebral artery |

| **Phenotype** | **phenotype description** | **vocabulary** | **code** | **Code description** |
|---|---|---|---|---|
| IscStroke | ischemic stroke | ICD10 | I63.433 | Cerebral infarction due to embolism of bilateral posterior cerebral arteries |
| IscStroke | ischemic stroke | ICD10 | I63.439 | Cerebral infarction due to embolism of unspecified posterior cerebral artery |
| IscStroke | ischemic stroke | ICD10 | I63.441 | Cerebral infarction due to embolism of right cerebellar artery |
| IscStroke | ischemic stroke | ICD10 | I63.442 | Cerebral infarction due to embolism of left cerebellar artery |
| IscStroke | ischemic stroke | ICD10 | I63.443 | Cerebral infarction due to embolism of bilateral cerebellar arteries |
| IscStroke | ischemic stroke | ICD10 | I63.449 | Cerebral infarction due to embolism of unspecified cerebellar artery |
| IscStroke | ischemic stroke | ICD10 | I63.49 | Cerebral infarction due to embolism of other cerebral artery |
| IscStroke | ischemic stroke | ICD10 | I63.50 | Cerebral infarction due to unspecified occlusion or stenosis of unspecified cerebral artery |
| IscStroke | ischemic stroke | ICD10 | I63.511 | Cerebral infarction due to unspecified occlusion or stenosis of right middle cerebral artery |
| IscStroke | ischemic stroke | ICD10 | I63.512 | Cerebral infarction due to unspecified occlusion or stenosis of left middle cerebral artery |

| Phenotype | phenotype description | vocabulary | code | Code description |
|---|---|---|---|---|
| IscStroke | ischemic stroke | ICD10 | I63.513 | Cerebral infarction due to unspecified occlusion or stenosis of bilateral middle cerebral arteries |
| IscStroke | ischemic stroke | ICD10 | I63.519 | Cerebral infarction due to unspecified occlusion or stenosis of unspecified middle cerebral artery |
| IscStroke | ischemic stroke | ICD10 | I63.521 | Cerebral infarction due to unspecified occlusion or stenosis of right anterior cerebral artery |
| IscStroke | ischemic stroke | ICD10 | I63.522 | Cerebral infarction due to unspecified occlusion or stenosis of left anterior cerebral artery |
| IscStroke | ischemic stroke | ICD10 | I63.523 | Cerebral infarction due to unspecified occlusion or stenosis of bilateral anterior cerebral arteries |
| IscStroke | ischemic stroke | ICD10 | I63.529 | Cerebral infarction due to unspecified occlusion or stenosis of unspecified anterior cerebral artery |
| IscStroke | ischemic stroke | ICD10 | I63.531 | Cerebral infarction due to unspecified occlusion or stenosis of right posterior cerebral artery |
| IscStroke | ischemic stroke | ICD10 | I63.532 | Cerebral infarction due to unspecified occlusion or stenosis of left posterior cerebral artery |
| IscStroke | ischemic stroke | ICD10 | I63.533 | Cerebral infarction due to unspecified occlusion or stenosis of bilateral posterior cerebral arteries |
| IscStroke | ischemic stroke | ICD10 | I63.539 | Cerebral infarction due to unspecified occlusion or stenosis of unspecified posterior cerebral artery |

| Phenotype | phenotype description | vocabulary | code | Code description |
|---|---|---|---|---|
| IscStroke | ischemic stroke | ICD10 | I63.541 | Cerebral infarction due to unspecified occlusion or stenosis of right cerebellar artery |
| IscStroke | ischemic stroke | ICD10 | I63.542 | Cerebral infarction due to unspecified occlusion or stenosis of left cerebellar artery |
| IscStroke | ischemic stroke | ICD10 | I63.543 | Cerebral infarction due to unspecified occlusion or stenosis of bilateral cerebellar arteries |
| IscStroke | ischemic stroke | ICD10 | I63.549 | Cerebral infarction due to unspecified occlusion or stenosis of unspecified cerebellar artery |
| IscStroke | ischemic stroke | ICD10 | I63.59 | Cerebral infarction due to unspecified occlusion or stenosis of other cerebral artery |
| IscStroke | ischemic stroke | ICD10 | I63.6 | Cerebral infarction due to cerebral venous thrombosis, nonpyogenic |
| IscStroke | ischemic stroke | ICD10 | I63.8 | Other cerebral infarction |
| IscStroke | ischemic stroke | ICD10 | I63.81 | Other cerebral infarction due to occlusion or stenosis of small artery |
| IscStroke | ischemic stroke | ICD10 | I63.89 | Other cerebral infarction |
| IscStroke | ischemic stroke | ICD10 | I63.9 | Cerebral infarction, unspecified |

| Phenotype | phenotype description | vocabulary | code | Code description |
|---|---|---|---|---|
| IscStroke | ischemic stroke | ICD10 | I66.01 | Occlusion and stenosis of right middle cerebral artery |
| IscStroke | ischemic stroke | ICD10 | I66.02 | Occlusion and stenosis of left middle cerebral artery |
| IscStroke | ischemic stroke | ICD10 | I66.03 | Occlusion and stenosis of bilateral middle cerebral arteries |
| IscStroke | ischemic stroke | ICD10 | I66.09 | Occlusion and stenosis of unspecified middle cerebral artery |
| IscStroke | ischemic stroke | ICD10 | I66.11 | Occlusion and stenosis of right anterior cerebral artery |
| IscStroke | ischemic stroke | ICD10 | I66.12 | Occlusion and stenosis of left anterior cerebral artery |
| IscStroke | ischemic stroke | ICD10 | I66.13 | Occlusion and stenosis of bilateral anterior cerebral arteries |
| IscStroke | ischemic stroke | ICD10 | I66.19 | Occlusion and stenosis of unspecified anterior cerebral artery |
| IscStroke | ischemic stroke | ICD10 | I66.21 | Occlusion and stenosis of right posterior cerebral artery |
| IscStroke | ischemic stroke | ICD10 | I66.22 | Occlusion and stenosis of left posterior cerebral artery |

| Phenotype | phenotype description | vocabulary | code | Code description |
|---|---|---|---|---|
| IscStroke | ischemic stroke | ICD10 | I66.23 | Occlusion and stenosis of bilateral posterior cerebral arteries |
| IscStroke | ischemic stroke | ICD10 | I66.29 | Occlusion and stenosis of unspecified posterior cerebral artery |
| IscStroke | ischemic stroke | ICD10 | I66.3 | Occlusion and stenosis of cerebellar arteries |
| IscStroke | ischemic stroke | ICD10 | I66.8 | Occlusion and stenosis of other cerebral arteries |
| IscStroke | ischemic stroke | ICD10 | I66.9 | Occlusion and stenosis of unspecified cerebral artery |
| IscStroke | ischemic stroke | ICD9 | 433.01 | OCCLUSION & STENOSIS OF BASILAR ARTERY, W/CEREBRAL INFARCTION |
| IscStroke | ischemic stroke | ICD9 | 433.11 | OCCLUSION & STENOSIS OF CAROTID ARTERY, W/ CEREBRAL INFARCTION |
| IscStroke | ischemic stroke | ICD9 | 433.21 | OCCLUSION & STENOSIS OF VERTEBRAL ARTERY, W/ CEREBRAL INFARCTION |
| IscStroke | ischemic stroke | ICD9 | 433.31 | OCCLUSION& STENOSIS OF MULTIPLE & BILATERAL ARTERIES, W/ CEREBRAL INFARCTION |
| IscStroke | ischemic stroke | ICD9 | 433.81 | OCCLUSION & STENOSIS OF OTHER SPECIFIED PRECEREBRAL ARTERY, W/ CEREBRAL INFARCTION |

| Phenotype | phenotype description | vocabul ary | code | Code description |
|---|---|---|---|---|
| IscStroke | ischemic stroke | ICD9 | 433.91 | OCCLUSION & STENOSIS OF UNSPECIFIED PRECEREBRAL ARTERY W/ CEREBRAL INFARCTION |
| IscStroke | ischemic stroke | ICD9 | 434.01 | CEREBRAL THROMBOSIS W/ CEREBRAL INFARCTION |
| IscStroke | ischemic stroke | ICD9 | 434.11 | CEREBRAL EMBOLISM W/ CEREBRAL INFARCTION |
| IscStroke | ischemic stroke | ICD9 | 434.91 | CEREBRAL ARTERY OCCLUSION, UNSPECIFIED, W/ CEREBRAL INFARCTION |
| IscStroke | ischemic stroke | ICD9 | 435.9 | UNSPECIFIED TRANSIENT CEREBRAL ISCHEMIA |
| LewyBodyDem | Lewy Body dementia | ICD10 | G31.83 | Dementia with Lewy bodies |
| LewyBodyDem | Lewy Body dementia | ICD9 | 331.82 | DEMENTIA WITH LEWY BODIES |
| OthDem | other related dementia | ICD10 | F02.80 | Dementia in other diseases classified elsewhere without behavioral disturbance |
| OthDem | other related dementia | ICD10 | F02.81 | Dementia in other diseases classified elsewhere with behavioral disturbance |
| OthDem | other related dementia | ICD10 | F03.90 | Unspecified dementia without behavioral disturbance |

| Phenotype | phenotype description | vocabul ary | code | Code description |
|---|---|---|---|---|
| OthDem | other related dementia | ICD10 | F03.91 | Unspecified dementia with behavioral disturbance |
| OthDem | other related dementia | ICD10 | F10.27 | Alcohol dependence with alcohol-induced persisting dementia |
| OthDem | other related dementia | ICD10 | F19.97 | Other psychoactive substance use, unspecified with psychoactive substance-induced persisting dementia |
| OthDem | other related dementia | ICD10 | G10. | Huntington's disease |
| OthDem | other related dementia | ICD10 | G31.01 | Pick's disease |
| OthDem | other related dementia | ICD9 | 290 | SENILE DEMENTIA, UNCOMPLICATED |
| OthDem | other related dementia | ICD9 | 290.1 | PRESENILE DEMENTIA, UNCOMPLICATED |
| OthDem | other related dementia | ICD9 | 290.11 | PRESENILE DEMENTIA WITH DELIRIUM |
| OthDem | other related dementia | ICD9 | 290.12 | PRESENILE DEMENTIA WITH DELUSIONAL FEATURES |
| OthDem | other related dementia | ICD9 | 290.13 | PRESENILE DEMENTIA WITH DEPRESSIVE FEATURES |

| Phenotype | phenotype description | vocabulary | code | Code description |
|---|---|---|---|---|
| OthDem | other related dementia | ICD9 | 290.2 | SENILE DEMENTIA WITH DELUSIONAL FEATURES |
| OthDem | other related dementia | ICD9 | 290.21 | SENILE DEMENTIA WITH DEPRESSIVE FEATURES |
| OthDem | other related dementia | ICD9 | 290.3 | SENILE DEMENTIA WITH DELIRIUM |
| OthDem | other related dementia | ICD9 | 291.2 | ALCOHOL-INDUCED PERSISTING DEMENTIA |
| OthDem | other related dementia | ICD9 | 291.2 | DEMENTIA ASSOCIATED WITH ALCOHOLISM, UNSPECIFIED |
| OthDem | other related dementia | ICD9 | 291.21 | DEMENTIA ASSOCIATED WITH ALCOHOLISM, MILD |
| OthDem | other related dementia | ICD9 | 291.22 | DEMENTIA ASSOCIATED WITH ALCOHOLISM, MODERATE |
| OthDem | other related dementia | ICD9 | 291.23 | DEMENTIA ASSOCIATED WITH ALCOHOLISM SEVERE |
| OthDem | other related dementia | ICD9 | 292.82 | DRUG-INDUCED PERSISTING DEMENTIA |
| OthDem | other related dementia | ICD9 | 294 | AMNESTIC DISORDER IN CONDITIONS CLASSIFIED ELSEWHERE |

| Phenotype | phenotype description | vocabulary | code | Code description |
|---|---|---|---|---|
| OthDem | other related dementia | ICD9 | 294.1 | DEMENTIA IN CONDITIONS CLASSIFIED ELSEWHERE |
| OthDem | other related dementia | ICD9 | 294.1 | DEMENTIA IN CONDITIONS CLASSIFIED ELSEWHERE WITHOUT BEHAVIORAL DISTURBANCE |
| OthDem | other related dementia | ICD9 | 294.11 | DEMENTIA IN CONDITIONS CLASSIFIED ELSEWHERE WITH BEHAVIORAL DISTURBANCE |
| OthDem | other related dementia | ICD9 | 294.2 | DEMENTIA, UNSPECIFIED, WITHOUT BEHAVIORAL DISTURBANCE |
| OthDem | other related dementia | ICD9 | 294.21 | DEMENTIA, UNSPECIFIED, WITH BEHAVIORAL DISTURBANCE |
| OthDem | other related dementia | ICD9 | 294.8 | OTHER PERSISTENT MENTAL DISORDERS DUE TO CONDITIONS CLASSIFIED ELSEWHERE |
| OthDem | other related dementia | ICD9 | 331.2 | SENILE DEGENERATION OF BRAIN |
| OthDem | other related dementia | ICD9 | 797 | SENILITY WITHOUT MENTION OF PSYCHOSIS |
| PAD | Peripheral Arterial Disease/Thrombosis | ICD10 | E08.52 | Diabetes mellitus due to underlying condition with diabetic peripheral angiopathy with gangrene |
| PAD | Peripheral Arterial Disease/Thrombosis | ICD10 | E09.52 | Drug or chemical induced diabetes mellitus with diabetic peripheral angiopathy with gangrene |

| Phenotype | phenotype description | vocabulary | code | Code description |
|---|---|---|---|---|
| PAD | Peripheral Arterial Disease/Thrombosis | ICD10 | E10.51 | Type 1 diabetes mellitus with diabetic peripheral angiopathy without gangrene |
| PAD | Peripheral Arterial Disease/Thrombosis | ICD10 | E10.52 | Type 1 diabetes mellitus with diabetic peripheral angiopathy with gangrene |
| PAD | Peripheral Arterial Disease/Thrombosis | ICD10 | E10.59 | Type 1 diabetes mellitus with other circulatory complications |
| PAD | Peripheral Arterial Disease/Thrombosis | ICD10 | E11.51 | Type 2 diabetes mellitus with diabetic peripheral angiopathy without gangrene |
| PAD | Peripheral Arterial Disease/Thrombosis | ICD10 | E11.52 | Type 2 diabetes mellitus with diabetic peripheral angiopathy with gangrene |
| PAD | Peripheral Arterial Disease/Thrombosis | ICD10 | E11.59 | Type 2 diabetes mellitus with other circulatory complications |
| PAD | Peripheral Arterial Disease/Thrombosis | ICD10 | E13.51 | Other specified diabetes mellitus with diabetic peripheral angiopathy without gangrene |
| PAD | Peripheral Arterial Disease/Thrombosis | ICD10 | E13.52 | Other specified diabetes mellitus with diabetic peripheral angiopathy with gangrene |
| PAD | Peripheral Arterial Disease/Thrombosis | ICD10 | E13.59 | Other specified diabetes mellitus with other circulatory complications |
| PAD | Peripheral Arterial Disease/Thrombosis | ICD10 | I70.0 | Atherosclerosis of aorta |

| Phenotype | phenotype description | vocabulary | code | Code description |
|---|---|---|---|---|
| PAD | Peripheral Arterial Disease/Thrombosis | ICD10 | I70.201 | Unspecified atherosclerosis of native arteries of extremities, right leg |
| PAD | Peripheral Arterial Disease/Thrombosis | ICD10 | I70.202 | Unspecified atherosclerosis of native arteries of extremities, left leg |
| PAD | Peripheral Arterial Disease/Thrombosis | ICD10 | I70.203 | Unspecified atherosclerosis of native arteries of extremities, bilateral legs |
| PAD | Peripheral Arterial Disease/Thrombosis | ICD10 | I70.208 | Unspecified atherosclerosis of native arteries of extremities, other extremity |
| PAD | Peripheral Arterial Disease/Thrombosis | ICD10 | I70.209 | Unspecified atherosclerosis of native arteries of extremities, unspecified extremity |
| PAD | Peripheral Arterial Disease/Thrombosis | ICD10 | I70.211 | Atherosclerosis of native arteries of extremities with intermittent claudication, right leg |
| PAD | Peripheral Arterial Disease/Thrombosis | ICD10 | I70.212 | Atherosclerosis of native arteries of extremities with intermittent claudication, left leg |
| PAD | Peripheral Arterial Disease/Thrombosis | ICD10 | I70.213 | Atherosclerosis of native arteries of extremities with intermittent claudication, bilateral legs |
| PAD | Peripheral Arterial Disease/Thrombosis | ICD10 | I70.218 | Atherosclerosis of native arteries of extremities with intermittent claudication, other extremity |
| PAD | Peripheral Arterial Disease/Thrombosis | ICD10 | I70.219 | Atherosclerosis of native arteries of extremities with intermittent claudication, unspecified extremity |

| Phenotype | phenotype description | vocabulary | code | Code description |
|---|---|---|---|---|
| PAD | Peripheral Arterial Disease/Thrombosis | ICD10 | I70.221 | Atherosclerosis of native arteries of extremities with rest pain, right leg |
| PAD | Peripheral Arterial Disease/Thrombosis | ICD10 | I70.222 | Atherosclerosis of native arteries of extremities with rest pain, left leg |
| PAD | Peripheral Arterial Disease/Thrombosis | ICD10 | I70.223 | Atherosclerosis of native arteries of extremities with rest pain, bilateral legs |
| PAD | Peripheral Arterial Disease/Thrombosis | ICD10 | I70.228 | Atherosclerosis of native arteries of extremities with rest pain, other extremity |
| PAD | Peripheral Arterial Disease/Thrombosis | ICD10 | I70.229 | Atherosclerosis of native arteries of extremities with rest pain, unspecified extremity |
| PAD | Peripheral Arterial Disease/Thrombosis | ICD10 | I70.231 | Atherosclerosis of native arteries of right leg with ulceration of thigh |
| PAD | Peripheral Arterial Disease/Thrombosis | ICD10 | I70.232 | Atherosclerosis of native arteries of right leg with ulceration of calf |
| PAD | Peripheral Arterial Disease/Thrombosis | ICD10 | I70.233 | Atherosclerosis of native arteries of right leg with ulceration of ankle |
| PAD | Peripheral Arterial Disease/Thrombosis | ICD10 | I70.234 | Atherosclerosis of native arteries of right leg with ulceration of heel and midfoot |
| PAD | Peripheral Arterial Disease/Thrombosis | ICD10 | I70.235 | Atherosclerosis of native arteries of right leg with ulceration of other part of foot |

| Phenotype | phenotype description | vocabulary | code | Code description |
|---|---|---|---|---|
| PAD | Peripheral Arterial Disease/Thrombosis | ICD10 | I70.238 | Atherosclerosis of native arteries of right leg with ulceration of other part of lower leg |
| PAD | Peripheral Arterial Disease/Thrombosis | ICD10 | I70.239 | Atherosclerosis of native arteries of right leg with ulceration of unspecified site |
| PAD | Peripheral Arterial Disease/Thrombosis | ICD10 | I70.241 | Atherosclerosis of native arteries of left leg with ulceration of thigh |
| PAD | Peripheral Arterial Disease/Thrombosis | ICD10 | I70.242 | Atherosclerosis of native arteries of left leg with ulceration of calf |
| PAD | Peripheral Arterial Disease/Thrombosis | ICD10 | I70.243 | Atherosclerosis of native arteries of left leg with ulceration of ankle |
| PAD | Peripheral Arterial Disease/Thrombosis | ICD10 | I70.244 | Atherosclerosis of native arteries of left leg with ulceration of heel and midfoot |
| PAD | Peripheral Arterial Disease/Thrombosis | ICD10 | I70.245 | Atherosclerosis of native arteries of left leg with ulceration of other part of foot |
| PAD | Peripheral Arterial Disease/Thrombosis | ICD10 | I70.248 | Atherosclerosis of native arteries of left leg with ulceration of other part of lower leg |
| PAD | Peripheral Arterial Disease/Thrombosis | ICD10 | I70.249 | Atherosclerosis of native arteries of left leg with ulceration of unspecified site |
| PAD | Peripheral Arterial Disease/Thrombosis | ICD10 | I70.25 | Atherosclerosis of native arteries of other extremities with ulceration |

| Phenotype | phenotype description | vocabulary | code | Code description |
|---|---|---|---|---|
| PAD | Peripheral Arterial Disease/Thrombosis | ICD10 | I70.261 | Atherosclerosis of native arteries of extremities with gangrene, right leg |
| PAD | Peripheral Arterial Disease/Thrombosis | ICD10 | I70.262 | Atherosclerosis of native arteries of extremities with gangrene, left leg |
| PAD | Peripheral Arterial Disease/Thrombosis | ICD10 | I70.263 | Atherosclerosis of native arteries of extremities with gangrene, bilateral legs |
| PAD | Peripheral Arterial Disease/Thrombosis | ICD10 | I70.268 | Atherosclerosis of native arteries of extremities with gangrene, other extremity |
| PAD | Peripheral Arterial Disease/Thrombosis | ICD10 | I70.269 | Atherosclerosis of native arteries of extremities with gangrene, unspecified extremity |
| PAD | Peripheral Arterial Disease/Thrombosis | ICD10 | I70.291 | Other atherosclerosis of native arteries of extremities, right leg |
| PAD | Peripheral Arterial Disease/Thrombosis | ICD10 | I70.292 | Other atherosclerosis of native arteries of extremities, left leg |
| PAD | Peripheral Arterial Disease/Thrombosis | ICD10 | I70.293 | Other atherosclerosis of native arteries of extremities, bilateral legs |
| PAD | Peripheral Arterial Disease/Thrombosis | ICD10 | I70.298 | Other atherosclerosis of native arteries of extremities, other extremity |
| PAD | Peripheral Arterial Disease/Thrombosis | ICD10 | I70.299 | Other atherosclerosis of native arteries of extremities, unspecified extremity |

| Phenotype | phenotype description | vocabulary | code | Code description |
|---|---|---|---|---|
| PAD | Peripheral Arterial Disease/Thrombosis | ICD10 | I70.301 | Unspecified atherosclerosis of unspecified type of bypass graft(s) of the extremities, right leg |
| PAD | Peripheral Arterial Disease/Thrombosis | ICD10 | I70.302 | Unspecified atherosclerosis of unspecified type of bypass graft(s) of the extremities, left leg |
| PAD | Peripheral Arterial Disease/Thrombosis | ICD10 | I70.303 | Unspecified atherosclerosis of unspecified type of bypass graft(s) of the extremities, bilateral legs |
| PAD | Peripheral Arterial Disease/Thrombosis | ICD10 | I70.308 | Unspecified atherosclerosis of unspecified type of bypass graft(s) of the extremities, other extremity |
| PAD | Peripheral Arterial Disease/Thrombosis | ICD10 | I70.309 | Unspecified atherosclerosis of unspecified type of bypass graft(s) of the extremities, unspecified extremity |
| PAD | Peripheral Arterial Disease/Thrombosis | ICD10 | I70.311 | Atherosclerosis of unspecified type of bypass graft(s) of the extremities with intermittent claudication, right leg |
| PAD | Peripheral Arterial Disease/Thrombosis | ICD10 | I70.312 | Atherosclerosis of unspecified type of bypass graft(s) of the extremities with intermittent claudication, left leg |
| PAD | Peripheral Arterial Disease/Thrombosis | ICD10 | I70.313 | Atherosclerosis of unspecified type of bypass graft(s) of the extremities with intermittent claudication, bilateral legs |
| PAD | Peripheral Arterial Disease/Thrombosis | ICD10 | I70.318 | Atherosclerosis of unspecified type of bypass graft(s) of the extremities with intermittent claudication, other extremity |
| PAD | Peripheral Arterial Disease/Thrombosis | ICD10 | I70.319 | Atherosclerosis of unspecified type of bypass graft(s) of the extremities with intermittent claudication, unspecified extremity |

| Phenotype | phenotype description | vocabulary | code | Code description |
|---|---|---|---|---|
| PAD | Peripheral Arterial Disease/Thrombosis | ICD10 | I70.321 | Atherosclerosis of unspecified type of bypass graft(s) of the extremities with rest pain, right leg |
| PAD | Peripheral Arterial Disease/Thrombosis | ICD10 | I70.322 | Atherosclerosis of unspecified type of bypass graft(s) of the extremities with rest pain, left leg |
| PAD | Peripheral Arterial Disease/Thrombosis | ICD10 | I70.323 | Atherosclerosis of unspecified type of bypass graft(s) of the extremities with rest pain, bilateral legs |
| PAD | Peripheral Arterial Disease/Thrombosis | ICD10 | I70.328 | Atherosclerosis of unspecified type of bypass graft(s) of the extremities with rest pain, other extremity |
| PAD | Peripheral Arterial Disease/Thrombosis | ICD10 | I70.329 | Atherosclerosis of unspecified type of bypass graft(s) of the extremities with rest pain, unspecified extremity |
| PAD | Peripheral Arterial Disease/Thrombosis | ICD10 | I70.331 | Atherosclerosis of unspecified type of bypass graft(s) of the right leg with ulceration of thigh |
| PAD | Peripheral Arterial Disease/Thrombosis | ICD10 | I70.332 | Atherosclerosis of unspecified type of bypass graft(s) of the right leg with ulceration of calf |
| PAD | Peripheral Arterial Disease/Thrombosis | ICD10 | I70.333 | Atherosclerosis of unspecified type of bypass graft(s) of the right leg with ulceration of ankle |
| PAD | Peripheral Arterial Disease/Thrombosis | ICD10 | I70.334 | Atherosclerosis of unspecified type of bypass graft(s) of the right leg with ulceration of heel and midfoot |
| PAD | Peripheral Arterial Disease/Thrombosis | ICD10 | I70.335 | Atherosclerosis of unspecified type of bypass graft(s) of the right leg with ulceration of other part of foot |

| Phenotype | phenotype description | vocabulary | code | Code description |
|---|---|---|---|---|
| PAD | Peripheral Arterial Disease/Thrombosis | ICD10 | I70.338 | Atherosclerosis of unspecified type of bypass graft(s) of the right leg with ulceration of other part of lower leg |
| PAD | Peripheral Arterial Disease/Thrombosis | ICD10 | I70.339 | Atherosclerosis of unspecified type of bypass graft(s) of the right leg with ulceration of unspecified site |
| PAD | Peripheral Arterial Disease/Thrombosis | ICD10 | I70.341 | Atherosclerosis of unspecified type of bypass graft(s) of the left leg with ulceration of thigh |
| PAD | Peripheral Arterial Disease/Thrombosis | ICD10 | I70.342 | Atherosclerosis of unspecified type of bypass graft(s) of the left leg with ulceration of calf |
| PAD | Peripheral Arterial Disease/Thrombosis | ICD10 | I70.343 | Atherosclerosis of unspecified type of bypass graft(s) of the left leg with ulceration of ankle |
| PAD | Peripheral Arterial Disease/Thrombosis | ICD10 | I70.344 | Atherosclerosis of unspecified type of bypass graft(s) of the left leg with ulceration of heel and midfoot |
| PAD | Peripheral Arterial Disease/Thrombosis | ICD10 | I70.345 | Atherosclerosis of unspecified type of bypass graft(s) of the left leg with ulceration of other part of foot |
| PAD | Peripheral Arterial Disease/Thrombosis | ICD10 | I70.348 | Atherosclerosis of unspecified type of bypass graft(s) of the left leg with ulceration of other part of lower leg |
| PAD | Peripheral Arterial Disease/Thrombosis | ICD10 | I70.349 | Atherosclerosis of unspecified type of bypass graft(s) of the left leg with ulceration of unspecified site |
| PAD | Peripheral Arterial Disease/Thrombosis | ICD10 | I70.35 | Atherosclerosis of unspecified type of bypass graft(s) of other extremity with ulceration |

| Phenotype | phenotype description | vocabulary | code | Code description |
|---|---|---|---|---|
| PAD | Peripheral Arterial Disease/Thrombosis | ICD10 | I70.361 | Atherosclerosis of unspecified type of bypass graft(s) of the extremities with gangrene, right leg |
| PAD | Peripheral Arterial Disease/Thrombosis | ICD10 | I70.362 | Atherosclerosis of unspecified type of bypass graft(s) of the extremities with gangrene, left leg |
| PAD | Peripheral Arterial Disease/Thrombosis | ICD10 | I70.363 | Atherosclerosis of unspecified type of bypass graft(s) of the extremities with gangrene, bilateral legs |
| PAD | Peripheral Arterial Disease/Thrombosis | ICD10 | I70.368 | Atherosclerosis of unspecified type of bypass graft(s) of the extremities with gangrene, other extremity |
| PAD | Peripheral Arterial Disease/Thrombosis | ICD10 | I70.369 | Atherosclerosis of unspecified type of bypass graft(s) of the extremities with gangrene, unspecified extremity |
| PAD | Peripheral Arterial Disease/Thrombosis | ICD10 | I70.391 | Other atherosclerosis of unspecified type of bypass graft(s) of the extremities, right leg |
| PAD | Peripheral Arterial Disease/Thrombosis | ICD10 | I70.392 | Other atherosclerosis of unspecified type of bypass graft(s) of the extremities, left leg |
| PAD | Peripheral Arterial Disease/Thrombosis | ICD10 | I70.393 | Other atherosclerosis of unspecified type of bypass graft(s) of the extremities, bilateral legs |
| PAD | Peripheral Arterial Disease/Thrombosis | ICD10 | I70.398 | Other atherosclerosis of unspecified type of bypass graft(s) of the extremities, other extremity |
| PAD | Peripheral Arterial Disease/Thrombosis | ICD10 | I70.399 | Other atherosclerosis of unspecified type of bypass graft(s) of the extremities, unspecified extremity |

| Phenotype | phenotype description | vocabulary | code | Code description |
|---|---|---|---|---|
| PAD | Peripheral Arterial Disease/Thrombosis | ICD10 | I70.401 | Unspecified atherosclerosis of autologous vein bypass graft(s) of the extremities, right leg |
| PAD | Peripheral Arterial Disease/Thrombosis | ICD10 | I70.402 | Unspecified atherosclerosis of autologous vein bypass graft(s) of the extremities, left leg |
| PAD | Peripheral Arterial Disease/Thrombosis | ICD10 | I70.403 | Unspecified atherosclerosis of autologous vein bypass graft(s) of the extremities, bilateral legs |
| PAD | Peripheral Arterial Disease/Thrombosis | ICD10 | I70.408 | Unspecified atherosclerosis of autologous vein bypass graft(s) of the extremities, other extremity |
| PAD | Peripheral Arterial Disease/Thrombosis | ICD10 | I70.409 | Unspecified atherosclerosis of autologous vein bypass graft(s) of the extremities, unspecified extremity |
| PAD | Peripheral Arterial Disease/Thrombosis | ICD10 | I70.411 | Atherosclerosis of autologous vein bypass graft(s) of the extremities with intermittent claudication, right leg |
| PAD | Peripheral Arterial Disease/Thrombosis | ICD10 | I70.412 | Atherosclerosis of autologous vein bypass graft(s) of the extremities with intermittent claudication, left leg |
| PAD | Peripheral Arterial Disease/Thrombosis | ICD10 | I70.413 | Atherosclerosis of autologous vein bypass graft(s) of the extremities with intermittent claudication, bilateral legs |
| PAD | Peripheral Arterial Disease/Thrombosis | ICD10 | I70.418 | Atherosclerosis of autologous vein bypass graft(s) of the extremities with intermittent claudication, other extremity |
| PAD | Peripheral Arterial Disease/Thrombosis | ICD10 | I70.419 | Atherosclerosis of autologous vein bypass graft(s) of the extremities with intermittent claudication, unspecified extremity |

| Phenotype | phenotype description | vocabulary | code | Code description |
|---|---|---|---|---|
| PAD | Peripheral Arterial Disease/Thrombosis | ICD10 | I70.421 | Atherosclerosis of autologous vein bypass graft(s) of the extremities with rest pain, right leg |
| PAD | Peripheral Arterial Disease/Thrombosis | ICD10 | I70.422 | Atherosclerosis of autologous vein bypass graft(s) of the extremities with rest pain, left leg |
| PAD | Peripheral Arterial Disease/Thrombosis | ICD10 | I70.423 | Atherosclerosis of autologous vein bypass graft(s) of the extremities with rest pain, bilateral legs |
| PAD | Peripheral Arterial Disease/Thrombosis | ICD10 | I70.428 | Atherosclerosis of autologous vein bypass graft(s) of the extremities with rest pain, other extremity |
| PAD | Peripheral Arterial Disease/Thrombosis | ICD10 | I70.429 | Atherosclerosis of autologous vein bypass graft(s) of the extremities with rest pain, unspecified extremity |
| PAD | Peripheral Arterial Disease/Thrombosis | ICD10 | I70.431 | Atherosclerosis of autologous vein bypass graft(s) of the right leg with ulceration of thigh |
| PAD | Peripheral Arterial Disease/Thrombosis | ICD10 | I70.432 | Atherosclerosis of autologous vein bypass graft(s) of the right leg with ulceration of calf |
| PAD | Peripheral Arterial Disease/Thrombosis | ICD10 | I70.433 | Atherosclerosis of autologous vein bypass graft(s) of the right leg with ulceration of ankle |
| PAD | Peripheral Arterial Disease/Thrombosis | ICD10 | I70.434 | Atherosclerosis of autologous vein bypass graft(s) of the right leg with ulceration of heel and midfoot |
| PAD | Peripheral Arterial Disease/Thrombosis | ICD10 | I70.435 | Atherosclerosis of autologous vein bypass graft(s) of the right leg with ulceration of other part of foot |

| Phenotype | phenotype description | vocabulary | code | Code description |
|---|---|---|---|---|
| PAD | Peripheral Arterial Disease/Thrombosis | ICD10 | I70.438 | Atherosclerosis of autologous vein bypass graft(s) of the right leg with ulceration of other part of lower leg |
| PAD | Peripheral Arterial Disease/Thrombosis | ICD10 | I70.439 | Atherosclerosis of autologous vein bypass graft(s) of the right leg with ulceration of unspecified site |
| PAD | Peripheral Arterial Disease/Thrombosis | ICD10 | I70.441 | Atherosclerosis of autologous vein bypass graft(s) of the left leg with ulceration of thigh |
| PAD | Peripheral Arterial Disease/Thrombosis | ICD10 | I70.442 | Atherosclerosis of autologous vein bypass graft(s) of the left leg with ulceration of calf |
| PAD | Peripheral Arterial Disease/Thrombosis | ICD10 | I70.443 | Atherosclerosis of autologous vein bypass graft(s) of the left leg with ulceration of ankle |
| PAD | Peripheral Arterial Disease/Thrombosis | ICD10 | I70.444 | Atherosclerosis of autologous vein bypass graft(s) of the left leg with ulceration of heel and midfoot |
| PAD | Peripheral Arterial Disease/Thrombosis | ICD10 | I70.445 | Atherosclerosis of autologous vein bypass graft(s) of the left leg with ulceration of other part of foot |
| PAD | Peripheral Arterial Disease/Thrombosis | ICD10 | I70.448 | Atherosclerosis of autologous vein bypass graft(s) of the left leg with ulceration of other part of lower leg |
| PAD | Peripheral Arterial Disease/Thrombosis | ICD10 | I70.449 | Atherosclerosis of autologous vein bypass graft(s) of the left leg with ulceration of unspecified site |
| PAD | Peripheral Arterial Disease/Thrombosis | ICD10 | I70.45 | Atherosclerosis of autologous vein bypass graft(s) of other extremity with ulceration |

| Phenotype | phenotype description | vocabulary | code | Code description |
|---|---|---|---|---|
| PAD | Peripheral Arterial Disease/Thrombosis | ICD10 | I70.461 | Atherosclerosis of autologous vein bypass graft(s) of the extremities with gangrene, right leg |
| PAD | Peripheral Arterial Disease/Thrombosis | ICD10 | I70.462 | Atherosclerosis of autologous vein bypass graft(s) of the extremities with gangrene, left leg |
| PAD | Peripheral Arterial Disease/Thrombosis | ICD10 | I70.463 | Atherosclerosis of autologous vein bypass graft(s) of the extremities with gangrene, bilateral legs |
| PAD | Peripheral Arterial Disease/Thrombosis | ICD10 | I70.468 | Atherosclerosis of autologous vein bypass graft(s) of the extremities with gangrene, other extremity |
| PAD | Peripheral Arterial Disease/Thrombosis | ICD10 | I70.469 | Atherosclerosis of autologous vein bypass graft(s) of the extremities with gangrene, unspecified extremity |
| PAD | Peripheral Arterial Disease/Thrombosis | ICD10 | I70.491 | Other atherosclerosis of autologous vein bypass graft(s) of the extremities, right leg |
| PAD | Peripheral Arterial Disease/Thrombosis | ICD10 | I70.492 | Other atherosclerosis of autologous vein bypass graft(s) of the extremities, left leg |
| PAD | Peripheral Arterial Disease/Thrombosis | ICD10 | I70.493 | Other atherosclerosis of autologous vein bypass graft(s) of the extremities, bilateral legs |
| PAD | Peripheral Arterial Disease/Thrombosis | ICD10 | I70.498 | Other atherosclerosis of autologous vein bypass graft(s) of the extremities, other extremity |
| PAD | Peripheral Arterial Disease/Thrombosis | ICD10 | I70.499 | Other atherosclerosis of autologous vein bypass graft(s) of the extremities, unspecified extremity |

| Phenotype | phenotype description | vocabulary | code | Code description |
|---|---|---|---|---|
| PAD | Peripheral Arterial Disease/Thrombosis | ICD10 | I70.501 | Unspecified atherosclerosis of nonautologous biological bypass graft(s) of the extremities, right leg |
| PAD | Peripheral Arterial Disease/Thrombosis | ICD10 | I70.502 | Unspecified atherosclerosis of nonautologous biological bypass graft(s) of the extremities, left leg |
| PAD | Peripheral Arterial Disease/Thrombosis | ICD10 | I70.503 | Unspecified atherosclerosis of nonautologous biological bypass graft(s) of the extremities, bilateral legs |
| PAD | Peripheral Arterial Disease/Thrombosis | ICD10 | I70.508 | Unspecified atherosclerosis of nonautologous biological bypass graft(s) of the extremities, other extremity |
| PAD | Peripheral Arterial Disease/Thrombosis | ICD10 | I70.509 | Unspecified atherosclerosis of nonautologous biological bypass graft(s) of the extremities, unspecified extremity |
| PAD | Peripheral Arterial Disease/Thrombosis | ICD10 | I70.511 | Atherosclerosis of nonautologous biological bypass graft(s) of the extremities with intermittent claudication, right leg |
| PAD | Peripheral Arterial Disease/Thrombosis | ICD10 | I70.512 | Atherosclerosis of nonautologous biological bypass graft(s) of the extremities with intermittent claudication, left leg |
| PAD | Peripheral Arterial Disease/Thrombosis | ICD10 | I70.513 | Atherosclerosis of nonautologous biological bypass graft(s) of the extremities with intermittent claudication, bilateral legs |
| PAD | Peripheral Arterial Disease/Thrombosis | ICD10 | I70.518 | Atherosclerosis of nonautologous biological bypass graft(s) of the extremities with intermittent claudication, other extremity |
| PAD | Peripheral Arterial Disease/Thrombosis | ICD10 | I70.519 | Atherosclerosis of nonautologous biological bypass graft(s) of the extremities with intermittent claudication, unspecified extremity |

| Phenotype | phenotype description | vocabulary | code | Code description |
|---|---|---|---|---|
| PAD | Peripheral Arterial Disease/Thrombosis | ICD10 | I70.521 | Atherosclerosis of nonautologous biological bypass graft(s) of the extremities with rest pain, right leg |
| PAD | Peripheral Arterial Disease/Thrombosis | ICD10 | I70.522 | Atherosclerosis of nonautologous biological bypass graft(s) of the extremities with rest pain, left leg |
| PAD | Peripheral Arterial Disease/Thrombosis | ICD10 | I70.523 | Atherosclerosis of nonautologous biological bypass graft(s) of the extremities with rest pain, bilateral legs |
| PAD | Peripheral Arterial Disease/Thrombosis | ICD10 | I70.528 | Atherosclerosis of nonautologous biological bypass graft(s) of the extremities with rest pain, other extremity |
| PAD | Peripheral Arterial Disease/Thrombosis | ICD10 | I70.529 | Atherosclerosis of nonautologous biological bypass graft(s) of the extremities with rest pain, unspecified extremity |
| PAD | Peripheral Arterial Disease/Thrombosis | ICD10 | I70.531 | Atherosclerosis of nonautologous biological bypass graft(s) of the right leg with ulceration of thigh |
| PAD | Peripheral Arterial Disease/Thrombosis | ICD10 | I70.532 | Atherosclerosis of nonautologous biological bypass graft(s) of the right leg with ulceration of calf |
| PAD | Peripheral Arterial Disease/Thrombosis | ICD10 | I70.533 | Atherosclerosis of nonautologous biological bypass graft(s) of the right leg with ulceration of ankle |
| PAD | Peripheral Arterial Disease/Thrombosis | ICD10 | I70.534 | Atherosclerosis of nonautologous biological bypass graft(s) of the right leg with ulceration of heel and midfoot |
| PAD | Peripheral Arterial Disease/Thrombosis | ICD10 | I70.535 | Atherosclerosis of nonautologous biological bypass graft(s) of the right leg with ulceration of other part of foot |

| Phenotype | phenotype description | vocabulary | code | Code description |
|---|---|---|---|---|
| PAD | Peripheral Arterial Disease/Thrombosis | ICD10 | I70.538 | Atherosclerosis of nonautologous biological bypass graft(s) of the right leg with ulceration of other part of lower leg |
| PAD | Peripheral Arterial Disease/Thrombosis | ICD10 | I70.539 | Atherosclerosis of nonautologous biological bypass graft(s) of the right leg with ulceration of unspecified site |
| PAD | Peripheral Arterial Disease/Thrombosis | ICD10 | I70.541 | Atherosclerosis of nonautologous biological bypass graft(s) of the left leg with ulceration of thigh |
| PAD | Peripheral Arterial Disease/Thrombosis | ICD10 | I70.542 | Atherosclerosis of nonautologous biological bypass graft(s) of the left leg with ulceration of calf |
| PAD | Peripheral Arterial Disease/Thrombosis | ICD10 | I70.543 | Atherosclerosis of nonautologous biological bypass graft(s) of the left leg with ulceration of ankle |
| PAD | Peripheral Arterial Disease/Thrombosis | ICD10 | I70.544 | Atherosclerosis of nonautologous biological bypass graft(s) of the left leg with ulceration of heel and midfoot |
| PAD | Peripheral Arterial Disease/Thrombosis | ICD10 | I70.545 | Atherosclerosis of nonautologous biological bypass graft(s) of the left leg with ulceration of other part of foot |
| PAD | Peripheral Arterial Disease/Thrombosis | ICD10 | I70.548 | Atherosclerosis of nonautologous biological bypass graft(s) of the left leg with ulceration of other part of lower leg |
| PAD | Peripheral Arterial Disease/Thrombosis | ICD10 | I70.549 | Atherosclerosis of nonautologous biological bypass graft(s) of the left leg with ulceration of unspecified site |
| PAD | Peripheral Arterial Disease/Thrombosis | ICD10 | I70.55 | Atherosclerosis of nonautologous biological bypass graft(s) of other extremity with ulceration |

| Phenotype | phenotype description | vocabulary | code | Code description |
|---|---|---|---|---|
| PAD | Peripheral Arterial Disease/Thrombosis | ICD10 | I70.561 | Atherosclerosis of nonautologous biological bypass graft(s) of the extremities with gangrene, right leg |
| PAD | Peripheral Arterial Disease/Thrombosis | ICD10 | I70.562 | Atherosclerosis of nonautologous biological bypass graft(s) of the extremities with gangrene, left leg |
| PAD | Peripheral Arterial Disease/Thrombosis | ICD10 | I70.563 | Atherosclerosis of nonautologous biological bypass graft(s) of the extremities with gangrene, bilateral legs |
| PAD | Peripheral Arterial Disease/Thrombosis | ICD10 | I70.568 | Atherosclerosis of nonautologous biological bypass graft(s) of the extremities with gangrene, other extremity |
| PAD | Peripheral Arterial Disease/Thrombosis | ICD10 | I70.569 | Atherosclerosis of nonautologous biological bypass graft(s) of the extremities with gangrene, unspecified extremity |
| PAD | Peripheral Arterial Disease/Thrombosis | ICD10 | I70.591 | Other atherosclerosis of nonautologous biological bypass graft(s) of the extremities, right leg |
| PAD | Peripheral Arterial Disease/Thrombosis | ICD10 | I70.592 | Other atherosclerosis of nonautologous biological bypass graft(s) of the extremities, left leg |
| PAD | Peripheral Arterial Disease/Thrombosis | ICD10 | I70.593 | Other atherosclerosis of nonautologous biological bypass graft(s) of the extremities, bilateral legs |
| PAD | Peripheral Arterial Disease/Thrombosis | ICD10 | I70.598 | Other atherosclerosis of nonautologous biological bypass graft(s) of the extremities, other extremity |
| PAD | Peripheral Arterial Disease/Thrombosis | ICD10 | I70.599 | Other atherosclerosis of nonautologous biological bypass graft(s) of the extremities, unspecified extremity |

| Phenotype | phenotype description | vocabulary | code | Code description |
|---|---|---|---|---|
| PAD | Peripheral Arterial Disease/Thrombosis | ICD10 | I70.601 | Unspecified atherosclerosis of nonbiological bypass graft(s) of the extremities, right leg |
| PAD | Peripheral Arterial Disease/Thrombosis | ICD10 | I70.602 | Unspecified atherosclerosis of nonbiological bypass graft(s) of the extremities, left leg |
| PAD | Peripheral Arterial Disease/Thrombosis | ICD10 | I70.603 | Unspecified atherosclerosis of nonbiological bypass graft(s) of the extremities, bilateral legs |
| PAD | Peripheral Arterial Disease/Thrombosis | ICD10 | I70.608 | Unspecified atherosclerosis of nonbiological bypass graft(s) of the extremities, other extremity |
| PAD | Peripheral Arterial Disease/Thrombosis | ICD10 | I70.609 | Unspecified atherosclerosis of nonbiological bypass graft(s) of the extremities, unspecified extremity |
| PAD | Peripheral Arterial Disease/Thrombosis | ICD10 | I70.611 | Atherosclerosis of nonbiological bypass graft(s) of the extremities with intermittent claudication, right leg |
| PAD | Peripheral Arterial Disease/Thrombosis | ICD10 | I70.612 | Atherosclerosis of nonbiological bypass graft(s) of the extremities with intermittent claudication, left leg |
| PAD | Peripheral Arterial Disease/Thrombosis | ICD10 | I70.613 | Atherosclerosis of nonbiological bypass graft(s) of the extremities with intermittent claudication, bilateral legs |
| PAD | Peripheral Arterial Disease/Thrombosis | ICD10 | I70.618 | Atherosclerosis of nonbiological bypass graft(s) of the extremities with intermittent claudication, other extremity |
| PAD | Peripheral Arterial Disease/Thrombosis | ICD10 | I70.619 | Atherosclerosis of nonbiological bypass graft(s) of the extremities with intermittent claudication, unspecified extremity |

| Phenotype | phenotype description | vocabulary | code | Code description |
|---|---|---|---|---|
| PAD | Peripheral Arterial Disease/Thrombosis | ICD10 | I70.621 | Atherosclerosis of nonbiological bypass graft(s) of the extremities with rest pain, right leg |
| PAD | Peripheral Arterial Disease/Thrombosis | ICD10 | I70.622 | Atherosclerosis of nonbiological bypass graft(s) of the extremities with rest pain, left leg |
| PAD | Peripheral Arterial Disease/Thrombosis | ICD10 | I70.623 | Atherosclerosis of nonbiological bypass graft(s) of the extremities with rest pain, bilateral legs |
| PAD | Peripheral Arterial Disease/Thrombosis | ICD10 | I70.628 | Atherosclerosis of nonbiological bypass graft(s) of the extremities with rest pain, other extremity |
| PAD | Peripheral Arterial Disease/Thrombosis | ICD10 | I70.629 | Atherosclerosis of nonbiological bypass graft(s) of the extremities with rest pain, unspecified extremity |
| PAD | Peripheral Arterial Disease/Thrombosis | ICD10 | I70.631 | Atherosclerosis of nonbiological bypass graft(s) of the right leg with ulceration of thigh |
| PAD | Peripheral Arterial Disease/Thrombosis | ICD10 | I70.632 | Atherosclerosis of nonbiological bypass graft(s) of the right leg with ulceration of calf |
| PAD | Peripheral Arterial Disease/Thrombosis | ICD10 | I70.633 | Atherosclerosis of nonbiological bypass graft(s) of the right leg with ulceration of ankle |
| PAD | Peripheral Arterial Disease/Thrombosis | ICD10 | I70.634 | Atherosclerosis of nonbiological bypass graft(s) of the right leg with ulceration of heel and midfoot |
| PAD | Peripheral Arterial Disease/Thrombosis | ICD10 | I70.635 | Atherosclerosis of nonbiological bypass graft(s) of the right leg with ulceration of other part of foot |

| Phenotype | phenotype description | vocabulary | code | Code description |
|---|---|---|---|---|
| PAD | Peripheral Arterial Disease/Thrombosis | ICD10 | I70.638 | Atherosclerosis of nonbiological bypass graft(s) of the right leg with ulceration of other part of lower leg |
| PAD | Peripheral Arterial Disease/Thrombosis | ICD10 | I70.639 | Atherosclerosis of nonbiological bypass graft(s) of the right leg with ulceration of unspecified site |
| PAD | Peripheral Arterial Disease/Thrombosis | ICD10 | I70.641 | Atherosclerosis of nonbiological bypass graft(s) of the left leg with ulceration of thigh |
| PAD | Peripheral Arterial Disease/Thrombosis | ICD10 | I70.642 | Atherosclerosis of nonbiological bypass graft(s) of the left leg with ulceration of calf |
| PAD | Peripheral Arterial Disease/Thrombosis | ICD10 | I70.643 | Atherosclerosis of nonbiological bypass graft(s) of the left leg with ulceration of ankle |
| PAD | Peripheral Arterial Disease/Thrombosis | ICD10 | I70.644 | Atherosclerosis of nonbiological bypass graft(s) of the left leg with ulceration of heel and midfoot |
| PAD | Peripheral Arterial Disease/Thrombosis | ICD10 | I70.645 | Atherosclerosis of nonbiological bypass graft(s) of the left leg with ulceration of other part of foot |
| PAD | Peripheral Arterial Disease/Thrombosis | ICD10 | I70.648 | Atherosclerosis of nonbiological bypass graft(s) of the left leg with ulceration of other part of lower leg |
| PAD | Peripheral Arterial Disease/Thrombosis | ICD10 | I70.649 | Atherosclerosis of nonbiological bypass graft(s) of the left leg with ulceration of unspecified site |
| PAD | Peripheral Arterial Disease/Thrombosis | ICD10 | I70.65 | Atherosclerosis of nonbiological bypass graft(s) of other extremity with ulceration |

| Phenotype | phenotype description | vocabulary | code | Code description |
|---|---|---|---|---|
| PAD | Peripheral Arterial Disease/Thrombosis | ICD10 | I70.661 | Atherosclerosis of nonbiological bypass graft(s) of the extremities with gangrene, right leg |
| PAD | Peripheral Arterial Disease/Thrombosis | ICD10 | I70.662 | Atherosclerosis of nonbiological bypass graft(s) of the extremities with gangrene, left leg |
| PAD | Peripheral Arterial Disease/Thrombosis | ICD10 | I70.663 | Atherosclerosis of nonbiological bypass graft(s) of the extremities with gangrene, bilateral legs |
| PAD | Peripheral Arterial Disease/Thrombosis | ICD10 | I70.668 | Atherosclerosis of nonbiological bypass graft(s) of the extremities with gangrene, other extremity |
| PAD | Peripheral Arterial Disease/Thrombosis | ICD10 | I70.669 | Atherosclerosis of nonbiological bypass graft(s) of the extremities with gangrene, unspecified extremity |
| PAD | Peripheral Arterial Disease/Thrombosis | ICD10 | I70.691 | Other atherosclerosis of nonbiological bypass graft(s) of the extremities, right leg |
| PAD | Peripheral Arterial Disease/Thrombosis | ICD10 | I70.692 | Other atherosclerosis of nonbiological bypass graft(s) of the extremities, left leg |
| PAD | Peripheral Arterial Disease/Thrombosis | ICD10 | I70.693 | Other atherosclerosis of nonbiological bypass graft(s) of the extremities, bilateral legs |
| PAD | Peripheral Arterial Disease/Thrombosis | ICD10 | I70.698 | Other atherosclerosis of nonbiological bypass graft(s) of the extremities, other extremity |
| PAD | Peripheral Arterial Disease/Thrombosis | ICD10 | I70.699 | Other atherosclerosis of nonbiological bypass graft(s) of the extremities, unspecified extremity |

| Phenotype | phenotype description | vocabulary | code | Code description |
|---|---|---|---|---|
| PAD | Peripheral Arterial Disease/Thrombosis | ICD10 | I70.701 | Unspecified atherosclerosis of other type of bypass graft(s) of the extremities, right leg |
| PAD | Peripheral Arterial Disease/Thrombosis | ICD10 | I70.702 | Unspecified atherosclerosis of other type of bypass graft(s) of the extremities, left leg |
| PAD | Peripheral Arterial Disease/Thrombosis | ICD10 | I70.703 | Unspecified atherosclerosis of other type of bypass graft(s) of the extremities, bilateral legs |
| PAD | Peripheral Arterial Disease/Thrombosis | ICD10 | I70.708 | Unspecified atherosclerosis of other type of bypass graft(s) of the extremities, other extremity |
| PAD | Peripheral Arterial Disease/Thrombosis | ICD10 | I70.709 | Unspecified atherosclerosis of other type of bypass graft(s) of the extremities, unspecified extremity |
| PAD | Peripheral Arterial Disease/Thrombosis | ICD10 | I70.711 | Atherosclerosis of other type of bypass graft(s) of the extremities with intermittent claudication, right leg |
| PAD | Peripheral Arterial Disease/Thrombosis | ICD10 | I70.712 | Atherosclerosis of other type of bypass graft(s) of the extremities with intermittent claudication, left leg |
| PAD | Peripheral Arterial Disease/Thrombosis | ICD10 | I70.713 | Atherosclerosis of other type of bypass graft(s) of the extremities with intermittent claudication, bilateral legs |
| PAD | Peripheral Arterial Disease/Thrombosis | ICD10 | I70.718 | Atherosclerosis of other type of bypass graft(s) of the extremities with intermittent claudication, other extremity |
| PAD | Peripheral Arterial Disease/Thrombosis | ICD10 | I70.719 | Atherosclerosis of other type of bypass graft(s) of the extremities with intermittent claudication, unspecified extremity |

| Phenotype | phenotype description | vocabulary | code | Code description |
|---|---|---|---|---|
| PAD | Peripheral Arterial Disease/Thrombosis | ICD10 | I70.721 | Atherosclerosis of other type of bypass graft(s) of the extremities with rest pain, right leg |
| PAD | Peripheral Arterial Disease/Thrombosis | ICD10 | I70.722 | Atherosclerosis of other type of bypass graft(s) of the extremities with rest pain, left leg |
| PAD | Peripheral Arterial Disease/Thrombosis | ICD10 | I70.723 | Atherosclerosis of other type of bypass graft(s) of the extremities with rest pain, bilateral legs |
| PAD | Peripheral Arterial Disease/Thrombosis | ICD10 | I70.728 | Atherosclerosis of other type of bypass graft(s) of the extremities with rest pain, other extremity |
| PAD | Peripheral Arterial Disease/Thrombosis | ICD10 | I70.729 | Atherosclerosis of other type of bypass graft(s) of the extremities with rest pain, unspecified extremity |
| PAD | Peripheral Arterial Disease/Thrombosis | ICD10 | I70.731 | Atherosclerosis of other type of bypass graft(s) of the right leg with ulceration of thigh |
| PAD | Peripheral Arterial Disease/Thrombosis | ICD10 | I70.732 | Atherosclerosis of other type of bypass graft(s) of the right leg with ulceration of calf |
| PAD | Peripheral Arterial Disease/Thrombosis | ICD10 | I70.733 | Atherosclerosis of other type of bypass graft(s) of the right leg with ulceration of ankle |
| PAD | Peripheral Arterial Disease/Thrombosis | ICD10 | I70.734 | Atherosclerosis of other type of bypass graft(s) of the right leg with ulceration of heel and midfoot |
| PAD | Peripheral Arterial Disease/Thrombosis | ICD10 | I70.735 | Atherosclerosis of other type of bypass graft(s) of the right leg with ulceration of other part of foot |

| Phenotype | phenotype description | vocabulary | code | Code description |
|---|---|---|---|---|
| PAD | Peripheral Arterial Disease/Thrombosis | ICD10 | I70.738 | Atherosclerosis of other type of bypass graft(s) of the right leg with ulceration of other part of lower leg |
| PAD | Peripheral Arterial Disease/Thrombosis | ICD10 | I70.739 | Atherosclerosis of other type of bypass graft(s) of the right leg with ulceration of unspecified site |
| PAD | Peripheral Arterial Disease/Thrombosis | ICD10 | I70.741 | Atherosclerosis of other type of bypass graft(s) of the left leg with ulceration of thigh |
| PAD | Peripheral Arterial Disease/Thrombosis | ICD10 | I70.742 | Atherosclerosis of other type of bypass graft(s) of the left leg with ulceration of calf |
| PAD | Peripheral Arterial Disease/Thrombosis | ICD10 | I70.743 | Atherosclerosis of other type of bypass graft(s) of the left leg with ulceration of ankle |
| PAD | Peripheral Arterial Disease/Thrombosis | ICD10 | I70.744 | Atherosclerosis of other type of bypass graft(s) of the left leg with ulceration of heel and midfoot |
| PAD | Peripheral Arterial Disease/Thrombosis | ICD10 | I70.745 | Atherosclerosis of other type of bypass graft(s) of the left leg with ulceration of other part of foot |
| PAD | Peripheral Arterial Disease/Thrombosis | ICD10 | I70.748 | Atherosclerosis of other type of bypass graft(s) of the left leg with ulceration of other part of lower leg |
| PAD | Peripheral Arterial Disease/Thrombosis | ICD10 | I70.749 | Atherosclerosis of other type of bypass graft(s) of the left leg with ulceration of unspecified site |
| PAD | Peripheral Arterial Disease/Thrombosis | ICD10 | I70.75 | Atherosclerosis of other type of bypass graft(s) of other extremity with ulceration |

| Phenotype | phenotype description | vocabulary | code | Code description |
| --- | --- | --- | --- | --- |
| PAD | Peripheral Arterial Disease/Thrombosis | ICD10 | I70.761 | Atherosclerosis of other type of bypass graft(s) of the extremities with gangrene, right leg |
| PAD | Peripheral Arterial Disease/Thrombosis | ICD10 | I70.762 | Atherosclerosis of other type of bypass graft(s) of the extremities with gangrene, left leg |
| PAD | Peripheral Arterial Disease/Thrombosis | ICD10 | I70.763 | Atherosclerosis of other type of bypass graft(s) of the extremities with gangrene, bilateral legs |
| PAD | Peripheral Arterial Disease/Thrombosis | ICD10 | I70.768 | Atherosclerosis of other type of bypass graft(s) of the extremities with gangrene, other extremity |
| PAD | Peripheral Arterial Disease/Thrombosis | ICD10 | I70.769 | Atherosclerosis of other type of bypass graft(s) of the extremities with gangrene, unspecified extremity |
| PAD | Peripheral Arterial Disease/Thrombosis | ICD10 | I70.791 | Other atherosclerosis of other type of bypass graft(s) of the extremities, right leg |
| PAD | Peripheral Arterial Disease/Thrombosis | ICD10 | I70.792 | Other atherosclerosis of other type of bypass graft(s) of the extremities, left leg |
| PAD | Peripheral Arterial Disease/Thrombosis | ICD10 | I70.793 | Other atherosclerosis of other type of bypass graft(s) of the extremities, bilateral legs |
| PAD | Peripheral Arterial Disease/Thrombosis | ICD10 | I70.798 | Other atherosclerosis of other type of bypass graft(s) of the extremities, other extremity |
| PAD | Peripheral Arterial Disease/Thrombosis | ICD10 | I70.799 | Other atherosclerosis of other type of bypass graft(s) of the extremities, unspecified extremity |

| Phenotype | phenotype description | vocabulary | code | Code description |
|---|---|---|---|---|
| PAD | Peripheral Arterial Disease/Thrombosis | ICD10 | I70.90 | Unspecified atherosclerosis |
| PAD | Peripheral Arterial Disease/Thrombosis | ICD10 | I70.91 | Generalized atherosclerosis |
| PAD | Peripheral Arterial Disease/Thrombosis | ICD10 | I70.92 | Chronic total occlusion of artery of the extremities |
| PAD | Peripheral Arterial Disease/Thrombosis | ICD10 | I73.9 | Peripheral vascular disease, unspecified |
| PAD | Peripheral Arterial Disease/Thrombosis | ICD10 | I74.01 | Saddle embolus of abdominal aorta |
| PAD | Peripheral Arterial Disease/Thrombosis | ICD10 | I74.09 | Other arterial embolism and thrombosis of abdominal aorta |
| PAD | Peripheral Arterial Disease/Thrombosis | ICD10 | I74.10 | Embolism and thrombosis of unspecified parts of aorta |
| PAD | Peripheral Arterial Disease/Thrombosis | ICD10 | I74.19 | Embolism and thrombosis of other parts of aorta |
| PAD | Peripheral Arterial Disease/Thrombosis | ICD10 | I74.3 | Embolism and thrombosis of arteries of the lower extremities |
| PAD | Peripheral Arterial Disease/Thrombosis | ICD10 | I74.4 | Embolism and thrombosis of arteries of extremities, unspecified |

| Phenotype | phenotype description | vocabulary | code | Code description |
|---|---|---|---|---|
| PAD | Peripheral Arterial Disease/Thrombosis | ICD10 | I74.5 | Embolism and thrombosis of iliac artery |
| PAD | Peripheral Arterial Disease/Thrombosis | ICD10 | I74.8 | Embolism and thrombosis of other arteries |
| PAD | Peripheral Arterial Disease/Thrombosis | ICD10 | I75.021 | Atheroembolism of right lower extremity |
| PAD | Peripheral Arterial Disease/Thrombosis | ICD10 | I75.022 | Atheroembolism of left lower extremity |
| PAD | Peripheral Arterial Disease/Thrombosis | ICD10 | I75.023 | Atheroembolism of bilateral lower extremities |
| PAD | Peripheral Arterial Disease/Thrombosis | ICD10 | I75.029 | Atheroembolism of unspecified lower extremity |
| PAD | Peripheral Arterial Disease/Thrombosis | ICD10 | I77.1 | Stricture of artery |
| PAD | Peripheral Arterial Disease/Thrombosis | ICD10 | I96. | Gangrene, not elsewhere classified |
| PAD | Peripheral Arterial Disease/Thrombosis | ICD10 | L97.101 | Non-pressure chronic ulcer of unspecified thigh limited to breakdown of skin |
| PAD | Peripheral Arterial Disease/Thrombosis | ICD10 | L97.102 | Non-pressure chronic ulcer of unspecified thigh with fat layer exposed |

| Phenotype | phenotype description | vocabulary | code | Code description |
|---|---|---|---|---|
| PAD | Peripheral Arterial Disease/Thrombosis | ICD10 | L97.103 | Non-pressure chronic ulcer of unspecified thigh with necrosis of muscle |
| PAD | Peripheral Arterial Disease/Thrombosis | ICD10 | L97.104 | Non-pressure chronic ulcer of unspecified thigh with necrosis of bone |
| PAD | Peripheral Arterial Disease/Thrombosis | ICD10 | L97.109 | Non-pressure chronic ulcer of unspecified thigh with unspecified severity |
| PAD | Peripheral Arterial Disease/Thrombosis | ICD10 | L97.111 | Non-pressure chronic ulcer of right thigh limited to breakdown of skin |
| PAD | Peripheral Arterial Disease/Thrombosis | ICD10 | L97.112 | Non-pressure chronic ulcer of right thigh with fat layer exposed |
| PAD | Peripheral Arterial Disease/Thrombosis | ICD10 | L97.113 | Non-pressure chronic ulcer of right thigh with necrosis of muscle |
| PAD | Peripheral Arterial Disease/Thrombosis | ICD10 | L97.114 | Non-pressure chronic ulcer of right thigh with necrosis of bone |
| PAD | Peripheral Arterial Disease/Thrombosis | ICD10 | L97.119 | Non-pressure chronic ulcer of right thigh with unspecified severity |
| PAD | Peripheral Arterial Disease/Thrombosis | ICD10 | L97.121 | Non-pressure chronic ulcer of left thigh limited to breakdown of skin |
| PAD | Peripheral Arterial Disease/Thrombosis | ICD10 | L97.122 | Non-pressure chronic ulcer of left thigh with fat layer exposed |

| Phenotype | phenotype description | vocabulary | code | Code description |
|---|---|---|---|---|
| PAD | Peripheral Arterial Disease/Thrombosis | ICD10 | L97.123 | Non-pressure chronic ulcer of left thigh with necrosis of muscle |
| PAD | Peripheral Arterial Disease/Thrombosis | ICD10 | L97.124 | Non-pressure chronic ulcer of left thigh with necrosis of bone |
| PAD | Peripheral Arterial Disease/Thrombosis | ICD10 | L97.129 | Non-pressure chronic ulcer of left thigh with unspecified severity |
| PAD | Peripheral Arterial Disease/Thrombosis | ICD10 | L97.201 | Non-pressure chronic ulcer of unspecified calf limited to breakdown of skin |
| PAD | Peripheral Arterial Disease/Thrombosis | ICD10 | L97.202 | Non-pressure chronic ulcer of unspecified calf with fat layer exposed |
| PAD | Peripheral Arterial Disease/Thrombosis | ICD10 | L97.203 | Non-pressure chronic ulcer of unspecified calf with necrosis of muscle |
| PAD | Peripheral Arterial Disease/Thrombosis | ICD10 | L97.204 | Non-pressure chronic ulcer of unspecified calf with necrosis of bone |
| PAD | Peripheral Arterial Disease/Thrombosis | ICD10 | L97.209 | Non-pressure chronic ulcer of unspecified calf with unspecified severity |
| PAD | Peripheral Arterial Disease/Thrombosis | ICD10 | L97.211 | Non-pressure chronic ulcer of right calf limited to breakdown of skin |
| PAD | Peripheral Arterial Disease/Thrombosis | ICD10 | L97.212 | Non-pressure chronic ulcer of right calf with fat layer exposed |

| Phenotype | phenotype description | vocabulary | code | Code description |
|---|---|---|---|---|
| PAD | Peripheral Arterial Disease/Thrombosis | ICD10 | L97.213 | Non-pressure chronic ulcer of right calf with necrosis of muscle |
| PAD | Peripheral Arterial Disease/Thrombosis | ICD10 | L97.214 | Non-pressure chronic ulcer of right calf with necrosis of bone |
| PAD | Peripheral Arterial Disease/Thrombosis | ICD10 | L97.219 | Non-pressure chronic ulcer of right calf with unspecified severity |
| PAD | Peripheral Arterial Disease/Thrombosis | ICD10 | L97.221 | Non-pressure chronic ulcer of left calf limited to breakdown of skin |
| PAD | Peripheral Arterial Disease/Thrombosis | ICD10 | L97.222 | Non-pressure chronic ulcer of left calf with fat layer exposed |
| PAD | Peripheral Arterial Disease/Thrombosis | ICD10 | L97.223 | Non-pressure chronic ulcer of left calf with necrosis of muscle |
| PAD | Peripheral Arterial Disease/Thrombosis | ICD10 | L97.224 | Non-pressure chronic ulcer of left calf with necrosis of bone |
| PAD | Peripheral Arterial Disease/Thrombosis | ICD10 | L97.229 | Non-pressure chronic ulcer of left calf with unspecified severity |
| PAD | Peripheral Arterial Disease/Thrombosis | ICD10 | L97.301 | Non-pressure chronic ulcer of unspecified ankle limited to breakdown of skin |
| PAD | Peripheral Arterial Disease/Thrombosis | ICD10 | L97.302 | Non-pressure chronic ulcer of unspecified ankle with fat layer exposed |

| Phenotype | phenotype description | vocabul ary | code | Code description |
|---|---|---|---|---|
| PAD | Peripheral Arterial Disease/Thrombosis | ICD10 | L97.303 | Non-pressure chronic ulcer of unspecified ankle with necrosis of muscle |
| PAD | Peripheral Arterial Disease/Thrombosis | ICD10 | L97.304 | Non-pressure chronic ulcer of unspecified ankle with necrosis of bone |
| PAD | Peripheral Arterial Disease/Thrombosis | ICD10 | L97.309 | Non-pressure chronic ulcer of unspecified ankle with unspecified severity |
| PAD | Peripheral Arterial Disease/Thrombosis | ICD10 | L97.311 | Non-pressure chronic ulcer of right ankle limited to breakdown of skin |
| PAD | Peripheral Arterial Disease/Thrombosis | ICD10 | L97.312 | Non-pressure chronic ulcer of right ankle with fat layer exposed |
| PAD | Peripheral Arterial Disease/Thrombosis | ICD10 | L97.313 | Non-pressure chronic ulcer of right ankle with necrosis of muscle |
| PAD | Peripheral Arterial Disease/Thrombosis | ICD10 | L97.314 | Non-pressure chronic ulcer of right ankle with necrosis of bone |
| PAD | Peripheral Arterial Disease/Thrombosis | ICD10 | L97.319 | Non-pressure chronic ulcer of right ankle with unspecified severity |
| PAD | Peripheral Arterial Disease/Thrombosis | ICD10 | L97.321 | Non-pressure chronic ulcer of left ankle limited to breakdown of skin |
| PAD | Peripheral Arterial Disease/Thrombosis | ICD10 | L97.322 | Non-pressure chronic ulcer of left ankle with fat layer exposed |

| Phenotype | phenotype description | vocabulary | code | Code description |
|---|---|---|---|---|
| PAD | Peripheral Arterial Disease/Thrombosis | ICD10 | L97.323 | Non-pressure chronic ulcer of left ankle with necrosis of muscle |
| PAD | Peripheral Arterial Disease/Thrombosis | ICD10 | L97.324 | Non-pressure chronic ulcer of left ankle with necrosis of bone |
| PAD | Peripheral Arterial Disease/Thrombosis | ICD10 | L97.329 | Non-pressure chronic ulcer of left ankle with unspecified severity |
| PAD | Peripheral Arterial Disease/Thrombosis | ICD10 | L97.401 | Non-pressure chronic ulcer of unspecified heel and midfoot limited to breakdown of skin |
| PAD | Peripheral Arterial Disease/Thrombosis | ICD10 | L97.402 | Non-pressure chronic ulcer of unspecified heel and midfoot with fat layer exposed |
| PAD | Peripheral Arterial Disease/Thrombosis | ICD10 | L97.403 | Non-pressure chronic ulcer of unspecified heel and midfoot with necrosis of muscle |
| PAD | Peripheral Arterial Disease/Thrombosis | ICD10 | L97.404 | Non-pressure chronic ulcer of unspecified heel and midfoot with necrosis of bone |
| PAD | Peripheral Arterial Disease/Thrombosis | ICD10 | L97.409 | Non-pressure chronic ulcer of unspecified heel and midfoot with unspecified severity |
| PAD | Peripheral Arterial Disease/Thrombosis | ICD10 | L97.411 | Non-pressure chronic ulcer of right heel and midfoot limited to breakdown of skin |
| PAD | Peripheral Arterial Disease/Thrombosis | ICD10 | L97.412 | Non-pressure chronic ulcer of right heel and midfoot with fat layer exposed |

| Phenotype | phenotype description | vocabulary | code | Code description |
|---|---|---|---|---|
| PAD | Peripheral Arterial Disease/Thrombosis | ICD10 | L97.413 | Non-pressure chronic ulcer of right heel and midfoot with necrosis of muscle |
| PAD | Peripheral Arterial Disease/Thrombosis | ICD10 | L97.414 | Non-pressure chronic ulcer of right heel and midfoot with necrosis of bone |
| PAD | Peripheral Arterial Disease/Thrombosis | ICD10 | L97.419 | Non-pressure chronic ulcer of right heel and midfoot with unspecified severity |
| PAD | Peripheral Arterial Disease/Thrombosis | ICD10 | L97.421 | Non-pressure chronic ulcer of left heel and midfoot limited to breakdown of skin |
| PAD | Peripheral Arterial Disease/Thrombosis | ICD10 | L97.422 | Non-pressure chronic ulcer of left heel and midfoot with fat layer exposed |
| PAD | Peripheral Arterial Disease/Thrombosis | ICD10 | L97.423 | Non-pressure chronic ulcer of left heel and midfoot with necrosis of muscle |
| PAD | Peripheral Arterial Disease/Thrombosis | ICD10 | L97.424 | Non-pressure chronic ulcer of left heel and midfoot with necrosis of bone |
| PAD | Peripheral Arterial Disease/Thrombosis | ICD10 | L97.429 | Non-pressure chronic ulcer of left heel and midfoot with unspecified severity |
| PAD | Peripheral Arterial Disease/Thrombosis | ICD10 | L97.501 | Non-pressure chronic ulcer of other part of unspecified foot limited to breakdown of skin |
| PAD | Peripheral Arterial Disease/Thrombosis | ICD10 | L97.502 | Non-pressure chronic ulcer of other part of unspecified foot with fat layer exposed |

| Phenotype | phenotype description | vocabulary | code | Code description |
|---|---|---|---|---|
| PAD | Peripheral Arterial Disease/Thrombosis | ICD10 | L97.503 | Non-pressure chronic ulcer of other part of unspecified foot with necrosis of muscle |
| PAD | Peripheral Arterial Disease/Thrombosis | ICD10 | L97.504 | Non-pressure chronic ulcer of other part of unspecified foot with necrosis of bone |
| PAD | Peripheral Arterial Disease/Thrombosis | ICD10 | L97.509 | Non-pressure chronic ulcer of other part of unspecified foot with unspecified severity |
| PAD | Peripheral Arterial Disease/Thrombosis | ICD10 | L97.511 | Non-pressure chronic ulcer of other part of right foot limited to breakdown of skin |
| PAD | Peripheral Arterial Disease/Thrombosis | ICD10 | L97.512 | Non-pressure chronic ulcer of other part of right foot with fat layer exposed |
| PAD | Peripheral Arterial Disease/Thrombosis | ICD10 | L97.513 | Non-pressure chronic ulcer of other part of right foot with necrosis of muscle |
| PAD | Peripheral Arterial Disease/Thrombosis | ICD10 | L97.514 | Non-pressure chronic ulcer of other part of right foot with necrosis of bone |
| PAD | Peripheral Arterial Disease/Thrombosis | ICD10 | L97.519 | Non-pressure chronic ulcer of other part of right foot with unspecified severity |
| PAD | Peripheral Arterial Disease/Thrombosis | ICD10 | L97.521 | Non-pressure chronic ulcer of other part of left foot limited to breakdown of skin |
| PAD | Peripheral Arterial Disease/Thrombosis | ICD10 | L97.522 | Non-pressure chronic ulcer of other part of left foot with fat layer exposed |

| Phenotype | phenotype description | vocabulary | code | Code description |
|---|---|---|---|---|
| PAD | Peripheral Arterial Disease/Thrombosis | ICD10 | L97.523 | Non-pressure chronic ulcer of other part of left foot with necrosis of muscle |
| PAD | Peripheral Arterial Disease/Thrombosis | ICD10 | L97.524 | Non-pressure chronic ulcer of other part of left foot with necrosis of bone |
| PAD | Peripheral Arterial Disease/Thrombosis | ICD10 | L97.529 | Non-pressure chronic ulcer of other part of left foot with unspecified severity |
| PAD | Peripheral Arterial Disease/Thrombosis | ICD10 | L97.801 | Non-pressure chronic ulcer of other part of unspecified lower leg limited to breakdown of skin |
| PAD | Peripheral Arterial Disease/Thrombosis | ICD10 | L97.802 | Non-pressure chronic ulcer of other part of unspecified lower leg with fat layer exposed |
| PAD | Peripheral Arterial Disease/Thrombosis | ICD10 | L97.803 | Non-pressure chronic ulcer of other part of unspecified lower leg with necrosis of muscle |
| PAD | Peripheral Arterial Disease/Thrombosis | ICD10 | L97.804 | Non-pressure chronic ulcer of other part of unspecified lower leg with necrosis of bone |
| PAD | Peripheral Arterial Disease/Thrombosis | ICD10 | L97.809 | Non-pressure chronic ulcer of other part of unspecified lower leg with unspecified severity |
| PAD | Peripheral Arterial Disease/Thrombosis | ICD10 | L97.811 | Non-pressure chronic ulcer of other part of right lower leg limited to breakdown of skin |
| PAD | Peripheral Arterial Disease/Thrombosis | ICD10 | L97.812 | Non-pressure chronic ulcer of other part of right lower leg with fat layer exposed |

| Phenotype | phenotype description | vocabulary | code | Code description |
|---|---|---|---|---|
| PAD | Peripheral Arterial Disease/Thrombosis | ICD10 | L97.813 | Non-pressure chronic ulcer of other part of right lower leg with necrosis of muscle |
| PAD | Peripheral Arterial Disease/Thrombosis | ICD10 | L97.814 | Non-pressure chronic ulcer of other part of right lower leg with necrosis of bone |
| PAD | Peripheral Arterial Disease/Thrombosis | ICD10 | L97.819 | Non-pressure chronic ulcer of other part of right lower leg with unspecified severity |
| PAD | Peripheral Arterial Disease/Thrombosis | ICD10 | L97.821 | Non-pressure chronic ulcer of other part of left lower leg limited to breakdown of skin |
| PAD | Peripheral Arterial Disease/Thrombosis | ICD10 | L97.822 | Non-pressure chronic ulcer of other part of left lower leg with fat layer exposed |
| PAD | Peripheral Arterial Disease/Thrombosis | ICD10 | L97.823 | Non-pressure chronic ulcer of other part of left lower leg with necrosis of muscle |
| PAD | Peripheral Arterial Disease/Thrombosis | ICD10 | L97.824 | Non-pressure chronic ulcer of other part of left lower leg with necrosis of bone |
| PAD | Peripheral Arterial Disease/Thrombosis | ICD10 | L97.829 | Non-pressure chronic ulcer of other part of left lower leg with unspecified severity |
| PAD | Peripheral Arterial Disease/Thrombosis | ICD10 | L97.901 | Non-pressure chronic ulcer of unspecified part of unspecified lower leg limited to breakdown of skin |
| PAD | Peripheral Arterial Disease/Thrombosis | ICD10 | L97.902 | Non-pressure chronic ulcer of unspecified part of unspecified lower leg with fat layer exposed |

| Phenotype | phenotype description | vocabulary | code | Code description |
|---|---|---|---|---|
| PAD | Peripheral Arterial Disease/Thrombosis | ICD10 | L97.903 | Non-pressure chronic ulcer of unspecified part of unspecified lower leg with necrosis of muscle |
| PAD | Peripheral Arterial Disease/Thrombosis | ICD10 | L97.904 | Non-pressure chronic ulcer of unspecified part of unspecified lower leg with necrosis of bone |
| PAD | Peripheral Arterial Disease/Thrombosis | ICD10 | L97.909 | Non-pressure chronic ulcer of unspecified part of unspecified lower leg with unspecified severity |
| PAD | Peripheral Arterial Disease/Thrombosis | ICD10 | L97.911 | Non-pressure chronic ulcer of unspecified part of right lower leg limited to breakdown of skin |
| PAD | Peripheral Arterial Disease/Thrombosis | ICD10 | L97.912 | Non-pressure chronic ulcer of unspecified part of right lower leg with fat layer exposed |
| PAD | Peripheral Arterial Disease/Thrombosis | ICD10 | L97.913 | Non-pressure chronic ulcer of unspecified part of right lower leg with necrosis of muscle |
| PAD | Peripheral Arterial Disease/Thrombosis | ICD10 | L97.914 | Non-pressure chronic ulcer of unspecified part of right lower leg with necrosis of bone |
| PAD | Peripheral Arterial Disease/Thrombosis | ICD10 | L97.919 | Non-pressure chronic ulcer of unspecified part of right lower leg with unspecified severity |
| PAD | Peripheral Arterial Disease/Thrombosis | ICD10 | L97.921 | Non-pressure chronic ulcer of unspecified part of left lower leg limited to breakdown of skin |
| PAD | Peripheral Arterial Disease/Thrombosis | ICD10 | L97.922 | Non-pressure chronic ulcer of unspecified part of left lower leg with fat layer exposed |

| Phenotype | phenotype description | vocabulary | code | Code description |
|---|---|---|---|---|
| PAD | Peripheral Arterial Disease/Thrombosis | ICD10 | L97.923 | Non-pressure chronic ulcer of unspecified part of left lower leg with necrosis of muscle |
| PAD | Peripheral Arterial Disease/Thrombosis | ICD10 | L97.924 | Non-pressure chronic ulcer of unspecified part of left lower leg with necrosis of bone |
| PAD | Peripheral Arterial Disease/Thrombosis | ICD10 | L97.929 | Non-pressure chronic ulcer of unspecified part of left lower leg with unspecified severity |
| PAD | Peripheral Arterial Disease/Thrombosis | ICD9 | 250.71 | DIABETES WITH PERIPHERAL CIRCULATORY DISORDERS, TYPE I [JUVENILE TYPE], NOT STATED AS UNCONTROLLED |
| PAD | Peripheral Arterial Disease/Thrombosis | ICD9 | 250.72 | DIABETES WITH PERIPHERAL CIRCULATORY DISORDERS, TYPE II OR UNSPECIFIED TYPE, UNCONTROLLED |
| PAD | Peripheral Arterial Disease/Thrombosis | ICD9 | 250.73 | DIABETES WITH PERIPHERAL CIRCULATORY DISORDERS, TYPE I [JUVENILE TYPE], UNCONTROLLED |
| PAD | Peripheral Arterial Disease/Thrombosis | ICD9 | 440.2 | ATHEROSCLEROSIS OF ARTERIES OF THE EXTREMITIES |
| PAD | Peripheral Arterial Disease/Thrombosis | ICD9 | 440.21 | ATHEROSCLEROSIS OF NATIVE ARTERIES OF EXTREMITIES W/INTERMITTENT CLAUDICATION |
| PAD | Peripheral Arterial Disease/Thrombosis | ICD9 | 440.22 | ATHEROSCLEROSIS OF NATIVE ARTERIES OF EXTREMITIES WITH REST PAIN |
| PAD | Peripheral Arterial Disease/Thrombosis | ICD9 | 440.23 | ATHEROSCLEROSIS OF NATIVE ARTERIES OF EXTREMITIES W/ ULCERATION |

| Phenotype | phenotype description | vocabulary | code | Code description |
|---|---|---|---|---|
| PAD | Peripheral Arterial Disease/Thrombosis | ICD9 | 440.24 | ATHEROSCLEROSIS OF NATIVE ARTERIES OF EXTREMITIES W/ GANGRENE |
| PAD | Peripheral Arterial Disease/Thrombosis | ICD9 | 440.29 | OTHER ATHEROSCLEROSIS OF NATIVE ARTERIES OF EXTREMITIES |
| PAD | Peripheral Arterial Disease/Thrombosis | ICD9 | 440.31 | ATHEROSCLEROSIS OF AUTOLOGOUS VEIN BYPASS GRAFT OF THE EXTREMITIES |
| PAD | Peripheral Arterial Disease/Thrombosis | ICD9 | 440.32 | ATHEROSCLEROSIS OF NONAUTOLOGOUS BIOLOGICAL BYPASS GRAFT OF THE EXTREMITIES |
| PAD | Peripheral Arterial Disease/Thrombosis | ICD9 | 440.4 | CHRONIC TOTAL OCCLUSION OF ARTERY OF THE EXTREMITIES |
| PAD | Peripheral Arterial Disease/Thrombosis | ICD9 | 440.9 | GENERALIZED AND UNSPECIFIED ATHEROSCLEROSIS |
| PAD | Peripheral Arterial Disease/Thrombosis | ICD9 | 443.9 | PERIPHERAL VASCULAR DISEASE, UNSPECIFIED |
| PAD | Peripheral Arterial Disease/Thrombosis | ICD9 | 444.22 | ARTERIAL EMBOLISM AND THROMBOSIS OF LOWER EXTREMITY |
| PAD | Peripheral Arterial Disease/Thrombosis | ICD9 | 444.81 | EMBOLISM AND THROMBOSIS OF ILIAC ARTERY |
| PAD | Peripheral Arterial Disease/Thrombosis | ICD9 | 444.89 | EMBOLISM AND THROMBOSIS OF OTHER ARTERY |

| Phenotype | phenotype description | vocabulary | code | Code description |
|---|---|---|---|---|
| PAD | Peripheral Arterial Disease/Thrombosis | ICD9 | 445.02 | ATHEROEMBOLISM LOWER EXTREMITY |
| PAD | Peripheral Arterial Disease/Thrombosis | ICD9 | 447.1 | STRICTURE OF ARTERY |
| PAD | Peripheral Arterial Disease/Thrombosis | ICD9 | 459.9 | UNSPECIFIED CIRCULATORY SYSTEM DISORDER |
| PAD | Peripheral Arterial Disease/Thrombosis | ICD9 | 707.1 | ULCER OF LOWER LIMBS, EXCEPT DECUBITUS ULCER |
| PAD | Peripheral Arterial Disease/Thrombosis | ICD9 | 707.11 | ULCER OF THIGH |
| PAD | Peripheral Arterial Disease/Thrombosis | ICD9 | 707.12 | ULCER OF CALF |
| PAD | Peripheral Arterial Disease/Thrombosis | ICD9 | 707.13 | ULCER OF ANKLE |
| PAD | Peripheral Arterial Disease/Thrombosis | ICD9 | 707.14 | ULCER OF HEEL AND MIDFOOT |
| PAD | Peripheral Arterial Disease/Thrombosis | ICD9 | 707.15 | ULCER OF OTHER PART OF FOOT |
| PAD | Peripheral Arterial Disease/Thrombosis | ICD9 | 707.19 | ULCER OF OTHER PART OF LOWER LIMB |

| Phenotype | phenotype description | vocabulary | code | Code description |
|---|---|---|---|---|
| PAD | Peripheral Arterial Disease/Thrombosis | ICD9 | 785.4 | GANGRENE |
| VascDem | Vascular dementia | ICD10 | F01.50 | Vascular dementia without behavioral disturbance |
| VascDem | Vascular dementia | ICD10 | F01.51 | Vascular dementia with behavioral disturbance |
| VascDem | Vascular dementia | ICD9 | 290.4 | VASCULAR DEMENTIA, UNCOMPLICATED |
| VascDem | Vascular dementia | ICD9 | 290.41 | VASCULAR DEMENTIA, WITH DELIRIUM |
| VascDem | Vascular dementia | ICD9 | 290.42 | VASCULAR DEMENTIA, WITH DELUSIONS |
| VascDem | Vascular dementia | ICD9 | 290.43 | VASCULAR DEMENTIA, WITH DEPRESSED MOOD |
| VTE | Venous Thromboembolism (DVT or PE) | ICD10 | I26.01 | Septic pulmonary embolism with acute cor pulmonale |
| VTE | Venous Thromboembolism (DVT or PE) | ICD10 | I26.02 | Saddle embolus of pulmonary artery with acute cor pulmonale |
| VTE | Venous Thromboembolism (DVT or PE) | ICD10 | I26.09 | Other pulmonary embolism with acute cor pulmonale |

| Phenotype | phenotype description | vocabulary | code | Code description |
|---|---|---|---|---|
| VTE | Venous Thromboembolism (DVT or PE) | ICD10 | I26.90 | Septic pulmonary embolism without acute cor pulmonale |
| VTE | Venous Thromboembolism (DVT or PE) | ICD10 | I26.92 | Saddle embolus of pulmonary artery without acute cor pulmonale |
| VTE | Venous Thromboembolism (DVT or PE) | ICD10 | I26.93 | Single subsegmental pulmonary embolism without acute cor pulmonale |
| VTE | Venous Thromboembolism (DVT or PE) | ICD10 | I26.94 | Multiple subsegmental pulmonary emboli without acute cor pulmonale |
| VTE | Venous Thromboembolism (DVT or PE) | ICD10 | I26.99 | Other pulmonary embolism without acute cor pulmonale |
| VTE | Venous Thromboembolism (DVT or PE) | ICD10 | I80.10 | Phlebitis and thrombophlebitis of unspecified femoral vein |
| VTE | Venous Thromboembolism (DVT or PE) | ICD10 | I80.11 | Phlebitis and thrombophlebitis of right femoral vein |
| VTE | Venous Thromboembolism (DVT or PE) | ICD10 | I80.12 | Phlebitis and thrombophlebitis of left femoral vein |
| VTE | Venous Thromboembolism (DVT or PE) | ICD10 | I80.13 | Phlebitis and thrombophlebitis of femoral vein, bilateral |
| VTE | Venous Thromboembolism (DVT or PE) | ICD10 | I80.201 | Phlebitis and thrombophlebitis of unspecified deep vessels of right lower extremity |

| Phenotype | phenotype description | vocabulary | code | Code description |
|---|---|---|---|---|
| VTE | Venous Thromboembolism (DVT or PE) | ICD10 | I80.202 | Phlebitis and thrombophlebitis of unspecified deep vessels of left lower extremity |
| VTE | Venous Thromboembolism (DVT or PE) | ICD10 | I80.203 | Phlebitis and thrombophlebitis of unspecified deep vessels of lower extremities, bilateral |
| VTE | Venous Thromboembolism (DVT or PE) | ICD10 | I80.209 | Phlebitis and thrombophlebitis of unspecified deep vessels of unspecified lower extremity |
| VTE | Venous Thromboembolism (DVT or PE) | ICD10 | I80.211 | Phlebitis and thrombophlebitis of right iliac vein |
| VTE | Venous Thromboembolism (DVT or PE) | ICD10 | I80.212 | Phlebitis and thrombophlebitis of left iliac vein |
| VTE | Venous Thromboembolism (DVT or PE) | ICD10 | I80.213 | Phlebitis and thrombophlebitis of iliac vein, bilateral |
| VTE | Venous Thromboembolism (DVT or PE) | ICD10 | I80.219 | Phlebitis and thrombophlebitis of unspecified iliac vein |
| VTE | Venous Thromboembolism (DVT or PE) | ICD10 | I80.221 | Phlebitis and thrombophlebitis of right popliteal vein |
| VTE | Venous Thromboembolism (DVT or PE) | ICD10 | I80.222 | Phlebitis and thrombophlebitis of left popliteal vein |
| VTE | Venous Thromboembolism (DVT or PE) | ICD10 | I80.223 | Phlebitis and thrombophlebitis of popliteal vein, bilateral |

| Phenotype | phenotype description | vocabulary | code | Code description |
|---|---|---|---|---|
| VTE | Venous Thromboembolism (DVT or PE) | ICD10 | I80.229 | Phlebitis and thrombophlebitis of unspecified popliteal vein |
| VTE | Venous Thromboembolism (DVT or PE) | ICD10 | I80.231 | Phlebitis and thrombophlebitis of right tibial vein |
| VTE | Venous Thromboembolism (DVT or PE) | ICD10 | I80.232 | Phlebitis and thrombophlebitis of left tibial vein |
| VTE | Venous Thromboembolism (DVT or PE) | ICD10 | I80.233 | Phlebitis and thrombophlebitis of tibial vein, bilateral |
| VTE | Venous Thromboembolism (DVT or PE) | ICD10 | I80.239 | Phlebitis and thrombophlebitis of unspecified tibial vein |
| VTE | Venous Thromboembolism (DVT or PE) | ICD10 | I80.241 | Phlebitis and thrombophlebitis of right peroneal vein |
| VTE | Venous Thromboembolism (DVT or PE) | ICD10 | I80.242 | Phlebitis and thrombophlebitis of left peroneal vein |
| VTE | Venous Thromboembolism (DVT or PE) | ICD10 | I80.243 | Phlebitis and thrombophlebitis of peroneal vein, bilateral |
| VTE | Venous Thromboembolism (DVT or PE) | ICD10 | I80.249 | Phlebitis and thrombophlebitis of unspecified peroneal vein |
| VTE | Venous Thromboembolism (DVT or PE) | ICD10 | I80.251 | Phlebitis and thrombophlebitis of right calf muscular vein |

| Phenotype | phenotype description | vocabulary | code | Code description |
|---|---|---|---|---|
| VTE | Venous Thromboembolism (DVT or PE) | ICD10 | I80.252 | Phlebitis and thrombophlebitis of left calf muscular vein |
| VTE | Venous Thromboembolism (DVT or PE) | ICD10 | I80.253 | Phlebitis and thrombophlebitis of calf muscular vein, bilateral |
| VTE | Venous Thromboembolism (DVT or PE) | ICD10 | I80.259 | Phlebitis and thrombophlebitis of unspecified calf muscular vein |
| VTE | Venous Thromboembolism (DVT or PE) | ICD10 | I80.291 | Phlebitis and thrombophlebitis of other deep vessels of right lower extremity |
| VTE | Venous Thromboembolism (DVT or PE) | ICD10 | I80.292 | Phlebitis and thrombophlebitis of other deep vessels of left lower extremity |
| VTE | Venous Thromboembolism (DVT or PE) | ICD10 | I80.293 | Phlebitis and thrombophlebitis of other deep vessels of lower extremity, bilateral |
| VTE | Venous Thromboembolism (DVT or PE) | ICD10 | I80.299 | Phlebitis and thrombophlebitis of other deep vessels of unspecified lower extremity |
| VTE | Venous Thromboembolism (DVT or PE) | ICD10 | I82.210 | Acute embolism and thrombosis of superior vena cava |
| VTE | Venous Thromboembolism (DVT or PE) | ICD10 | I82.220 | Acute embolism and thrombosis of inferior vena cava |
| VTE | Venous Thromboembolism (DVT or PE) | ICD10 | I82.290 | Acute embolism and thrombosis of other thoracic veins |

| Phenotype | phenotype description | vocabulary | code | Code description |
|---|---|---|---|---|
| VTE | Venous Thromboembolism (DVT or PE) | ICD10 | I82.401 | Acute embolism and thrombosis of unspecified deep veins of right lower extremity |
| VTE | Venous Thromboembolism (DVT or PE) | ICD10 | I82.402 | Acute embolism and thrombosis of unspecified deep veins of left lower extremity |
| VTE | Venous Thromboembolism (DVT or PE) | ICD10 | I82.403 | Acute embolism and thrombosis of unspecified deep veins of lower extremity, bilateral |
| VTE | Venous Thromboembolism (DVT or PE) | ICD10 | I82.409 | Acute embolism and thrombosis of unspecified deep veins of unspecified lower extremity |
| VTE | Venous Thromboembolism (DVT or PE) | ICD10 | I82.411 | Acute embolism and thrombosis of right femoral vein |
| VTE | Venous Thromboembolism (DVT or PE) | ICD10 | I82.412 | Acute embolism and thrombosis of left femoral vein |
| VTE | Venous Thromboembolism (DVT or PE) | ICD10 | I82.413 | Acute embolism and thrombosis of femoral vein, bilateral |
| VTE | Venous Thromboembolism (DVT or PE) | ICD10 | I82.419 | Acute embolism and thrombosis of unspecified femoral vein |
| VTE | Venous Thromboembolism (DVT or PE) | ICD10 | I82.421 | Acute embolism and thrombosis of right iliac vein |
| VTE | Venous Thromboembolism (DVT or PE) | ICD10 | I82.422 | Acute embolism and thrombosis of left iliac vein |

| Phenotype | phenotype description | vocabulary | code | Code description |
|---|---|---|---|---|
| VTE | Venous Thromboembolism (DVT or PE) | ICD10 | I82.423 | Acute embolism and thrombosis of iliac vein, bilateral |
| VTE | Venous Thromboembolism (DVT or PE) | ICD10 | I82.429 | Acute embolism and thrombosis of unspecified iliac vein |
| VTE | Venous Thromboembolism (DVT or PE) | ICD10 | I82.431 | Acute embolism and thrombosis of right popliteal vein |
| VTE | Venous Thromboembolism (DVT or PE) | ICD10 | I82.432 | Acute embolism and thrombosis of left popliteal vein |
| VTE | Venous Thromboembolism (DVT or PE) | ICD10 | I82.433 | Acute embolism and thrombosis of popliteal vein, bilateral |
| VTE | Venous Thromboembolism (DVT or PE) | ICD10 | I82.439 | Acute embolism and thrombosis of unspecified popliteal vein |
| VTE | Venous Thromboembolism (DVT or PE) | ICD10 | I82.441 | Acute embolism and thrombosis of right tibial vein |
| VTE | Venous Thromboembolism (DVT or PE) | ICD10 | I82.442 | Acute embolism and thrombosis of left tibial vein |
| VTE | Venous Thromboembolism (DVT or PE) | ICD10 | I82.443 | Acute embolism and thrombosis of tibial vein, bilateral |
| VTE | Venous Thromboembolism (DVT or PE) | ICD10 | I82.449 | Acute embolism and thrombosis of unspecified tibial vein |

| Phenotype | phenotype description | vocabulary | code | Code description |
|---|---|---|---|---|
| VTE | Venous Thromboembolism (DVT or PE) | ICD10 | I82.451 | Acute embolism and thrombosis of right peroneal vein |
| VTE | Venous Thromboembolism (DVT or PE) | ICD10 | I82.452 | Acute embolism and thrombosis of left peroneal vein |
| VTE | Venous Thromboembolism (DVT or PE) | ICD10 | I82.453 | Acute embolism and thrombosis of peroneal vein, bilateral |
| VTE | Venous Thromboembolism (DVT or PE) | ICD10 | I82.459 | Acute embolism and thrombosis of unspecified peroneal vein |
| VTE | Venous Thromboembolism (DVT or PE) | ICD10 | I82.461 | Acute embolism and thrombosis of right calf muscular vein |
| VTE | Venous Thromboembolism (DVT or PE) | ICD10 | I82.462 | Acute embolism and thrombosis of left calf muscular vein |
| VTE | Venous Thromboembolism (DVT or PE) | ICD10 | I82.463 | Acute embolism and thrombosis of calf muscular vein, bilateral |
| VTE | Venous Thromboembolism (DVT or PE) | ICD10 | I82.469 | Acute embolism and thrombosis of unspecified calf muscular vein |
| VTE | Venous Thromboembolism (DVT or PE) | ICD10 | I82.491 | Acute embolism and thrombosis of other specified deep vein of right lower extremity |
| VTE | Venous Thromboembolism (DVT or PE) | ICD10 | I82.492 | Acute embolism and thrombosis of other specified deep vein of left lower extremity |

| Phenotype | phenotype description | vocabulary | code | Code description |
|---|---|---|---|---|
| VTE | Venous Thromboembolism (DVT or PE) | ICD10 | I82.493 | Acute embolism and thrombosis of other specified deep vein of lower extremity, bilateral |
| VTE | Venous Thromboembolism (DVT or PE) | ICD10 | I82.499 | Acute embolism and thrombosis of other specified deep vein of unspecified lower extremity |
| VTE | Venous Thromboembolism (DVT or PE) | ICD10 | I82.4Y1 | Acute embolism and thrombosis of unspecified deep veins of right proximal lower extremity |
| VTE | Venous Thromboembolism (DVT or PE) | ICD10 | I82.4Y2 | Acute embolism and thrombosis of unspecified deep veins of left proximal lower extremity |
| VTE | Venous Thromboembolism (DVT or PE) | ICD10 | I82.4Y3 | Acute embolism and thrombosis of unspecified deep veins of proximal lower extremity, bilateral |
| VTE | Venous Thromboembolism (DVT or PE) | ICD10 | I82.4Y9 | Acute embolism and thrombosis of unspecified deep veins of unspecified proximal lower extremity |
| VTE | Venous Thromboembolism (DVT or PE) | ICD10 | I82.4Z1 | Acute embolism and thrombosis of unspecified deep veins of right distal lower extremity |
| VTE | Venous Thromboembolism (DVT or PE) | ICD10 | I82.4Z2 | Acute embolism and thrombosis of unspecified deep veins of left distal lower extremity |
| VTE | Venous Thromboembolism (DVT or PE) | ICD10 | I82.4Z3 | Acute embolism and thrombosis of unspecified deep veins of distal lower extremity, bilateral |
| VTE | Venous Thromboembolism (DVT or PE) | ICD10 | I82.4Z9 | Acute embolism and thrombosis of unspecified deep veins of unspecified distal lower extremity |

| Phenotype | phenotype description | vocabulary | code | Code description |
|---|---|---|---|---|
| VTE | Venous Thromboembolism (DVT or PE) | ICD10 | I82.621 | Acute embolism and thrombosis of deep veins of right upper extremity |
| VTE | Venous Thromboembolism (DVT or PE) | ICD10 | I82.622 | Acute embolism and thrombosis of deep veins of left upper extremity |
| VTE | Venous Thromboembolism (DVT or PE) | ICD10 | I82.623 | Acute embolism and thrombosis of deep veins of upper extremity, bilateral |
| VTE | Venous Thromboembolism (DVT or PE) | ICD10 | I82.629 | Acute embolism and thrombosis of deep veins of unspecified upper extremity |
| VTE | Venous Thromboembolism (DVT or PE) | ICD10 | I82.A11 | Acute embolism and thrombosis of right axillary vein |
| VTE | Venous Thromboembolism (DVT or PE) | ICD10 | I82.A12 | Acute embolism and thrombosis of left axillary vein |
| VTE | Venous Thromboembolism (DVT or PE) | ICD10 | I82.A13 | Acute embolism and thrombosis of axillary vein, bilateral |
| VTE | Venous Thromboembolism (DVT or PE) | ICD10 | I82.A19 | Acute embolism and thrombosis of unspecified axillary vein |
| VTE | Venous Thromboembolism (DVT or PE) | ICD10 | I82.B11 | Acute embolism and thrombosis of right subclavian vein |
| VTE | Venous Thromboembolism (DVT or PE) | ICD10 | I82.B12 | Acute embolism and thrombosis of left subclavian vein |

| Phenotype | phenotype description | vocabulary | code | Code description |
|---|---|---|---|---|
| VTE | Venous Thromboembolism (DVT or PE) | ICD10 | I82.B13 | Acute embolism and thrombosis of subclavian vein, bilateral |
| VTE | Venous Thromboembolism (DVT or PE) | ICD10 | I82.B19 | Acute embolism and thrombosis of unspecified subclavian vein |
| VTE | Venous Thromboembolism (DVT or PE) | ICD10 | I82.C11 | Acute embolism and thrombosis of right internal jugular vein |
| VTE | Venous Thromboembolism (DVT or PE) | ICD10 | I82.C12 | Acute embolism and thrombosis of left internal jugular vein |
| VTE | Venous Thromboembolism (DVT or PE) | ICD10 | I82.C13 | Acute embolism and thrombosis of internal jugular vein, bilateral |
| VTE | Venous Thromboembolism (DVT or PE) | ICD10 | I82.C19 | Acute embolism and thrombosis of unspecified internal jugular vein |
| VTE | Venous Thromboembolism (DVT or PE) | ICD9 | 415.11 | IATROGENIC PULMONARY EMBOLISM AND INFARCTION |
| VTE | Venous Thromboembolism (DVT or PE) | ICD9 | 415.12 | SEPTIC PULMONARY EMBOLISM |
| VTE | Venous Thromboembolism (DVT or PE) | ICD9 | 415.13 | SADDLE EMBOLUS OF PULMONARY ARTERY |
| VTE | Venous Thromboembolism (DVT or PE) | ICD9 | 415.19 | OTHER PULMONARY EMBOLISM AND INFARCTION |

| Phenotype | phenotype description | vocabulary | code | Code description |
|---|---|---|---|---|
| VTE | Venous Thromboembolism (DVT or PE) | ICD9 | 451.11 | PHLEBITIS AND THROMBOPHLEBITIS OF FEMORAL VEIN (DEEP)(SUPERFICIAL) |
| VTE | Venous Thromboembolism (DVT or PE) | ICD9 | 451.19 | PHLEBITIS AND THROMBOPHLEBITIS OF OTHER DEEP VESSELS OF LOWER EXTREMITIES |
| VTE | Venous Thromboembolism (DVT or PE) | ICD9 | 451.81 | PHLEBITIS AND THROMBOPHLEBITIS OF ILIAC VEIN |
| VTE | Venous Thromboembolism (DVT or PE) | ICD9 | 451.83 | PHLEBITIS & THROMBOPHLEBITIS OF DEEP VEINS OF UPPER EXTREMITIES |
| VTE | Venous Thromboembolism (DVT or PE) | ICD9 | 453.2 | OTHER VENOUS EMBOLISM AND THROMBOSIS OF INFERIOR VENA CAVA |
| VTE | Venous Thromboembolism (DVT or PE) | ICD9 | 453.4 | ACUTE VENOUS EMBOLISM AND THROMBOSIS OF UNSPECIFIED DEEP VESSELS OF LOWER EXTREMITY |
| VTE | Venous Thromboembolism (DVT or PE) | ICD9 | 453.41 | ACUTE VENOUS EMBOLISM AND THROMBOSIS OF DEEP VESSELS OF PROXIMAL LOWER EXTREMITY |
| VTE | Venous Thromboembolism (DVT or PE) | ICD9 | 453.42 | ACUTE VENOUS EMBOLISM AND THROMBOSIS OF DEEP VESSELS OF DISTAL LOWER EXTREMITY |
| VTE | Venous Thromboembolism (DVT or PE) | ICD9 | 453.82 | ACUTE VENOUS EMBOLISM AND THROMBOSIS OF DEEP VEINS OF UPPER EXTREMITY |
| VTE | Venous Thromboembolism (DVT or PE) | ICD9 | 453.83 | ACUTE VENOUS EMBOLISM AND THROMBOSIS OF UPPER EXTREMITY, UNSPECIFIED |

| Phenotype | phenotype description | vocabulary | code | Code description |
|---|---|---|---|---|
| VTE | Venous Thromboembolism (DVT or PE) | ICD9 | 453.84 | ACUTE VENOUS EMBOLISM AND THROMBOSIS OF AXILLARY VEINS |
| VTE | Venous Thromboembolism (DVT or PE) | ICD9 | 453.85 | ACUTE VENOUS EMBOLISM AND THROMBOSIS OF SUBCLAVIAN VEINS |
| VTE | Venous Thromboembolism (DVT or PE) | ICD9 | 453.86 | ACUTE VENOUS EMBOLISM AND THROMBOSIS OF INTERNAL JUGULAR VEINS |
| VTE | Venous Thromboembolism (DVT or PE) | ICD9 | 453.87 | ACUTE VENOUS EMBOLISM AND THROMBOSIS OF OTHER THORACIC VEINS |

**Supplemental Table 2. List of outpatient anticoagulation medications included in the anticoagulation medication variable.**

| |
|---|
| warfarin |
| enoxaparin |
| fondaparinux |
| dalteparin |
| dabigatran |
| bivalirudin |
| betrixaban |
| rivaroxaban |
| apixaban |
| pegnivacogin |
| sulodexide |
| argatroban |
| ximelagatran |
| miradon |
| anisindione |
| danaparoid |
| lepirudin |
| tinzaparin |
| betrixaban |
| clexane |
| fraxiparine |
| endoxaban |
| desirudin |
| jantoven |
| idraparinux |
| melagatran |
| otamixaban |

**Supplemental Table 3. 297 SNP rsIDs and PRS coefficients (Effect column) used to implement the polygenic risk score for VTE, as well as rs6025 (*F5*) and rs1799963 (*F2*) which were analyzed separately. 287 SNPs utilized in this analysis are denoted with an X in column 287_snps_used.**

| MarkerName | cptid | Allele1 | Allele2 | Effect | Consequence | SYMBOL | 287_snps_used |
|---|---|---|---|---|---|---|---|
| rs6025 | 1:169519049 | t | c | 0.926 | missense_variant | F5 | |
| rs9411377 | 9:136145404 | a | c | 0.308 | intron_variant | ABO | X |
| rs78516619 | 1:168889373 | g | a | 0.6841 | intron_variant | LINC00970 | X |
| rs56810541 | 4:187200550 | t | a | 0.1975 | intron_variant | F11 | X |
| rs13130318 | 4:155538470 | g | t | 0.1984 | upstream_gene_variant | FGG | X |
| rs41302673 | 9:136270538 | g | t | 0.242 | missense_variant | C9orf96 | X |
| rs2143289 | 1:169086013 | t | c | 0.2335 | intron_variant | ATP1B1 | X |
| rs1799963 | 11:46761055 | a | g | 0.6337 | 3_prime_UTR_variant | F2 | |
| rs11244035 | 9:136081319 | t | c | 0.2223 | missense_variant | OBP2B | X |
| rs78707713 | 10:71245276 | t | c | 0.2414 | intron_variant | TSPAN15 | X |
| rs3775303 | 4:187178014 | t | a | 0.1486 | intron_variant | KLKB1 | X |
| rs4253425 | 4:187205929 | c | t | 0.2399 | intron_variant | F11 | X |
| rs4656683 | 1:169463519 | t | c | 0.1781 | intergenic_variant | NA | X |
| rs185120584 | 1:169659128 | t | c | 0.5696 | downstream_gene_variant | SELL | X |
| rs9411395 | 9:136184782 | g | a | 0.1408 | intron_variant | LCN1P2 | X |
| rs116140155 | 1:169014610 | a | g | 0.2498 | intron_variant | LINC00970 | X |

| MarkerName | cptid | Allele1 | Allele2 | Effect | Consequence | SYMBOL | 287_snps_used |
|---|---|---|---|---|---|---|---|
| rs557317 | 9:136157037 | a | c | 0.1499 | intergenic_variant | NA | X |
| rs11789139 | 9:136185324 | g | c | 0.1669 | downstream_gene_variant | LCN1P2 | X |
| rs6032 | 1:169511555 | t | c | 0.1351 | missense_variant | F5 | X |
| rs11244051 | 9:136128731 | a | g | 0.384 | non_coding_transcript_exon_variant | ABO | X |
| rs8176630 | 9:136152722 | c | t | 0.1382 | upstream_gene_variant | ABO | X |
| rs3094373 | 9:136323826 | a | t | 0.2432 | intron_variant | ADAMTS13 | X |
| rs13435192 | 4:155450158 | t | c | 0.1124 | downstream_gene_variant | RP11-158C21.2 | X |
| rs6060288 | 20:33772243 | a | g | 0.1178 | intron_variant | EDEM2 | X |
| rs3917862 | 1:169593113 | g | a | 0.2198 | intron_variant | SELP | X |
| rs2856656 | 11:47373425 | c | t | 0.3976 | intron_variant | MYBPC3 | X |
| rs78755596 | 9:136124590 | a | t | 0.2855 | downstream_gene_variant | ABO | X |
| rs149181677 | 9:136296530 | t | c | 0.2461 | intron_variant | ADAMTS13 | X |
| rs113123846 | 1:168937072 | g | a | 0.4417 | intron_variant | LINC00970 | X |
| rs13126546 | 4:187220894 | t | c | 0.1142 | intron_variant | F11-AS1 | X |
| rs11791119 | 9:136184985 | t | c | 0.1052 | intron_variant | LCN1P2 | X |
| rs12117978 | 1:168729611 | a | g | 0.2166 | downstream_gene_variant | RP1-10C16.1 | X |

| MarkerName | cptid | Allele1 | Allele2 | Effect | Consequence | SYMBOL | 287_snps_used |
|---|---|---|---|---|---|---|---|
| rs12710257 | 19:10741622 | g | a | 0.1164 | intron_variant | SLC44A2 | X |
| rs12445050 | 16:81870969 | t | c | 0.1248 | intron_variant | PLCG2 | X |
| rs1200118 | 1:169064630 | g | a | 0.1111 | regulatory_region_variant | NA | X |
| rs652600 | 9:136311017 | a | g | 0.0992 | intron_variant | ADAMTS13 | X |
| rs4323084 | 4:155424231 | t | c | 0.1012 | intergenic_variant | NA | X |
| rs7520186 | 1:169107377 | t | c | 0.0873 | intron_variant | NME7 | X |
| rs8176634 | 9:136152070 | g | c | 0.1117 | upstream_gene_variant | ABO | X |
| rs11244032 | 9:136077004 | c | g | 0.0837 | downstream_gene_variant | OBP2B | X |
| rs79726896 | 4:155494926 | g | a | 0.1598 | downstream_gene_variant | FGB | X |
| rs10886430 | 10:121010256 | g | a | 0.1289 | intron_variant | GRK5 | X |
| rs10732287 | 1:169276816 | t | c | 0.0916 | intron_variant | NME7 | X |
| rs149189328 | 9:136080512 | c | a | 0.2669 | downstream_gene_variant | OBP2B | X |
| rs1933116 | 1:168619548 | t | c | 0.1724 | intergenic_variant | NA | X |
| rs183356 | 12:6150824 | a | g | 0.1405 | intron_variant | VWF | X |
| rs113079063 | 1:169031755 | t | g | 0.2356 | intron_variant | LINC00970 | X |
| rs174548 | 11:61571348 | c | g | 0.0842 | intron_variant | FADS1 | X |

| MarkerName | cptid | Allele1 | Allele2 | Effect | Consequence | SYMBOL | 287_snps_used |
|---|---|---|---|---|---|---|---|
| rs1665581 | 10:71262048 | g | t | 0.0771 | intron_variant | TSPAN15 | X |
| rs4541868 | 8:106590705 | c | a | 0.0876 | intron_variant | ZFPM2 | X |
| rs7135039 | 12:6160614 | t | c | 0.0781 | intron_variant | VWF | X |
| rs72705895 | 1:168572720 | t | c | 0.1313 | regulatory_region_variant | NA | X |
| rs7855466 | 9:136121303 | t | c | 0.1054 | downstream_gene_variant | ABO | X |
| rs2285488 | 9:136277854 | g | a | 0.1007 | intron_variant | REXO4 | X |
| rs3094326 | 9:136343647 | g | c | 0.1027 | intron_variant | SLC2A6 | X |
| rs72681247 | 4:155544958 | t | c | 0.2163 | upstream_gene_variant | LRAT | X |
| rs72703796 | 1:168500034 | g | a | 0.2014 | intergenic_variant | NA | X |
| rs59234924 | 4:155542926 | g | a | 0.1077 | intergenic_variant | NA | X |
| rs3124747 | 9:136268084 | a | g | 0.0757 | missense_variant | C9orf96 | X |
| rs28615587 | 9:136365146 | t | g | 0.078 | intergenic_variant | NA | X |
| rs1799809 | 2:128175875 | g | a | 0.072 | upstream_gene_variant | PROC | X |
| rs35990973 | 1:168969254 | a | t | 0.1057 | intron_variant | LINC00970 | X |
| rs113435394 | 4:187136519 | c | t | 0.2091 | downstream_gene_variant | CYP4V2 | X |
| rs117119759 | 9:136212168 | a | g | 0.2099 | intron_variant | MED22 | X |

| MarkerName | cptid | Allele1 | Allele2 | Effect | Consequence | SYMBOL | 287_snps_used |
|---|---|---|---|---|---|---|---|
| rs12416320 | 10:71148728 | g | a | 0.1553 | intron_variant | HK1 | X |
| rs78590974 | 9:136156230 | t | c | 0.2221 | regulatory_region_variant | NA | X |
| rs564783003 | 20:33612647 | c | a | 0.1488 | intron_variant | TRPC4AP | X |
| rs10998791 | 10:71218059 | a | c | 0.0677 | intron_variant | TSPAN15 | X |
| rs10441806 | 9:136062437 | c | t | 0.105 | intergenic_variant | NA | X |
| rs55988407 | 9:136156064 | g | a | 0.1664 | regulatory_region_variant | NA | X |
| rs11038913 | 11:46559730 | t | c | 0.1126 | intron_variant | AMBRA1 | X |
| rs2306029 | 11:46893108 | t | c | 0.0641 | missense_variant | LRP4 | X |
| rs4547780 | 4:155432944 | g | a | 0.064 | intergenic_variant | NA | X |
| rs2070008 | 4:155513276 | t | c | 0.0975 | upstream_gene_variant | FGA | X |
| rs72712626 | 4:187234735 | g | a | 0.1365 | intron_variant | F11-AS1 | X |
| rs543926510 | 11:56526894 | c | t | 0.1339 | intergenic_variant | NA | X |
| rs77542162 | 17:67081278 | g | a | 0.1932 | missense_variant | ABCA6 | X |
| rs3124755 | 9:136240304 | c | t | 0.1278 | intron_variant | SURF4 | X |
| rs137936874 | 10:71215107 | g | t | 0.2231 | intron_variant | TSPAN15 | X |
| rs1048483 | 17:1966457 | t | c | 0.0616 | intron_variant | SMG6 | X |

| MarkerName | cptid | Allele1 | Allele2 | Effect | Consequence | SYMBOL | 287_snps_used |
|---|---|---|---|---|---|---|---|
| rs925452 | 4:187190285 | a | t | 0.2681 | intron_variant | F11 | |
| rs56379917 | 4:187219599 | a | g | 0.1638 | intron_variant | F11-AS1 | X |
| rs4962043 | 9:136177993 | g | a | 0.0607 | non_coding_transcript_exon_variant | Y_RNA | X |
| rs28413626 | 12:123861452 | g | a | 0.0739 | intergenic_variant | NA | X |
| rs9802874 | 9:136382716 | a | g | 0.0636 | intron_variant | TMEM8C | X |
| rs3211752 | 13:113787459 | g | a | 0.06 | intron_variant | F10 | X |
| rs1613662 | 19:55536595 | a | g | 0.0818 | missense_variant | GP6 | X |
| rs6083037 | 20:23182559 | a | t | 0.0798 | intergenic_variant | NA | X |
| rs9373523 | 6:147701133 | g | t | 0.06 | intron_variant | STXBP5 | X |
| rs76771223 | 9:136193356 | g | a | 0.1431 | downstream_gene_variant | SURF6 | X |
| rs13137269 | 4:187240280 | t | a | 0.0617 | intron_variant | F11-AS1 | X |
| rs7027827 | 9:136031918 | a | g | 0.0729 | intron_variant | GBGT1 | X |
| rs75440104 | 4:187267791 | c | t | 0.1177 | intron_variant | F11-AS1 | X |
| rs1867312 | 2:68619981 | c | a | 0.0585 | intron_variant | PLEK | X |
| rs551251712 | 4:187161283 | c | t | 0.2385 | intron_variant | KLKB1 | X |
| rs12747018 | 1:169038856 | t | c | 0.0717 | intron_variant | LINC00970 | X |

| MarkerName | cptid | Allele1 | Allele2 | Effect | Consequence | SYMBOL | 287_snps_used |
|---|---|---|---|---|---|---|---|
| rs13300181 | 9:136390015 | a | g | 0.0616 | 5_prime_UTR_variant | TMEM8C | X |
| rs34397775 | 20:23170450 | a | g | 0.084 | non_coding_transcript_exon_variant | RP4-737E23.2 | X |
| rs2644120 | 1:201882277 | c | g | 0.058 | intron_variant | LMOD1 | X |
| rs2203111 | 4:187169469 | g | a | 0.0901 | intron_variant | KLKB1 | X |
| rs117653193 | 11:50242788 | t | c | 0.1049 | downstream_gene_variant | RP11-347H15.5 | X |
| rs12567872 | 1:168577239 | t | g | 0.0893 | intergenic_variant | NA | X |
| rs72910502 | 11:55436134 | c | g | 0.1161 | downstream_gene_variant | OR4C6 | X |
| rs115999709 | 4:155474683 | g | a | 0.1456 | upstream_gene_variant | PLRG1 | X |
| rs56244533 | 20:33434252 | a | t | 0.0567 | intron_variant | GGT7 | X |
| rs62575992 | 9:136255149 | c | t | 0.0967 | intron_variant | C9orf96 | X |
| rs966751 | 1:169486141 | g | c | 0.1142 | intron_variant | F5 | X |
| rs2685413 | 8:27810577 | g | t | 0.0608 | intron_variant | SCARA5 | X |
| rs190543502 | 15:43757184 | t | c | 0.1973 | intron_variant | TP53BP1 | X |
| rs12144655 | 1:170174298 | a | g | 0.0893 | intron_variant | RP11-297H3.3 | X |
| rs34258243 | 1:168812398 | g | c | 0.1015 | intergenic_variant | NA | X |
| rs61988257 | 14:92217670 | a | g | 0.0603 | intron_variant | CATSPERB | X |

| MarkerName | cptid | Allele1 | Allele2 | Effect | Consequence | SYMBOL | 287_snps_used |
|---|---|---|---|---|---|---|---|
| rs2069946 | 20:33762035 | c | t | 0.1255 | intron_variant | PROCR | X |
| rs7039497 | 9:136069931 | g | a | 0.1081 | intergenic_variant | NA | X |
| rs75627267 | 20:33451060 | t | c | 0.1209 | intron_variant | GGT7 | X |
| rs3793846 | 10:71153882 | t | c | 0.0572 | intron_variant | HK1 | X |
| rs12128208 | 1:170080014 | c | t | 0.1373 | upstream_gene_variant | SIGLEC30P | X |
| rs141912156 | 3:89679389 | t | c | 0.1656 | intergenic_variant | NA | X |
| rs16867574 | 5:38708554 | c | t | 0.059 | downstream_gene_variant | RP11-122C5.1 | X |
| rs11507716 | 9:136397195 | t | c | 0.0802 | upstream_gene_variant | ADAMTSL2 | X |
| rs79201245 | 4:155553541 | c | t | 0.1462 | intron_variant | LRAT | X |
| rs9620086 | 22:43107837 | g | c | 0.0609 | intron_variant | A4GALT | X |
| rs7526462 | 1:168909952 | t | c | 0.0714 | intron_variant | LINC00970 | X |
| rs141798115 | 11:56875074 | t | c | 0.2207 | intergenic_variant | NA | X |
| rs4253414 | 4:187196853 | c | t | 0.1544 | intron_variant | F11 | X |
| rs147474835 | 1:169463296 | g | t | 0.12 | intergenic_variant | NA | X |
| rs2842700 | 1:207282149 | a | c | 0.1138 | intron_variant | C4BPA | X |
| rs563259534 | 11:32967270 | t | c | 0.1048 | intron_variant | QSER1 | |

| MarkerName | cptid | Allele1 | Allele2 | Effect | Consequence | SYMBOL | 287_snps_used |
|---|---|---|---|---|---|---|---|
| rs11606922 | 11:51282525 | c | a | 0.1115 | intergenic_variant | NA | |
| rs1892091 | 1:169070213 | c | t | 0.0526 | upstream_gene_variant | ATP1B1 | X |
| rs3888561 | 9:136025460 | c | g | 0.0559 | downstream_gene_variant | GBGT1 | X |
| rs4805881 | 19:33896432 | c | a | 0.0544 | intron_variant | PEPD | X |
| rs8107372 | 19:10898413 | t | c | 0.0515 | intron_variant | DNM2 | X |
| rs61374069 | 16:81874200 | a | g | 0.0524 | intron_variant | PLCG2 | X |
| rs7622284 | 3:39240559 | t | c | 0.0741 | intergenic_variant | NA | X |
| rs139727584 | 12:6071943 | c | t | 0.1076 | intron_variant | VWF | X |
| rs12274057 | 11:73283937 | t | a | 0.0751 | intron_variant | FAM168A | X |
| rs3095304 | 6:31092767 | t | c | 0.0634 | intron_variant | PSORS1C1 | X |
| rs12785008 | 10:71333897 | c | t | 0.0691 | upstream_gene_variant | NEUROG3 | X |
| rs1984906 | 4:155515486 | g | a | 0.0671 | upstream_gene_variant | FGA | X |
| rs577695638 | 1:207285043 | c | a | 0.1708 | intron_variant | C4BPA | X |
| rs11668544 | 19:10659971 | g | c | 0.1314 | intron_variant | ATG4D | X |
| rs28470788 | 9:136098498 | t | g | 0.0723 | downstream_gene_variant | LCN1P1 | X |
| rs72646294 | 4:187130620 | a | g | 0.1947 | intron_variant | CYP4V2 | X |

| MarkerName | cptid | Allele1 | Allele2 | Effect | Consequence | SYMBOL | 287_snps_used |
|---|---|---|---|---|---|---|---|
| rs67076363 | 2:128398892 | a | g | 0.0733 | intron_variant | LIMS2 | X |
| rs698915 | 1:150388318 | a | g | 0.0561 | intron_variant | RPRD2 | X |
| rs2305196 | 10:71144324 | a | g | 0.0555 | intron_variant | HK1 | X |
| rs7374904 | 3:89250573 | a | g | 0.0751 | intron_variant | EPHA3 | X |
| rs2074492 | 6:31239869 | t | c | 0.051 | upstream_gene_variant | HLA-C | X |
| rs115476742 | 1:170051841 | c | t | 0.0857 | intron_variant | KIFAP3 | X |
| rs631126 | 18:8800723 | c | t | 0.0579 | intron_variant | SOGA2 | X |
| rs7004172 | 8:108340982 | g | c | 0.051 | intron_variant | ANGPT1 | X |
| rs10793953 | 9:136056956 | g | a | 0.0496 | intergenic_variant | NA | X |
| rs2050652 | 20:33733180 | g | a | 0.0564 | intron_variant | EDEM2 | X |
| rs66697526 | 9:136144593 | g | t | 0.1399 | intron_variant | ABO | X |
| rs140438685 | 10:76189250 | a | g | 0.2024 | intron_variant | ADK | X |
| rs1627764 | 6:29894392 | g | t | 0.0525 | non_coding_transcript_exon_variant | HCG4B | X |
| rs116333064 | 4:155251071 | a | g | 0.1157 | intron_variant | DCHS2 | X |
| rs78915411 | 12:6170645 | g | a | 0.0823 | intron_variant | VWF | X |
| rs3761824 | 9:135985796 | c | t | 0.0538 | synonymous_variant | RALGDS | X |

| MarkerName | cptid | Allele1 | Allele2 | Effect | Consequence | SYMBOL | 287_snps_used |
|---|---|---|---|---|---|---|---|
| rs369876615 | 11:48865680 | c | a | 0.1184 | intergenic_variant | NA | X |
| rs2341097 | 19:46268902 | t | c | 0.0502 | missense_variant | SIX5 | X |
| rs10918970 | 1:168714137 | t | c | 0.0572 | downstream_gene_variant | AL049798.1 | X |
| rs9290378 | 3:93580976 | t | g | 0.0509 | intergenic_variant | NA | X |
| rs3811444 | 1:248039451 | c | t | 0.0513 | missense_variant | TRIM58 | X |
| rs12075684 | 1:169620673 | g | a | 0.0534 | intergenic_variant | NA | X |
| rs137870902 | 12:39156743 | t | c | 0.2329 | intron_variant | CPNE8 | X |
| rs10087301 | 8:27820792 | a | g | 0.0632 | intron_variant | SCARA5 | X |
| rs10158131 | 1:169316610 | g | a | 0.0673 | intron_variant | NME7 | X |
| rs8100818 | 19:10639312 | t | c | 0.0483 | intergenic_variant | NA | X |
| rs111736896 | 2:127979335 | a | c | 0.0717 | upstream_gene_variant | CYP27C1 | X |
| rs1071644 | 16:81971403 | c | t | 0.0475 | synonymous_variant | PLCG2 | X |
| rs141397052 | 9:136226421 | g | c | 0.2259 | intron_variant | SURF2 | X |
| rs6137727 | 20:22672552 | a | g | 0.0534 | regulatory_region_variant | NA | X |
| rs113114306 | 3:36140951 | t | a | 0.1047 | intergenic_variant | NA | X |
| rs13013670 | 2:68530180 | t | c | 0.0485 | intron_variant | CNRIP1 | X |

| MarkerName | cptid | Allele1 | Allele2 | Effect | Consequence | SYMBOL | 287_snps_used |
|---|---|---|---|---|---|---|---|
| rs3751198 | 12:104147207 | g | a | 0.0491 | intron_variant | STAB2 | X |
| rs12858483 | 13:113808274 | g | c | 0.0588 | upstream_gene_variant | PROZ | X |
| rs7812868 | 8:87143573 | c | t | 0.0621 | intron_variant | ATP6V0D2 | X |
| rs2854827 | 22:42461918 | g | a | 0.0589 | intron_variant | NAGA | X |
| rs5896 | 11:46745003 | t | c | 0.0668 | missense_variant | F2 | X |
| rs143383 | 20:34025983 | g | a | 0.0482 | intron_variant | GDF5 | X |
| rs149439892 | 20:23077117 | a | g | 0.1527 | intergenic_variant | NA | |
| rs6058218 | 20:33858772 | g | a | 0.0552 | intron_variant | MMP24 | X |
| rs56347914 | 5:75992254 | c | a | 0.0843 | intron_variant | IQGAP2 | X |
| rs78209469 | 17:7785590 | t | c | 0.0867 | upstream_gene_variant | CHD3 | X |
| rs139156297 | 3:93518545 | t | c | 0.1588 | intergenic_variant | NA | X |
| rs11150422 | 16:81915832 | g | c | 0.053 | intron_variant | PLCG2 | X |
| rs11600151 | 11:126300537 | t | c | 0.0793 | intron_variant | KIRREL3 | X |
| rs736418 | 9:136359182 | g | a | 0.0556 | regulatory_region_variant | NA | X |
| rs1547643 | 10:96011865 | g | t | 0.0476 | intron_variant | PLCE1 | X |
| rs738408 | 22:44324730 | c | t | 0.0566 | synonymous_variant | PNPLA3 | X |

| MarkerName | cptid | Allele1 | Allele2 | Effect | Consequence | SYMBOL | 287_snps_used |
|---|---|---|---|---|---|---|---|
| rs185663249 | 20:34562935 | a | t | 0.1487 | intron_variant | CNBD2 | X |
| rs12981072 | 19:49241006 | c | g | 0.0469 | intron_variant | RASIP1 | X |
| rs111414961 | 1:168406710 | a | g | 0.1183 | intergenic_variant | NA | |
| rs764024 | 1:166533517 | t | c | 0.0502 | upstream_gene_variant | FMO8P | X |
| rs72712610 | 4:187213360 | a | g | 0.1509 | downstream_gene_variant | F11 | X |
| rs12783163 | 10:71346272 | a | t | 0.0596 | downstream_gene_variant | RP11-343J3.5 | X |
| rs73487492 | 11:61621611 | a | g | 0.1034 | intron_variant | FADS2 | X |
| rs1611128 | 9:136509514 | g | a | 0.0495 | intron_variant | DBH | X |
| rs611769 | 1:168673803 | c | a | 0.0562 | intron_variant | DPT | X |
| rs36054387 | 10:71181371 | c | g | 0.0729 | upstream_gene_variant | TACR2 | X |
| rs16956040 | 16:81976177 | c | t | 0.0613 | intron_variant | PLCG2 | X |
| rs141622900 | 19:45426792 | a | g | 0.1007 | downstream_gene_variant | APOC1 | X |
| rs6076004 | 20:23000653 | t | a | 0.0472 | intron_variant | RP4-753D10.5 | X |
| rs211416 | 10:32397591 | t | a | 0.0684 | upstream_gene_variant | RP11-241I20.4 | X |
| rs115384559 | 15:43911751 | t | g | 0.3853 | upstream_gene_variant | STRC | X |
| rs632793 | 1:11910677 | g | a | 0.0466 | upstream_gene_variant | NPPA | X |

| MarkerName | cptid | Allele1 | Allele2 | Effect | Consequence | SYMBOL | 287_snps_used |
|---|---|---|---|---|---|---|---|
| rs146383320 | 9:124362398 | t | g | 0.1278 | intron_variant | DAB2IP | X |
| rs57222984 | 17:43758898 | g | a | 0.0522 | intron_variant | RP11-105N13.4 | X |
| rs677665 | 1:9341786 | t | c | 0.0521 | downstream_gene_variant | Z98044.1 | X |
| rs4889419 | 16:81902990 | g | a | 0.0575 | intron_variant | PLCG2 | X |
| rs17092148 | 20:33435161 | g | t | 0.0612 | intron_variant | GGT7 | X |
| rs11712865 | 3:6084766 | g | c | 0.0799 | intron_variant | AC027119.1 | X |
| rs2229678 | 14:66082793 | c | a | 0.1842 | missense_variant | FUT8 | X |
| rs1347390 | 7:157763424 | g | a | 0.0515 | intron_variant | PTPRN2 | X |
| rs6059574 | 20:32523172 | g | c | 0.0479 | intergenic_variant | NA | X |
| rs148770227 | 15:44880783 | c | t | 0.1837 | intron_variant | SPG11 | |
| rs198428 | 11:61489705 | a | t | 0.0464 | intron_variant | DAGLA | X |
| rs9858006 | 3:90390344 | a | t | 0.0465 | intergenic_variant | NA | X |
| rs7685922 | 4:187347217 | c | t | 0.054 | downstream_gene_variant | RP11-215A19.2 | X |
| rs3936939 | 1:181031783 | a | g | 0.0458 | downstream_gene_variant | MR1 | X |
| rs1322487 | 1:168960286 | a | g | 0.046 | intron_variant | LINC00970 | X |
| rs67495946 | 2:128187428 | c | t | 0.0631 | downstream_gene_variant | PROC | X |

| MarkerName | cptid | Allele1 | Allele2 | Effect | Consequence | SYMBOL | 287_snps_used |
|---|---|---|---|---|---|---|---|
| rs17502085 | 15:96125226 | a | g | 0.0525 | intergenic_variant | NA | X |
| rs35204896 | 15:65114833 | g | a | 0.0738 | intron_variant | PIF1 | X |
| rs551986443 | 20:33587569 | c | t | 0.1488 | intron_variant | MYH7B | |
| rs3088075 | 1:230417394 | t | c | 0.0798 | 3_prime_UTR_variant | GALNT2 | X |
| rs115110838 | 3:185234111 | t | c | 0.0649 | intron_variant | LIPH | X |
| rs2211163 | 10:45632668 | a | g | 0.0724 | intron_variant | CUBNP3 | X |
| rs1841009 | 3:88354842 | t | g | 0.0567 | intergenic_variant | NA | X |
| rs34603417 | 16:81840709 | a | g | 0.0456 | intron_variant | PLCG2 | X |
| rs6058227 | 20:33895947 | t | c | 0.0813 | intron_variant | UQCC1 | X |
| rs61926202 | 12:32844798 | a | g | 0.2109 | intron_variant | DNM1L | X |
| rs6087538 | 20:32485961 | c | t | 0.0585 | intergenic_variant | NA | X |
| rs116468525 | 16:1405044 | a | g | 0.2059 | intron_variant | GNPTG | X |
| rs4236786 | 8:108291878 | c | g | 0.0512 | intron_variant | ANGPT1 | X |
| rs75347181 | 3:150872174 | a | g | 0.1323 | intron_variant | MED12L | X |
| rs62112094 | 18:74465347 | a | g | 0.059 | downstream_gene_variant | AC139085.1 | X |
| rs10198483 | 2:127934102 | a | g | 0.0559 | regulatory_region_variant | NA | X |

| MarkerName | cptid | Allele1 | Allele2 | Effect | Consequence | SYMBOL | 287_snps_used |
|---|---|---|---|---|---|---|---|
| rs116764841 | 3:194787481 | t | c | 0.0661 | downstream_gene_variant | XXYLT1 | X |
| rs1358714 | 1:169079419 | a | g | 0.0444 | intron_variant | ATP1B1 | X |
| rs874492 | 19:7832001 | a | t | 0.0481 | intron_variant | CLEC4M | X |
| rs2061997 | 11:33247621 | t | c | 0.049 | intergenic_variant | NA | X |
| rs4253333 | 4:187180115 | g | a | 0.0494 | downstream_gene_variant | KLKB1 | X |
| rs214057 | 6:25531133 | c | t | 0.0446 | intron_variant | LRRC16A | X |
| rs142170418 | 19:3464793 | c | t | 0.1574 | downstream_gene_variant | NFIC | X |
| rs11721316 | 3:126238371 | a | t | 0.061 | upstream_gene_variant | UROC1 | X |
| rs1769758 | 10:80898969 | g | t | 0.0436 | intron_variant | ZMIZ1 | X |
| rs75145714 | 2:161845453 | c | t | 0.1264 | intergenic_variant | NA | X |
| rs10866290 | 4:187114479 | c | t | 0.0464 | intron_variant | CYP4V2 | X |
| rs34953939 | 11:47794348 | a | g | 0.0931 | intergenic_variant | NA | X |
| rs62204096 | 20:22938940 | a | g | 0.1565 | downstream_gene_variant | RP11-189G24.2 | X |
| rs189064188 | 9:116253293 | t | c | 0.0804 | intron_variant | RGS3 | X |
| rs6991048 | 8:108347806 | t | c | 0.0904 | intron_variant | ANGPT1 | X |
| rs325247 | 5:143119511 | c | t | 0.0439 | intron_variant | CTB-57H20.1 | X |

| MarkerName | cptid | Allele1 | Allele2 | Effect | Consequence | SYMBOL | 287_snps_used |
|---|---|---|---|---|---|---|---|
| rs35293119 | 2:128133076 | g | c | 0.0442 | intron_variant | MAP3K2 | X |
| rs12636448 | 3:126276710 | c | a | 0.0456 | intron_variant | C3orf22 | X |
| rs75348906 | 11:56735815 | a | t | 0.0695 | upstream_gene_variant | OR5AK3P | X |
| rs12234072 | 5:172785895 | g | t | 0.0661 | downstream_gene_variant | RNU6-500P | X |
| rs79322592 | 1:248028780 | a | c | 0.0517 | intron_variant | TRIM58 | X |
| rs12656497 | 5:32831939 | t | c | 0.0438 | regulatory_region_variant | NA | X |
| rs117564659 | 8:30265541 | g | a | 0.1314 | intron_variant | RBPMS | X |
| rs11170877 | 12:54734289 | a | g | 0.0696 | intron_variant | COPZ1 | X |
| rs74245462 | 15:66430422 | t | g | 0.0503 | intron_variant | MEGF11 | X |
| rs77398404 | 14:26690604 | t | c | 0.0794 | intergenic_variant | NA | X |
| rs187758170 | 19:10688153 | a | g | 0.1465 | intron_variant | AP1M2 | X |
| rs112635299 | 14:94838142 | t | g | 0.1463 | downstream_gene_variant | SERPINA1 | X |
| rs55909816 | 16:81896523 | c | a | 0.0437 | intron_variant | PLCG2 | X |
| rs535527575 | 2:198894550 | g | a | 0.1319 | intron_variant | PLCL1 | |
| rs2531815 | 6:28436060 | t | c | 0.047 | intergenic_variant | NA | X |
| rs138757339 | 8:53204323 | c | t | 0.2102 | intron_variant | ST18 | X |

| MarkerName | cptid | Allele1 | Allele2 | Effect | Consequence | SYMBOL | 287_snps_used |
|---|---|---|---|---|---|---|---|
| rs11759438 | 6:169633335 | c | t | 0.0436 | intron_variant | THBS2 | X |
| rs2032276 | 18:75283432 | g | c | 0.0435 | intergenic_variant | NA | X |
| rs12886724 | 14:100108918 | g | a | 0.0501 | upstream_gene_variant | HHIPL1 | X |
| rs4700642 | 5:63916834 | a | t | 0.0491 | intergenic_variant | NA | X |
| rs8110479 | 19:10734951 | c | t | 0.1131 | upstream_gene_variant | SLC44A2 | X |
| rs12926888 | 16:89265466 | a | g | 0.0449 | downstream_gene_variant | CDH15 | X |
| rs6141600 | 20:34712310 | c | t | 0.0482 | downstream_gene_variant | snoU13 | X |
| rs4893934 | 2:178238316 | g | c | 0.0449 | intron_variant | AC074286.1 | X |
| rs72702145 | 1:169318242 | a | g | 0.1342 | intron_variant | NME7 | X |
| rs11981586 | 7:151028181 | c | t | 0.0657 | intergenic_variant | NA | X |
| rs3116549 | 3:150880908 | c | t | 0.0522 | intron_variant | MED12L | X |
| rs112089121 | 14:83281618 | g | a | 0.0752 | intergenic_variant | NA | X |
| rs9937779 | 16:81844607 | c | t | 0.0568 | intron_variant | PLCG2 | X |
| rs1320969 | 1:169012124 | g | a | 0.0478 | intron_variant | LINC00970 | X |
| rs72897640 | 11:46896126 | t | g | 0.0672 | intron_variant | LRP4 | X |
| rs6434955 | 2:198946551 | g | a | 0.0587 | intron_variant | PLCL1 | X |

| MarkerName | cptid | Allele1 | Allele2 | Effect | Consequence | SYMBOL | 287_snps_used |
|---|---|---|---|---|---|---|---|
| rs6799348 | 3:77048575 | g | a | 0.1488 | intron_variant | ROBO2 | |
| rs216181 | 17:2172753 | a | t | 0.072 | intron_variant | SMG6 | X |
| rs140303646 | 4:79917638 | t | c | 0.218 | intron_variant | LINC01088 | |
| rs118105926 | 8:102881195 | a | g | 0.1379 | intron_variant | NCALD | X |
| rs7137828 | 12:111932800 | c | t | 0.0424 | intron_variant | ATXN2 | X |
| rs1054533 | 19:17004049 | t | c | 0.0451 | missense_variant | CPAMD8 | X |
| rs17383689 | 8:78572803 | g | t | 0.0833 | intergenic_variant | NA | X |
| rs2270744 | 17:8393900 | c | g | 0.0447 | intron_variant | MYH10 | X |
| rs9290227 | 3:94209055 | t | c | 0.0849 | intergenic_variant | NA | X |
| rs3094094 | 6:30639412 | a | g | 0.0793 | intron_variant | DHX16 | X |
| rs7341574 | 8:106573309 | t | c | 0.0452 | intron_variant | ZFPM2 | X |

**Supplemental Table 4. COVID-19 testing rates with 95% confidence intervals for overall MVP cohort and within strata defined by PRS quintiles, FVL carrier status, and *F2* G20210A carrier status.**

| Group | Strata | n Total | n Tested | Percent (95% CI) |
|---|---|---|---|---|
| Overall | Overall | 464961 | 108437 | 23.32% (23.20 - 23.44) |
| PRS Quintiles | q1 | 92993 | 21739 | 23.38% (23.11 - 23.65) |

| | | | | |
|---|---|---|---|---|
| PRS Quintiles | q2 | 92999 | 21584 | 23.21% (22.94 - 23.48) |
| PRS Quintiles | q3 | 92986 | 21608 | 23.24% (22.97 - 23.51) |
| PRS Quintiles | q4 | 92992 | 21863 | 23.51% (23.24 - 23.78) |
| PRS Quintiles | q5 | 92991 | 21643 | 23.27% (23.00 - 23.55) |
| F5 | WT | 439690 | 102485 | 23.31% (23.18 - 23.43) |
| F5 | mutant | 24931 | 5871 | 23.55% (23.02 - 24.08) |
| F2 | WT | 452202 | 105453 | 23.32% (23.20 - 23.44) |
| F2 | mutant | 12700 | 2968 | 23.37% (22.63 - 24.11) |

**Supplemental Table 5. Odds ratios and 95% confidence intervals. Odds ratios (ORs) reported for PRS(VTE) continuous (OR with respect to 1SD increase), PRS(VTE) quintiles, PRS(VTE) top 5% compared to bottom 95%, FVL carriers, F2 G20210A carriers, and outpatient anticoagulation medication usage (anticoagulants) for study outcomes: COVID-19 positive test result, COVID-19 severity (severe or death), pre-index history of VTE (within 2 years of index date), prevalent VTE. Strata denotes stratified models within FVL carriers versus F5 WT individuals, and all participants. Separate analyses are reported for genetic data version 4 and version 4 - v2.1.**

| Outcome | Covariate | Strata | Genetic cohort v4 OR (95%CI) | Genetic cohort v4-v2.1 OR (95%CI) |
|---|---|---|---|---|
| COVID-19 Positive | PRS[1SD] | all | 1.04 (1.02 - 1.06) | 1.02 (0.99 - 1.05) |
| COVID-19 Positive | f5 L vs f5 WT | all | 1.00 (0.91 - 1.10) | 1.03 (0.91 - 1.18) |
| COVID-19 Positive | Anticoagulants (Y/N) | all | 1.07 (1.01 - 1.13) | 1.07 (0.99 - 1.16) |
| COVID-19 Positive | PRS[1SD] | f5 L | 0.97 (0.91 - 1.04) | 0.91 (0.83 - 1.01) |
| COVID-19 Positive | Anticoagulants (Y/N) | f5 L | 1.04 (0.85 - 1.28) | 1.08 (0.80 - 1.45) |
| COVID-19 Positive | PRS[1SD] | f5 WT | 1.05 (1.02 - 1.07) | 1.03 (1.00 - 1.06) |

| **Outcome** | **Covariate** | **Strata** | **Genetic cohort v4 OR (95%CI)** | **Genetic cohort v4-v2.1 OR (95%CI)** |
|---|---|---|---|---|
| COVID-19 Positive | Anticoagulants (Y/N) | f5 WT | 1.07 (1.01 - 1.13) | 1.07 (0.99 - 1.17) |
| COVID-19 Positive | f2 mutant vs WT | all | 0.99 (0.87 - 1.13) | 0.98 (0.81 - 1.17) |
| COVID-19 Positive | PRS[5pct] | all | 1.01 (0.92 - 1.12) | 0.96 (0.84 - 1.11) |
| COVID-19 Positive | PRS[5pct] | f5 L | 0.87 (0.72 - 1.06) | 0.75 (0.57 - 1.00) |
| COVID-19 Positive | PRS[5pct] | f5 WT | 1.07 (0.95 - 1.20) | 1.05 (0.89 - 1.23) |
| COVID-19 Positive | PRS_q2 | all | 1.06 (0.99 - 1.13) | 1.08 (0.99 - 1.19) |
| COVID-19 Positive | PRS_q3 | all | 1.08 (1.01 - 1.16) | 1.08 (0.98 - 1.18) |
| COVID-19 Positive | PRS_q4 | all | 1.15 (1.08 - 1.23) | 1.11 (1.01 - 1.22) |
| COVID-19 Positive | PRS_q5 | all | 1.11 (1.04 - 1.19) | 1.06 (0.97 - 1.17) |

| Outcome | Covariate | Strata | Genetic cohort v4 OR (95%CI) | Genetic cohort v4-v2.1 OR (95%CI) |
|---|---|---|---|---|
| COVID-19 Severe/Death | PRS[1SD] | all | 0.98 (0.91 - 1.05) | 1.01 (0.91 - 1.11) |
| COVID-19 Severe/Death | f5 L vs f5 WT | all | 0.95 (0.69 - 1.29) | 0.81 (0.51 - 1.29) |
| COVID-19 Severe/Death | Anticoagulants (Y/N) | all | 1.16 (0.99 - 1.36) | 1.13 (0.89 - 1.42) |
| COVID-19 Severe/Death | PRS[1SD] | f5 L | 1.21 (0.94 - 1.55) | 1.15 (0.79 - 1.67) |
| COVID-19 Severe/Death | Anticoagulants (Y/N) | f5 L | 1.17 (0.61 - 2.23) | 0.77 (0.27 - 2.22) |
| COVID-19 Severe/Death | PRS[1SD] | f5 WT | 0.96 (0.89 - 1.03) | 1.00 (0.90 - 1.11) |
| COVID-19 Severe/Death | Anticoagulants (Y/N) | f5 WT | 1.17 (0.99 - 1.38) | 1.16 (0.91 - 1.47) |
| COVID-19 Severe/Death | f2 mutant vs WT | all | 0.65 (0.41 - 1.04) | 0.45 (0.20 - 1.02) |
| COVID-19 Severe/Death | PRS[5pct] | all | 1.16 (0.86 - 1.58) | 1.06 (0.67 - 1.67) |

| Outcome | Covariate | Strata | Genetic cohort v4 OR (95%CI) | Genetic cohort v4-v2.1 OR (95%CI) |
|---|---|---|---|---|
| COVID-19 Severe/Death | PRS[5pct] | f5 L | 1.73 (0.94 - 3.19) | 1.07 (0.40 - 2.91) |
| COVID-19 Severe/Death | PRS[5pct] | f5 WT | 1.02 (0.71 - 1.46) | 1.06 (0.63 - 1.77) |
| COVID-19 Severe/Death | PRS_q2 | all | 0.90 (0.73 - 1.12) | 1.00 (0.73 - 1.37) |
| COVID-19 Severe/Death | PRS_q3 | all | 1.02 (0.83 - 1.26) | 1.03 (0.76 - 1.40) |
| COVID-19 Severe/Death | PRS_q4 | all | 0.87 (0.70 - 1.08) | 0.98 (0.71 - 1.33) |
| COVID-19 Severe/Death | PRS_q5 | all | 0.86 (0.69 - 1.07) | 1.00 (0.73 - 1.37) |
| VTE 2yrs Pre-index | PRS[1SD] | all | 1.31 (1.25 - 1.37) | 1.30 (1.22 - 1.38) |
| VTE 2yrs Pre-index | f5 L vs f5 WT | all | 1.71 (1.45 - 2.01) | 1.88 (1.50 - 2.35) |
| VTE 2yrs Pre-index | PRS[1SD] | f5 L | 1.21 (1.09 - 1.36) | 1.24 (1.06 - 1.44) |

| Outcome | Covariate | Strata | Genetic cohort v4 OR (95%CI) | Genetic cohort v4-v2.1 OR (95%CI) |
|---|---|---|---|---|
| VTE<br>2yrs Pre-index | PRS[1SD] | f5 WT | 1.33 (1.27 - 1.40) | 1.31 (1.22 - 1.41) |
| VTE<br>2yrs Pre-index | f2 mutant vs WT | all | 1.51 (1.19 - 1.91) | 1.33 (0.92 - 1.90) |
| VTE<br>2yrs Pre-index | PRS[5pct] | all | 1.79 (1.52 - 2.11) | 1.66 (1.31 - 2.10) |
| VTE<br>2yrs Pre-index | PRS[5pct] | f5 L | 1.56 (1.16 - 2.09) | 1.57 (1.05 - 2.36) |
| VTE<br>2yrs Pre-index | PRS[5pct] | f5 WT | 1.93 (1.59 - 2.34) | 1.74 (1.30 - 2.32) |
| VTE<br>2yrs Pre-index | PRS_q2 | all | 1.16 (0.98 - 1.37) | 1.02 (0.80 - 1.30) |
| VTE<br>2yrs Pre-index | PRS_q3 | all | 1.27 (1.07 - 1.49) | 1.17 (0.93 - 1.48) |
| VTE<br>2yrs Pre-index | PRS_q4 | all | 1.65 (1.41 - 1.93) | 1.53 (1.23 - 1.90) |
| VTE<br>2yrs Pre-index | PRS_q5 | all | 2.06 (1.77 - 2.40) | 1.94 (1.56 - 2.40) |

| Outcome | Covariate | Strata | Genetic cohort v4 OR (95%CI) | Genetic cohort v4-v2.1 OR (95%CI) |
|---|---|---|---|---|
| VTE Prevalent | PRS[1SD] | all | 1.39 (1.36 - 1.42) | 1.35 (1.31 - 1.39) |
| VTE Prevalent | f5 L vs f5 WT | all | 1.72 (1.59 - 1.85) | 1.74 (1.55 - 1.95) |
| VTE Prevalent | PRS[1SD] | f5 L | 1.33 (1.26 - 1.40) | 1.32 (1.22 - 1.43) |
| VTE Prevalent | PRS[1SD] | f5 WT | 1.40 (1.37 - 1.44) | 1.35 (1.31 - 1.40) |
| VTE Prevalent | f2 mutant vs WT | all | 1.75 (1.57 - 1.95) | 1.65 (1.40 - 1.95) |
| VTE Prevalent | PRS[5pct] | all | 1.93 (1.78 - 2.09) | 1.71 (1.52 - 1.94) |
| VTE Prevalent | PRS[5pct] | f5 L | 1.81 (1.58 - 2.09) | 1.68 (1.36 - 2.07) |
| VTE Prevalent | PRS[5pct] | f5 WT | 2.00 (1.81 - 2.20) | 1.74 (1.50 - 2.02) |
| VTE Prevalent | PRS_q2 | all | 1.25 (1.15 - 1.36) | 1.11 (0.99 - 1.25) |

| Outcome | Covariate | Strata | Genetic cohort v4 OR (95%CI) | Genetic cohort v4-v2.1 OR (95%CI) |
|---|---|---|---|---|
| VTE Prevalent | PRS_q3 | all | 1.46 (1.35 - 1.58) | 1.42 (1.27 - 1.59) |
| VTE Prevalent | PRS_q4 | all | 1.82 (1.69 - 1.97) | 1.69 (1.52 - 1.89) |
| VTE Prevalent | PRS_q5 | all | 2.48 (2.31 - 2.67) | 2.24 (2.01 - 2.50) |

### Supplemental Figures

**Supplemental Figure 1. Study cohort flowchart showing analytical sample sizes for each outcome (genetic data version 4 - v2.1).**

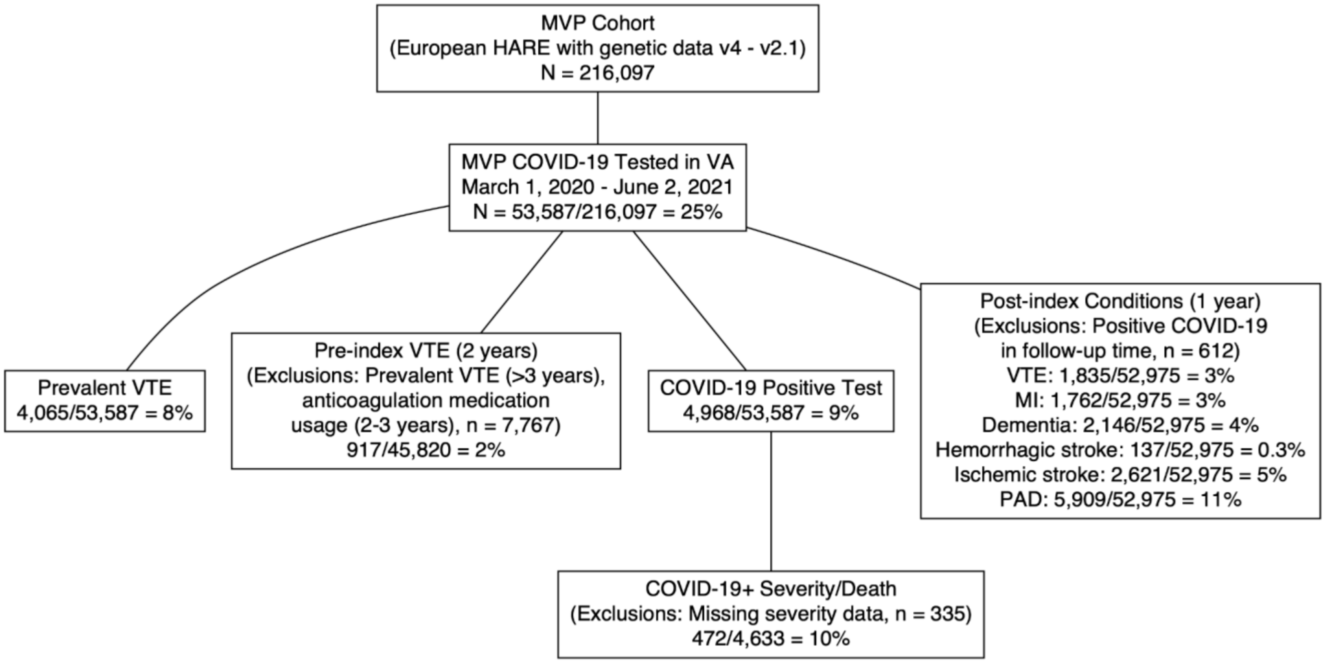

**Supplemental Figure 2. Predicted relative log-odds ratio curves (v4 - v2.1) from spline regression analyses for log-odds of COVID-19 positive test (A) and Prevalent VTE (B), and Pre-index VTE (C). ORs for PRS are compared to scaled PRS value of 0 (mean PRS), from cubic spline models stratified by FVL carrier status. ORs evaluated at median or reference group of other model covariates. Shaded area represents the 95% confidence interval. Models fit in subcohort v4 - v2.1.**

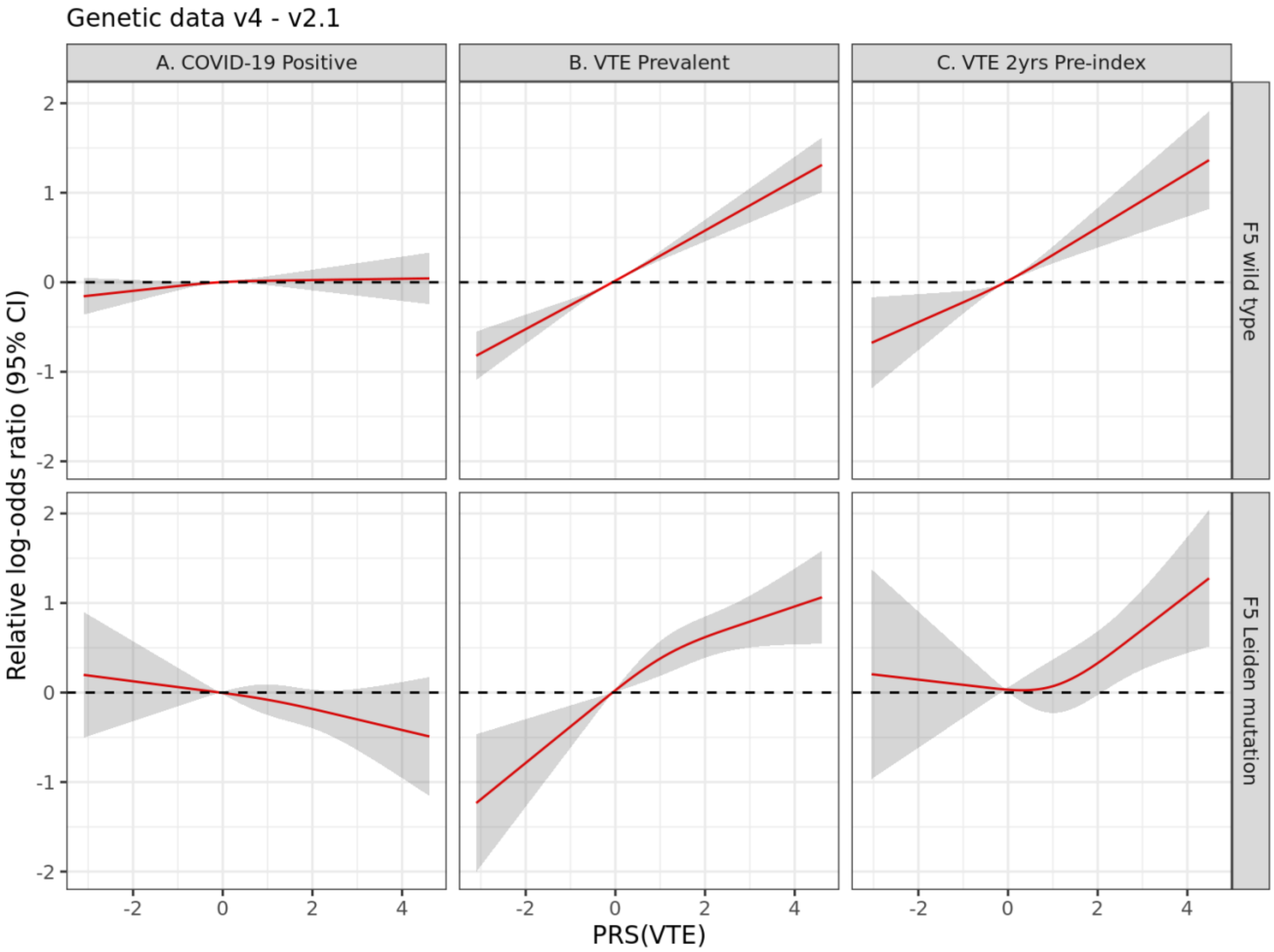

### Appendices

*VA Million Veteran Program COVID-19 Science Initiative Membership & Acknowledgements*


**<u>VA Million Veteran Program COVID-19 Science Initiative</u>**

**MVP COVID-19 Science Program Steering Committee**

- Christopher J. O'Donnell, M.D., M.P.H. (Co-Chair)
  VA Boston Healthcare System, 150 S. Huntington Avenue, Boston, MA 02130
- J. Michael Gaziano, M.D., M.P.H. (Co-Chair)
  VA Boston Healthcare System, 150 S. Huntington Avenue, Boston, MA 02130
- Philip S. Tsao, Ph.D. (Co-Chair)
  VA Palo Alto Health Care System, 3801 Miranda Avenue, Palo Alto, CA 94304
- Sumitra Muralidhar, Ph.D.
  US Department of Veterans Affairs, 810 Vermont Avenue NW, Washington, DC 20420
- Jean Beckham, Ph.D.
  Durham VA Medical Center, 508 Fulton Street, Durham, NC 27705
- Kyong-Mi Chang, M.D.
  Philadelphia VA Medical Center, 3900 Woodland Avenue, Philadelphia, PA 19104
- Juan P. Casas, M.D., Ph.D.
  VA Boston Healthcare System, 150 S. Huntington Avenue, Boston, MA 02130
- Kelly Cho, M.P.H., Ph.D.
  VA Boston Healthcare System, 150 S. Huntington Avenue, Boston, MA 02130
- Saiju Pyarajan, Ph.D.
  VA Boston Healthcare System, 150 S. Huntington Avenue, Boston, MA 02130
- Jennifer Huffman, Ph.D.
  VA Boston Healthcare System, 150 S. Huntington Avenue, Boston, MA 02130
- Jennifer Moser, Ph.D.
  US Department of Veterans Affairs, 810 Vermont Avenue NW, Washington, DC 20420

**MVP COVID-19 Science Program Steering Committee Support**

- Lauren Thomann, M.P.H. (P&P Committee Representative, Working Group Coordinator)
  VA Boston Healthcare System, 150 S. Huntington Avenue, Boston, MA 02130
- Helene Garcon, M.D. (Program Coordinator, Working Group Coordinator)
  VA Boston Healthcare System, 150 S. Huntington Avenue, Boston, MA 02130
- Nicole Kosik, M.P.H. (Working Group Coordinator)
  VA Boston Healthcare System, 150 S. Huntington Avenue, Boston, MA 02130

**MVP COVID-19 Science Program Working Groups and Associated Chairs**

- COVID-19 Related PheWAS
  - Katherine Liao, M.D.
    VA Boston Healthcare System, 150 S. Huntington Avenue, Boston, MA 02130
  - Scott Damrauer, M.D.
    Philadelphia VA Medical Center, 3900 Woodland Avenue, Philadelphia, PA 19104

- Disease Mechanisms
    - Richard Hauger, M.D.
      VA San Diego Healthcare System, 3350 La Jolla Village Drive, San Diego, CA 92161
    - Shiuh-Wen Luoh, M.D., Ph.D.
      Portland VA Medical Center, 3710 SW U.S. Veterans Hospital Road, Portland, OR 97239
    - Sudha Iyengar, Ph.D.
      VA Northeast Ohio Healthcare System, 10701 East Boulevard, Cleveland, OH 44106
- Druggable Genome
    - Juan P. Casas, M.D., Ph.D.
      VA Boston Healthcare System, 150 S. Huntington Avenue, Boston, MA 02130
- Genomics for Risk Prediction, PRS, and MR
    - Themistocles Assimes, M.D., Ph.D.
      VA Palo Alto Health Care System, 3801 Miranda Avenue, Palo Alto, CA 94304
    - Panagiotis Roussos, M.D., Ph.D.
      James J. Peters VA Medical Center, 130 W Kingsbridge Rd, Bronx, NY 10468
    - Robert Striker, M.D., Ph.D.
      William S. Middleton Memorial Veterans Hospital, 2500 Overlook Terrace, Madison, WI 53705
- GWAS & Downstream Analysis
    - Jennifer Huffman, Ph.D.
      VA Boston Healthcare System, 150 S. Huntington Avenue, Boston, MA 02130
    - Yan Sun, Ph.D.
      Atlanta VA Medical Center, 1670 Clairmont Road, Decatur, GA 30033
- Pharmacogenomics
    - Adriana Hung, M.D., M.P.H.
      VA Tennessee Valley Healthcare System, 1310 24th Avenue, South Nashville, TN 37212
    - Sony Tuteja, Pharm.D., M.S.
      Philadelphia VA Medical Center, 3900 Woodland Avenue, Philadelphia, PA 19104
- VA COVID-19 Shared Data Resource – Scott L. DuVall, Ph.D.; Kristine E. Lynch, Ph.D.; Elise Gatsby, M.P.H.
  VA Informatics and Computing Infrastructure (VINCI), VA Salt Lake City Health Care System, 500 Foothill Drive, Salt Lake City, UT 84148
- MVP COVID-19 Data Core – Kelly Cho, M.P.H., Ph.D.; Lauren Costa, M.P.H.; Anne Yuk-Lam Ho, M.P.H.; Rebecca Song, M.P.H.
  VA Boston Healthcare System, 150 S. Huntington Avenue, Boston, MA 02130

**<u>VA Million Veteran Program</u>**

**MVP Executive Committee**

- Co-Chair: J. Michael Gaziano, M.D., M.P.H.

VA Boston Healthcare System, 150 S. Huntington Avenue, Boston, MA 02130

- Co-Chair: Sumitra Muralidhar, Ph.D.
  US Department of Veterans Affairs, 810 Vermont Avenue NW, Washington, DC 20420
- Rachel Ramoni, D.M.D., Sc.D., Chief VA Research and Development Officer
  US Department of Veterans Affairs, 810 Vermont Avenue NW, Washington, DC 20420
- Jean Beckham, Ph.D.
  Durham VA Medical Center, 508 Fulton Street, Durham, NC 27705
- Kyong-Mi Chang, M.D.
  Philadelphia VA Medical Center, 3900 Woodland Avenue, Philadelphia, PA 19104
- Christopher J. O'Donnell, M.D., M.P.H.
  VA Boston Healthcare System, 150 S. Huntington Avenue, Boston, MA 02130
- Philip S. Tsao, Ph.D.
  VA Palo Alto Health Care System, 3801 Miranda Avenue, Palo Alto, CA 94304
- James Breeling, M.D., Ex-Officio
  US Department of Veterans Affairs, 810 Vermont Avenue NW, Washington, DC 20420
- Grant Huang, Ph.D., Ex-Officio
  US Department of Veterans Affairs, 810 Vermont Avenue NW, Washington, DC 20420
- Juan P. Casas, M.D., Ph.D., Ex-Officio
  VA Boston Healthcare System, 150 S. Huntington Avenue, Boston, MA 02130

**MVP Program Office**

- Sumitra Muralidhar, Ph.D.
  US Department of Veterans Affairs, 810 Vermont Avenue NW, Washington, DC 20420
- Jennifer Moser, Ph.D.
  US Department of Veterans Affairs, 810 Vermont Avenue NW, Washington, DC 20420

**MVP Recruitment/Enrollment**

- Recruitment/Enrollment Director/Deputy Director, Boston – Stacey B. Whitbourne, Ph.D.; Jessica V. Brewer, M.P.H.
  VA Boston Healthcare System, 150 S. Huntington Avenue, Boston, MA 02130
- MVP Coordinating Centers
  - Clinical Epidemiology Research Center (CERC), West Haven – Mihaela Aslan, Ph.D.
    West Haven VA Medical Center, 950 Campbell Avenue, West Haven, CT 06516
  - Cooperative Studies Program Clinical Research Pharmacy Coordinating Center, Albuquerque – Todd Connor, Pharm.D.; Dean P. Argyres, B.S., M.S.
    New Mexico VA Health Care System, 1501 San Pedro Drive SE, Albuquerque, NM 87108
  - Genomics Coordinating Center, Palo Alto – Philip S. Tsao, Ph.D.
    VA Palo Alto Health Care System, 3801 Miranda Avenue, Palo Alto, CA 94304
  - MVP Boston Coordinating Center, Boston - J. Michael Gaziano, M.D., M.P.H.
    VA Boston Healthcare System, 150 S. Huntington Avenue, Boston, MA 02130
  - MVP Information Center, Canandaigua – Brady Stephens, M.S.
    Canandaigua VA Medical Center, 400 Fort Hill Avenue, Canandaigua, NY 14424

- VA Central Biorepository, Boston – Mary T. Brophy M.D., M.P.H.; Donald E. Humphries, Ph.D.; Luis E. Selva, Ph.D.
  VA Boston Healthcare System, 150 S. Huntington Avenue, Boston, MA 02130
- MVP Informatics, Boston – Nhan Do, M.D.; Shahpoor (Alex) Shayan, M.S.
  VA Boston Healthcare System, 150 S. Huntington Avenue, Boston, MA 02130
- MVP Data Operations/Analytics, Boston – Kelly Cho, M.P.H., Ph.D.
  VA Boston Healthcare System, 150 S. Huntington Avenue, Boston, MA 02130
- Director of Regulatory Affairs – Lori Churby, B.S.
  VA Palo Alto Health Care System, 3801 Miranda Avenue, Palo Alto, CA 94304

**MVP Science**
- Science Operations – Christopher J. O'Donnell, M.D., M.P.H.
  VA Boston Healthcare System, 150 S. Huntington Avenue, Boston, MA 02130
- Genomics Core - Christopher J. O'Donnell, M.D., M.P.H.
  VA Boston Healthcare System, 150 S. Huntington Avenue, Boston, MA 02130
  Saiju Pyarajan Ph.D.
  VA Boston Healthcare System, 150 S. Huntington Avenue, Boston, MA 02130
  Philip S. Tsao, Ph.D.
  VA Palo Alto Health Care System, 3801 Miranda Avenue, Palo Alto, CA 94304
- Data Core - Kelly Cho, M.P.H, Ph.D.
  VA Boston Healthcare System, 150 S. Huntington Avenue, Boston, MA 02130
- VA Informatics and Computing Infrastructure (VINCI) – Scott L. DuVall, Ph.D.
  VA Salt Lake City Health Care System, 500 Foothill Drive, Salt Lake City, UT 84148
- Data and Computational Sciences – Saiju Pyarajan, Ph.D.
  VA Boston Healthcare System, 150 S. Huntington Avenue, Boston, MA 02130
- Statistical Genetics – Elizabeth Hauser, Ph.D.
  Durham VA Medical Center, 508 Fulton Street, Durham, NC 27705
  Yan Sun, Ph.D.
  Atlanta VA Medical Center, 1670 Clairmont Road, Decatur, GA 30033
  Hongyu Zhao, Ph.D.
  West Haven VA Medical Center, 950 Campbell Avenue, West Haven, CT 06516

**Current MVP Local Site Investigators**
- Atlanta VA Medical Center (Peter Wilson, M.D.)
  1670 Clairmont Road, Decatur, GA 30033
- Bay Pines VA Healthcare System (Rachel McArdle, Ph.D.)
  10,000 Bay Pines Blvd Bay Pines, FL 33744
- Birmingham VA Medical Center (Louis Dellitalia, M.D.)
  700 S. 19th Street, Birmingham AL 35233
- Central Western Massachusetts Healthcare System (Kristin Mattocks, Ph.D., M.P.H.)
  421 North Main Street, Leeds, MA 01053
- Cincinnati VA Medical Center (John Harley, M.D., Ph.D.)
  3200 Vine Street, Cincinnati, OH 45220
- Clement J. Zablocki VA Medical Center (Jeffrey Whittle, M.D., M.P.H.)

5000 West National Avenue, Milwaukee, WI 53295
- VA Northeast Ohio Healthcare System (Frank Jacono, M.D.)
  10701 East Boulevard, Cleveland, OH 44106
- Durham VA Medical Center (Jean Beckham, Ph.D.)
  508 Fulton Street, Durham, NC 27705
- Edith Nourse Rogers Memorial Veterans Hospital (John Wells., Ph.D.)
  200 Springs Road, Bedford, MA 01730
- Edward Hines, Jr. VA Medical Center (Salvador Gutierrez, M.D.)
  5000 South 5th Avenue, Hines, IL 60141
- Veterans Health Care System of the Ozarks (Gretchen Gibson, D.D.S., M.P.H.)
  1100 North College Avenue, Fayetteville, AR 72703
- Fargo VA Health Care System (Kimberly Hammer, Ph.D.)
  2101 N. Elm, Fargo, ND 58102
- VA Health Care Upstate New York (Laurence Kaminsky, Ph.D.)
  113 Holland Avenue, Albany, NY 12208
- New Mexico VA Health Care System (Gerardo Villareal, M.D.)
  1501 San Pedro Drive, S.E. Albuquerque, NM 87108
- VA Boston Healthcare System (Scott Kinlay, M.B.B.S., Ph.D.)
  150 S. Huntington Avenue, Boston, MA 02130
- VA Western New York Healthcare System (Junzhe Xu, M.D.)
  3495 Bailey Avenue, Buffalo, NY 14215-1199
- Ralph H. Johnson VA Medical Center (Mark Hamner, M.D.)
  109 Bee Street, Mental Health Research, Charleston, SC 29401
- Columbia VA Health Care System (Roy Mathew, M.D.)
  6439 Garners Ferry Road, Columbia, SC 29209
- VA North Texas Health Care System (Sujata Bhushan, M.D.)
  4500 S. Lancaster Road, Dallas, TX 75216
- Hampton VA Medical Center (Pran Iruvanti, D.O., Ph.D.)
  100 Emancipation Drive, Hampton, VA 23667
- Richmond VA Medical Center (Michael Godschalk, M.D.)
  1201 Broad Rock Blvd., Richmond, VA 23249
- Iowa City VA Health Care System (Zuhair Ballas, M.D.)
  601 Highway 6 West, Iowa City, IA 52246-2208
- Eastern Oklahoma VA Health Care System (Douglas Ivins, M.D.)
  1011 Honor Heights Drive, Muskogee, OK 74401
- James A. Haley Veterans' Hospital (Stephen Mastorides, M.D.)
  13000 Bruce B. Downs Blvd, Tampa, FL 33612
- James H. Quillen VA Medical Center (Jonathan Moorman, M.D., Ph.D.)
  Corner of Lamont & Veterans Way, Mountain Home, TN 37684
- John D. Dingell VA Medical Center (Saib Gappy, M.D.)
  4646 John R Street, Detroit, MI 48201
- Louisville VA Medical Center (Jon Klein, M.D., Ph.D.)
  800 Zorn Avenue, Louisville, KY 40206
- Manchester VA Medical Center (Nora Ratcliffe, M.D.)

718 Smyth Road, Manchester, NH 03104

- Miami VA Health Care System (Hermes Florez, M.D., Ph.D.)
  1201 NW 16th Street, 11 GRC, Miami FL 33125
- Michael E. DeBakey VA Medical Center (Olaoluwa Okusaga, M.D.)
  2002 Holcombe Blvd, Houston, TX 77030
- Minneapolis VA Health Care System (Maureen Murdoch, M.D., M.P.H.)
  One Veterans Drive, Minneapolis, MN 55417
- N. FL/S. GA Veterans Health System (Peruvemba Sriram, M.D.)
  1601 SW Archer Road, Gainesville, FL 32608
- Northport VA Medical Center (Shing Shing Yeh, Ph.D., M.D.)
  79 Middleville Road, Northport, NY 11768
- Overton Brooks VA Medical Center (Neeraj Tandon, M.D.)
  510 East Stoner Ave, Shreveport, LA 71101
- Philadelphia VA Medical Center (Darshana Jhala, M.D.)
  3900 Woodland Avenue, Philadelphia, PA 19104
- Phoenix VA Health Care System (Samuel Aguayo, M.D.)
  650 E. Indian School Road, Phoenix, AZ 85012
- Portland VA Medical Center (David Cohen, M.D.)
  3710 SW U.S. Veterans Hospital Road, Portland, OR 97239
- Providence VA Medical Center (Satish Sharma, M.D.)
  830 Chalkstone Avenue, Providence, RI 02908
- Richard Roudebush VA Medical Center (Suthat Liangpunsakul, M.D., M.P.H.)
  1481 West 10th Street, Indianapolis, IN 46202
- Salem VA Medical Center (Kris Ann Oursler, M.D.)
  1970 Roanoke Blvd, Salem, VA 24153
- San Francisco VA Health Care System (Mary Whooley, M.D.)
  4150 Clement Street, San Francisco, CA 94121
- South Texas Veterans Health Care System (Sunil Ahuja, M.D.)
  7400 Merton Minter Boulevard, San Antonio, TX 78229
- Southeast Louisiana Veterans Health Care System (Joseph Constans, Ph.D.)
  2400 Canal Street, New Orleans, LA 70119
- Southern Arizona VA Health Care System (Paul Meyer, M.D., Ph.D.)
  3601 S 6th Avenue, Tucson, AZ 85723
- Sioux Falls VA Health Care System (Jennifer Greco, M.D.)
  2501 W 22nd Street, Sioux Falls, SD 57105
- St. Louis VA Health Care System (Michael Rauchman, M.D.)
  915 North Grand Blvd, St. Louis, MO 63106
- Syracuse VA Medical Center (Richard Servatius, Ph.D.)
  800 Irving Avenue, Syracuse, NY 13210
- VA Eastern Kansas Health Care System (Melinda Gaddy, Ph.D.)
  4101 S 4th Street Trafficway, Leavenworth, KS 66048
- VA Greater Los Angeles Health Care System (Agnes Wallbom, M.D., M.S.)
  11301 Wilshire Blvd, Los Angeles, CA 90073
- VA Long Beach Healthcare System (Timothy Morgan, M.D.)

5901 East 7th Street Long Beach, CA 90822

- VA Maine Healthcare System (Todd Stapley, D.O.)
  1 VA Center, Augusta, ME 04330
- VA New York Harbor Healthcare System (Scott Sherman, M.D., M.P.H.)
  423 East 23rd Street, New York, NY 10010
- VA Pacific Islands Health Care System (George Ross, M.D.)
  459 Patterson Rd, Honolulu, HI 96819
- VA Palo Alto Health Care System (Philip Tsao, Ph.D.)
  3801 Miranda Avenue, Palo Alto, CA 94304-1290
- VA Pittsburgh Health Care System (Patrick Strollo, Jr., M.D.)
  University Drive, Pittsburgh, PA 15240
- VA Puget Sound Health Care System (Edward Boyko, M.D.)
  1660 S. Columbian Way, Seattle, WA 98108-1597
- VA Salt Lake City Health Care System (Laurence Meyer, M.D., Ph.D.)
  500 Foothill Drive, Salt Lake City, UT 84148
- VA San Diego Healthcare System (Samir Gupta, M.D., M.S.C.S.)
  3350 La Jolla Village Drive, San Diego, CA 92161
- VA Sierra Nevada Health Care System (Mostaqul Huq, Pharm.D., Ph.D.)
  975 Kirman Avenue, Reno, NV 89502
- VA Southern Nevada Healthcare System (Joseph Fayad, M.D.)
  6900 North Pecos Road, North Las Vegas, NV 89086
- VA Tennessee Valley Healthcare System (Adriana Hung, M.D., M.P.H.)
  1310 24th Avenue, South Nashville, TN 37212
- Washington DC VA Medical Center (Jack Lichy, M.D., Ph.D.)
  50 Irving St, Washington, D. C. 20422
- W.G. (Bill) Hefner VA Medical Center (Robin Hurley, M.D.)
  1601 Brenner Ave, Salisbury, NC 28144
- White River Junction VA Medical Center (Brooks Robey, M.D.)
  163 Veterans Drive, White River Junction, VT 05009
- William S. Middleton Memorial Veterans Hospital (Robert Striker, M.D., Ph.D.)
  2500 Overlook Terrace, Madison, WI 53705
